# Identification of ultra-rare genetic variants in Pediatric Acute Onset Neuropsychiatric Syndrome (PANS) by exome and whole genome sequencing

**DOI:** 10.1101/2021.05.25.21257256

**Authors:** Rosario Trifiletti, Herbert M. Lachman, Olivia Manusama, Deyou Zheng, Alberto Spalice, Pietro Chiurazzi, Allan Schornagel, Andreea M. Serban, Rogier van Wijck, Sigrid Swagemakers, Peter J. van der Spek

**Author notes:** These authors contributed equally to this work. Department of Psychiatry and Behavioral Sciences, Albert Einstein College Medicine. Bronx, New York, 10461, U.S.A.

## Abstract

Pediatric acute onset neuropsychiatric syndrome (PANS) is viewed as an autoimmune/autoinflammatory condition characterized by the abrupt onset of severe neurological and psychiatric symptoms, in particular obsessive-compulsive disorder (OCD), tics, anxiety, mood swings, irritability, and restricted eating, often triggered by infections. However, direct evidence of autoimmunity, infections, or a proinflammatory state is often lacking, and there is no unifying pathogenic pathway. This could be due to underlying genetic heterogeneity, which could lead to the development of PANS through different cellular and molecular pathways. Unfortunately, little is known about the genetic basis of PANS. Consequently, we carried out whole exome sequencing (WES) on a U.S. cohort of 386 cases who met diagnostic criteria for PANS, including 133 family triads, and whole genome sequencing (WGS) on ten cases from the European Union, who were selected for WGS because of severe PANS symptoms. We focused on identifying potentially deleterious genetic variants that were either *de novo* or ultra-rare with a minor allele frequency (MAF) < 0.001. Candidate mutations were found in 11 genes: *PPM1D, SGCE, PLCG2, NLRC4, CACNA1B, SHANK3, CHK2, GRIN2A*, *RAG1*, *GABRG2*, and *SYNGAP1* in a total of 20 cases, which included two sets of siblings, and two or more unrelated subjects with ultra-rare variants in *SGCE, NLRC4, RAG1,* and *SHANK3.* The PANS candidate genes we identified separate into two broad functional categories. One group regulates peripheral innate and adaptive immune responses (e.g., *PPM1D, CHK2, NLRC4, RAG1, PLCG2*), some of which also influence microglia function. Another is expressed primarily at neuronal synapses or directly modulates synaptic function (*SHANK3, SYNGAP1, GRIN2A, GABRG2, CACNA1B, SGCE*). These neuronal PANS candidate genes are often mutated in autism spectrum disorder, developmental disorders, and myoclonus-dystonia. In fact, eight out of 20 cases in this study developed PANS superimposed on a preexisting neurodevelopmental disorder. There is, however, clinical overlap between these two groups and some crossover expression (e.g., some neuronal genes are expressed in immune cells and vice versa) that diminishes the neuronal/immune dichotomy. Genes in both categories are also highly expressed in the enteric nervous system, and in the choroid plexus and brain vasculature, suggesting they might contribute to a breach in the blood-CSF barrier and blood-brain barrier (BBB) that would permit the entry of autoantibodies, inflammatory cytokines, chemokines, prostaglandins, and autoantibodies into the brain. Thus, PANS is a genetically heterogeneous condition that can occur as a stand-alone neuropsychiatric condition or co-morbid with neurodevelopmental disorders, with candidate genes functioning at several levels of the neuroinflammatory axis.

## Introduction

The clinical entity known as PANS emerged from problems related to the diagnosis of PANDAS (Pediatric Autoimmune Neuropsychiatric Disorders Associated with Streptococcal Infections). In its original description, PANDAS was characterized by acute onset obsessive-compulsive disorder (OCD) and tics following infection with group A beta-hemolytic streptococcus (GABHS)^1, 2^. The concept of PANS was introduced a decade later to expand the diagnosis to include a multitude of acute-onset neuropsychiatric problems and exclude the requirement of GABHS infection and tics (**Table 1**) ^3^. PANDAS is now considered as a subgroup of PANS, although some controversy remains. While some investigators have reported the presence of autoantibodies against neuronal antigens and elevated levels of inflammatory cytokines in PANS, these findings are inconsistent and the validity of the Cunningham panel of 5 biomarkers used in diagnosing PANS has been supported in some studies and questioned in others ^4–6^. Imaging studies in some PANS cases show local inflammation in the thalamus, basal ganglia, and amygdala, supporting a neuroinflammatory or autoimmune etiology. In addition, many patients respond to immunomodulators, such as non-steroidal anti-inflammatory drugs (NSAIDs), intravenous immunoglobulin (IVIG), corticosteroids, and more recently, the B-cell inhibitor Rituximab ^4, 8^. However, this finding is controversial as some investigators have expressed concerns about the dearth of large, well-controlled clinical trials ^6, 7, 9^. Because of these issues and the lack of consistent, objective biomarkers, many children remain undiagnosed and undertreated, adversely affecting both patients and their families, and perhaps contributing to disease chronicity. According to the Stanford PANS Clinic, the median Caregiver Burden Inventory (CBI) during a 1st PANS flare is 37, which is higher than the CBI for those caring for someone with Alzheimer disease and similar to that found in Rett syndrome ^10^. Although most children with PANS show significant improvement at follow-up, according to a recently published longitudinal study, full remission was rare, and more than one-third were classified as having a chronic course ^11^.

**Table 1.**
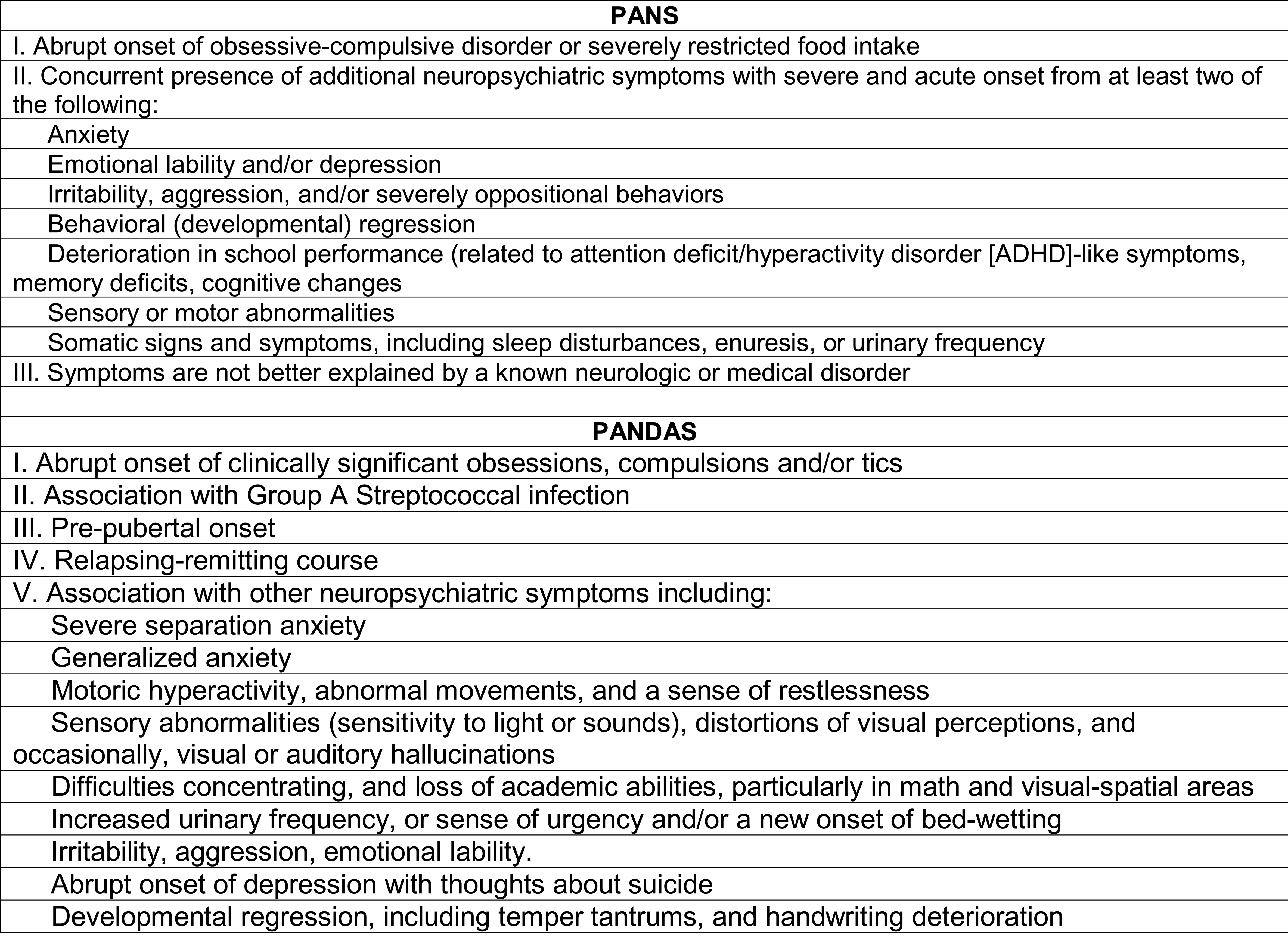
Diagnostic criteria for PANS/PANDAS adapted from reference 3 and NIMH (see Web Resources)

One plausible explanation for the inconsistent findings related to the presence or absence of autoantibodies and inflammatory markers, and the relationship to infectious pathogens, is heterogeneity in the infectious diseases other than GABHS, such as *Mycoplasma pneumoniae, Borrelia burgdorferi* and influenza virus, that might act as autoimmune or proinflammatory triggers ^12^. Another possibility is that non-infectious environmental factors that activate innate immune pathways (i.e. sterile or non-infectious inflammation), such as oxidative stress, exposure to toxins, or emotional stress, could trigger an abnormal inflammatory response in genetically susceptible children. Finally, heterogeneity in genetic risk factors that cause dysregulation of peripheral immunity, or central neuronal/innate immune pathways could be at play, all leading to a common clinical phenotype. In this model, some genetic subgroups will express classic markers of inflammation or autoantibodies, but not others. However, gene discovery in PANS that can test this model is still in its infancy.

Whole exome sequencing (WES) and whole genome sequencing (WGS) are gene discovery tools used by researchers, and increasingly by clinicians, to identify ultra-rare, biologically powerful genetic factors underlying disease states. However, such studies have not yet been reported in PANS. We now describe the discovery of ultra-rare variants in 11 genes in 20 PANS cases using WES and WGS technologies.

## Subjects and Methods

### Subjects

The European subjects were identified through a call for patients with severe symptoms from a PANS/PANDAS advocacy group called EXPAND after a teenage girl with chronic PANS was found to have an ultra-rare variant in the *PPM1D* gene by WGS. EXPAND is a European advocacy organization for families of children and adolescents with immune-mediated neuropsychiatric disorders. Parents, caretakers, or patients signed informed consents approved by the Ethical Committee at Erasmus MC (MEC-2011-253 for control samples and MEC-2021-0359 for PANS patients; IRB/ Human Subject Assurance number/Federal Wide Assurance, FWA00001336). Histories from this cohort were obtained by the participating physicians and collated by one of the co-authors (O.M).

Cases from the U.S. were obtained from a large private practice run by one of the authors (R.T) devoted to PANS (PANDAS/PANS Institute, Ramsey, NJ). 383 PANS cases and 263 controls that included 133 triads had WES carried out by Centogene (see below) (Rostock, Germany). The subjects signed an informed consent by Centogene to which they agreed to provide anonymized genetic information for research purposes. The corresponding author has IRB approval from the Albert Einstein College of Medicine to obtain and analyze deidentified data from human subjects (approval ID, 2000-014). Each case in the U.S. cohort was personally treated by one physician (R.T) who obtained a detailed personal and family history.

To protect patient confidentiality, detailed histories are only available upon request to the corresponding author. The consenting parents and participants have agreed to have the results of this research work published.

### WGS and WES data analysis

The EU cases were sequenced using DAB nanoball sequencing for WGS and Illumina for WES. We implemented ANNOVAR, an efficient software tool that utilizes up-to-date information to functionally annotate genetic variants and allele frequencies detected in each patient’s genome. In addition to gene-based annotation, ANNOVAR is used for region-based annotations to identify variants in specific genomic regions, such as ultra-conserved regions across species (Vista database), predicted transcription factor binding sites (Transfac), segmental duplication regions, genome wide association study (GWAS) hits, database of genomic variants, DNAse I hypersensitivity sites, ENCODE H3K4Me1/H3K4Me3/H3K27Ac/CTCF sites, ChIP-Seq peaks, RNA-Seq peaks, and other annotations in genomic intervals. DNA nanoball sequencing has been applied for WGS to study non-coding regions and in particular variants within the ultra-conserved regulatory regions. This provided highly reliable variant lists that were compared with our selected variants and filtered based on allele frequency in GNOMAD ClinVar (public NCBI repository derived data). Only variants that occurred in patients and are never detected in 597 healthy elderly (Welderly) sample genomes were considered ^13^. DNA Nanoball sequencing protocol is detailed in the classic paper by Drmanac et al ^14^. The ultra-rare variant assessment was confirmed using additional cohorts that have been sequenced by the Erasmus MC team headed by one of us (P.S.).

The U.S. samples were sequenced by Centogene using double stranded DNA capture baits against approximately 36.5 Mb of the human coding exome (targeting >98% of the coding elements). RefSeq from the human genome build GRCh37/hg19) are used to enrich target regions from fragmented genomic DNA with the Twist Human Core Exome Plus kit. The generated library was sequenced on an Illumina platform to obtain at least 20x coverage depth for >98% of the targeted bases. The investigation for relevant variants focused on coding exons and flanking +/-20 intronic nucleotides of genes with a clear gene-phenotype evidence (based on OMIM® information). The resulting variant call (VCF) file was then subjected to a custom pipeline developed at the PANDAS/PANS Institute. For this study, we searched for variants with all of the following properties: 1) minor allele frequency (MAF) of <0.001: 2) American College of Medical Genetics Class 5 or 4 (i.e., “pathogenic” or “likely pathogenic”): 3) among a PANDAS/PANS institute cohort of 383 cases and 263 controls variants that were never observed, with rare exceptions noted, in controls.

### Network analysis

Functional analysis was performed within Ingenuity Pathway Analysis (IPA) (QIAGEN Inc.). All candidate genes were imported in IPA to assess the pathways involved. Two additional, unpublished, PANS candidate genes (*MTHC2* and *BID*) were added to the analysis. Using the network tool within IPA, a connectivity network was constructed based on the IPA/QIAGEN Knowledge Base. Both direct and indirect relationships were used to construct the biological network.

### RNA expression analysis

The gene expression profiles of the candidate genes were analyzed using several datasets. First, we hypothesized that microglia might play a critical role in the PANS phenotype especially after an inflammatory stimulus. Consequently, a subset of 159 autism and pediatric immune disorder genes was analyzed based on a public dataset investigating the effect of LPS on murine microglia and macrophages (GSE102482) ^15^. After Robust Multichip Average normalization (RMA), a statistical analysis of microarray (SAM) was performed between non-stimulated microglia and LPS-stimulated microglia within OmniViz version 6.1.13.0 (Instem Scientific). The top 15 up-regulated and down-regulated genes were selected for visualization and further analysis.

Next, we investigated the gene expression profiles of our 11 PANS-associated candidates in peripheral blood mononuclear cells (PBMCs) under baseline conditions and after infection as many patients develop PANS symptoms following infections. Given the rising reporting of neuropsychiatric symptoms after COVID-19 infection, we used a recently published single cell RNA-seq (sc-RNA-seq) COVID-19 dataset to investigate the gene expression profiles^16^. This database contains gene expression patterns in control samples (N=6) and hospitalized severe COVID-19 patients (N=7).

To investigate the expression of the candidate genes in normal human tissue we used the Genotype-Tissue Expression (GTEx) dataset ^17^. We used the GTEx Multi Gene Query available on the GTEx portal to construct the RNA profiles in normal human tissue lineages. Finally, as the GTEx portal uses bulk RNA-seq data, we wanted to further investigate the gene expression profiles in more detail. Because neurological and psychiatric symptoms are cardinal in PANS, we were particularly interested in scRNA-seq in brain tissue. scRNA-seq data is not yet available for human brains, however, recently two datasets were published containing scRNA-seq data for adolescent and fetal mouse brains^18, 19^. All candidate genes were investigated in both datasets on the Mouse Brain Atlas portal.

## Results

We identified ultra-rare variants in 11 genes (*PPM1D, SGCE, PLCG2, NLRC4, CACNA1B, SHANK3, CHK2, GRIN2A, RAG1, GABRG2, and SYNGAP1*) in 20 patients who met diagnostic criteria for PANS as established by the PANS Consensus Conference ^3^. The ultra-rare variants and brief descriptions of the cases are shown on **Figure 1** and **Table 2**, respectively. In the European cohort, two affected siblings (cases 1 and 2) were found with an ultra-rare missense mutation in *PPM1D* (c.131C>G; p.S44W). They inherited the variant from an asymptomatic parent who has a family history autoimmune disorders, a common scenario in PANS families ^20, 21^. S44W is a known single nucleotide variant (SNV), rs373862041, with a minor allele frequency (MAF) of 0.000255, based on the TOPMed database of more than 125,000 samples ^22^. The ultra-rare variant in *CACNA1B,* found in two affected siblings (cases 3 and 4), is a 48 bp insert at the exon 2 splice donor site (c.390+1) that is predicted to disrupt splicing. It is part of a set of multiallelic insertion variants, rs370237172, that has an overall MAF of 0.017. However, the 48 bp insert is ultra-rare and has not been observed in control data sets. We have been unable to genotype the parents. Interestingly, the same 48 bp insert was identified in a young woman who has been incapacitated with chronic fatigue syndrome (myalgic encephalomyelitis) and anorexia (unpublished observations). This insert shows a 100% hit with a human oral metagenome assembly (unpublished results from ME/CFS project headed by Dr. Rogier Louwen, Erasmus MC).

**Figure 1.**
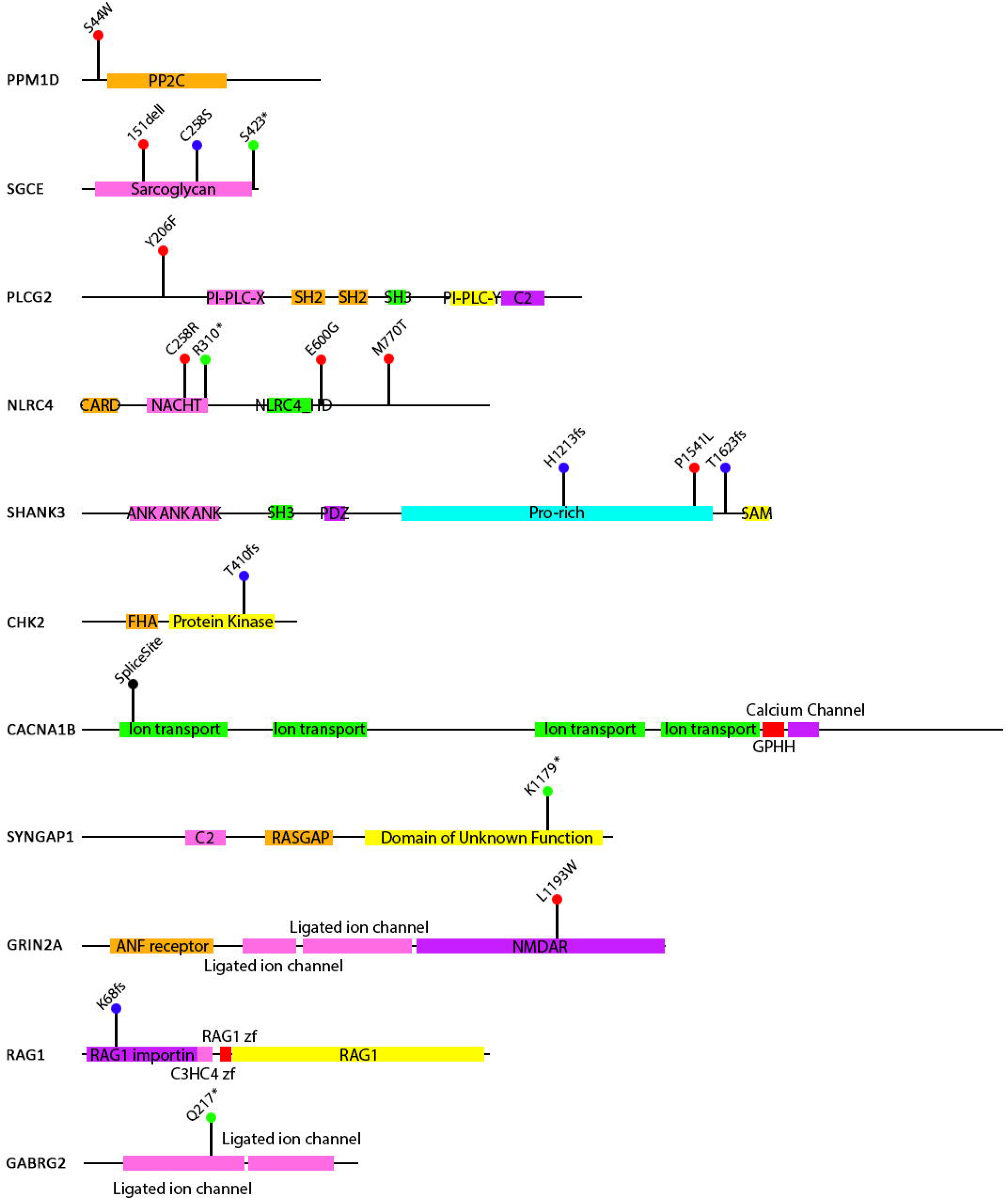
Position of PANS ultra-rare variants within each candidate gene

**Table 2.**
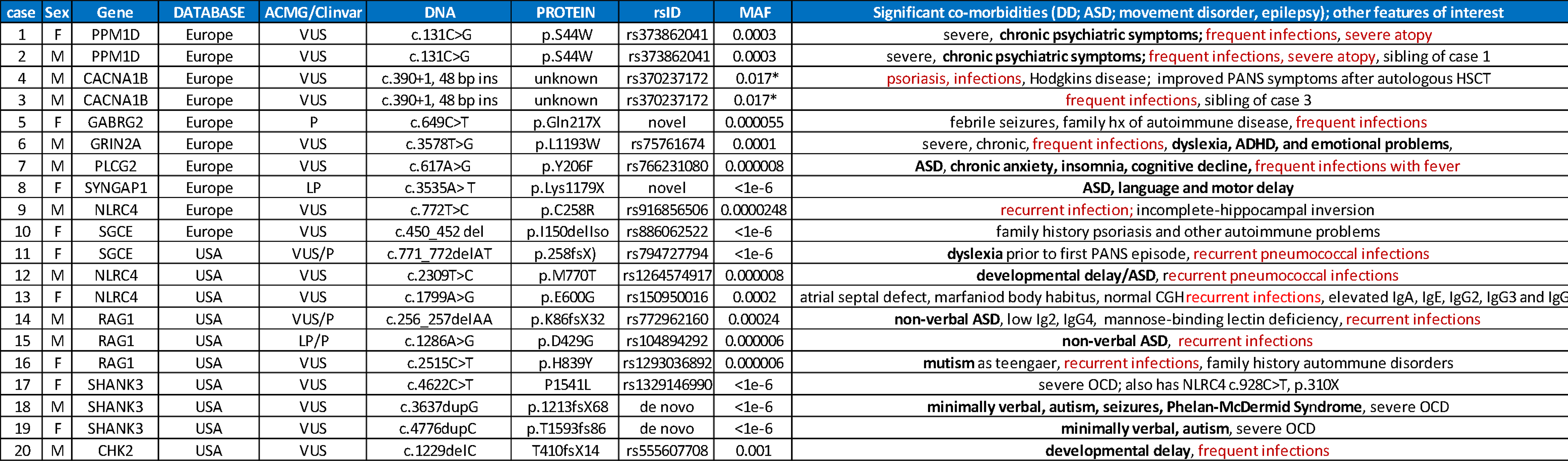
Summary of cases. Patient demographics, genetic variants, and brief clinical description. Bold type highlights cases with comorbid neurodevelopmental disorders, red type highlights those with a history of recurrent infections. For complete patient histories, see Supplemental Note: Case Reports. Abbreviations: pathogenic (P); likely pathogenic (LP); variant of unknown significance (VUS). To protect patient confidentiality, ages are not included. Each case showed the first signs of PANS before the age of 10.

Known transmission from a parent in the European cohort was also found in cases 9 and 10 (*NLRC4*: c.772T>C; p.C258R; and *SGCE*: del150 Iso; c.450_452, respectively), which will be discussed below. For the other ultra-rare variants, parental genotypes are not available. Remarkably, in the U.S. cohort, different ultra-rare variants were found in two of the same genes identified in the European cohort (*NLRC4* and *SGC2*), and another, *CHK2*, codes for a well-established PPM1D substrate ^23^. A total of four different candidate variants were found in *NLRC4*, the most for any PANS-associated gene. Ultra-rare variants were also found in *SHANK3* and *RAG1* gene in three patients each. Case 17 has an ultra-rare variant in *SHANK3* (c.4622C>T, P1541L) and a nonsense mutation in *NLRC4* (c.928C>T, p.310X).

The finding of ultra-rare mutations in the same genes in unrelated individuals points to a true association.

These 11 genes separate into two broad functional categories; those that affect peripheral innate and adaptive immune pathways, and those that are expressed primarily in cortical neurons, where they function as synaptic regulators. Some of the immune genes also have significant effects on microglia, and the synaptic genes have all been implicated in other neurological and neurodevelopmental disorders. In addition, several neuronal genes are expressed in the choroid plexus and brain vasculature, suggesting they might contribute to a breach in the blood-CSF barrier and blood-brain barrier (BBB) that accompanies inflammation and infection. However, as described below, this dichotomy between immune and neuronal candidate genes is an over-simplification.

### PANS candidate genes that primarily function in the peripheral immune system

*PPM1D, CHK2, PLCG2, RAG1,* and *NLRC4* have well-established effects on peripheral innate and adaptive immunity. *PPM1D* codes for a serine/threonine phosphatase that negatively regulates p53 and other members of the DNA repair pathway. Somatic *PPM1D* gain of function mutations act as tumor suppressor genes ^24–26^. The most common are frameshift and nonsense mutations in exons 5 and 6 that lead to the production of a truncated protein that has a stabilizing effect on the retained catalytic portion. Germline exons 5 and 6 truncating mutations are found in children with Jansen de Vries Syndrome (JdVS) ^27–31^. JdVS is characterized by intellectual and developmental disabilities (IDD), restricted eating, high pain threshold, ASD, and psychiatric symptoms (primarily severe anxiety). Thus, there is some overlap in the clinical features observed in PANS and JdVS (e.g., restricted eating, anxiety). However, IDD is nearly universal in JdVS, while the two cases with the S44W variant excelled at school until the development of PANS, which became chronic, leading to persistent neuropsychiatric problems and academic regression. Most children with JdVS have a history of recurrent infections ^27^. However, no consistent immunological findings have been reported. PPM1D regulates T- and B-lymphocyte differentiation, and cytokine production, with *Ppm1d* deficient mice exhibiting a proinflammatory state following an immune challenge, and an increase in macrophage phagocytosis and autophagy ^32–36^. PPM1D has also been found to inhibit intestinal inflammation in inflammatory bowel disease and negatively regulates cytokine production in human neutrophils during sepsis ^37^. The proinflammatory state in *Ppm1d* null mice mimics that seen in PANS patients, suggesting that S44W may induce an exaggerated inflammatory response following an immune challenge through a loss-of-function effect on PPM1D catalytic activity in immune cells. Alternatively, a proinflammatory state could occur through disruption of immune responses caused by other regulatory proteins affected by S44W (see discussion).

Similar to *PPM1D*, *CHK2* also codes for a regulator of the DNA repair response by phosphorylating p53 ^23, 38^. Germline loss of function mutations in the kinase domain are found in several inherited cancer syndromes ^39, 40^. The protein encoded by *CHK2* is expressed in T-cells and regulates IL-2 expression ^41^. The rare variant we identified, T410fsX14, is a truncating mutation in the distal end of the kinase domain and is probably a loss-of-function mutation. Chk2 inactivation in mouse B cells leads to decreased Ig hypermutation and Ig class switching ^42^, conditions that can, along with a DNA repair defect, lead to immune dysregulation ^43–45^. Defective DNA repair could also underlie the effects of the S44W *PPM1D* variant on immune function.

The finding of an ultra-rare variant in *PPM1D* and in one of its principal targets, *CHK2,* strongly supports the idea that these are indeed PANS-causing mutations.

*PLCG2* is primarily expressed in B-cells, and gain-of-function mutations have been found in patients with severe sterile inflammation, recurrent bacterial infections, autoimmune disorders, and humoral immunodeficiency ^46–49^. The *PLCG2* Y206F variant maps to the EF hand domain that binds calcium. Gain of function mutations in *PLCG2* cause autoinflammatory disease through an increase in calcium influx upon B-cell activation ^50^. However, the effect of Y206F on calcium homeostasis awaits experimental validation.

*NLRC4* codes for a component of the inflammasome, a cytosolic multiprotein complex that assembles in response to exogenous or endogenous stressors, thereby playing a major role in autoinflammatory diseases and macrophage activation syndrome ^51–56^. NLRC4 also regulates panoptosis, a recently identified inflammatory programmed cell death pathway, ^57^ and Th17 cells in mice exposed to osmotic stress ^58^.

A total of four patients with ultra-rare variants in *NLRC4* were found in our two cohorts (including patient 17 who also has a *SHANK3* variant). Two variants, c.772T>C (p.C258R) and c.928C>T (p.R310X) map to the NACHT domain, which has intrinsic ATPase activity and facilitates self-oligomerization ^59^. Gain of function mutations in this region lead to severe hyperinflammatory disorders ^55^. Such mutations abrogate autoinhibition by the C-terminal leucine rich region ^60^. p.R310X would eliminate this region from the protein. The other two ultra-rare variants, c.2309T>C (p.M770T) and c.1799A>G (p.E600G), map to the leucine-rich repeat region (LRR). Mutations in the LRR affecting the oligomerization interface were recently described in two patients with early-onset macrophage activation syndrome ^61^.

*RAG1* codes for recombination activating gene 1, which is involved in antibody and T-cell receptor V(D)J recombination. RAG1 deficiency leads to abnormalities in T- and B-cell tolerance and immune dysregulation, leading to both an increased risk of infection and autoimmune problems ^62, 63^. Ultra-rare mutations were found in three patients, one of which, c.256_257delAA causes a frameshift in exon 2 (p.K86VfsX33). This variant has previously been reported in a heterozygous patient with adult-onset lymphopenia, and in a homozygous patient with severe combined immunodeficiency (SCID) ^64^.

### PANS candidate genes that affect microglia function

Several of the PANS candidate genes that affect the peripheral immune system also disrupt microglia function. For example, *Ppm1d* null mice exposed to a hypobaric hypoxic environment showed an augmented release of inflammatory cytokines from activated microglia ^65^. In microglia derived from iPSCs, PLCG2 expression was found to mediate cell survival, phagocytosis, and processing of neuronal debris ^66^. Finally, Nlrc4 regulates apoptotic and pyroptotic microglial death following ischemic stroke ^67, 68^, and astrogliosis and microgliosis in a mouse model of neuroinflammation ^69^.

Additional support for an immune-related effect of a subgroup of PANS-associated candidate genes on microglia function comes from our analysis of a gene expression dataset in LPS-treated mice (GSE102482) ^15^. We found that *Sgce* and *Plcg2* were the 2^nd^ and 8^th^ most downregulated transcripts, and *Nlrc4* the 6^th^ most upregulated transcript, in microglia derived from control and LPS-treated mice upon an analysis of a subset of 148 autism and pediatric immune disorder genes (**Figure 2)**.

**Figure 2.**
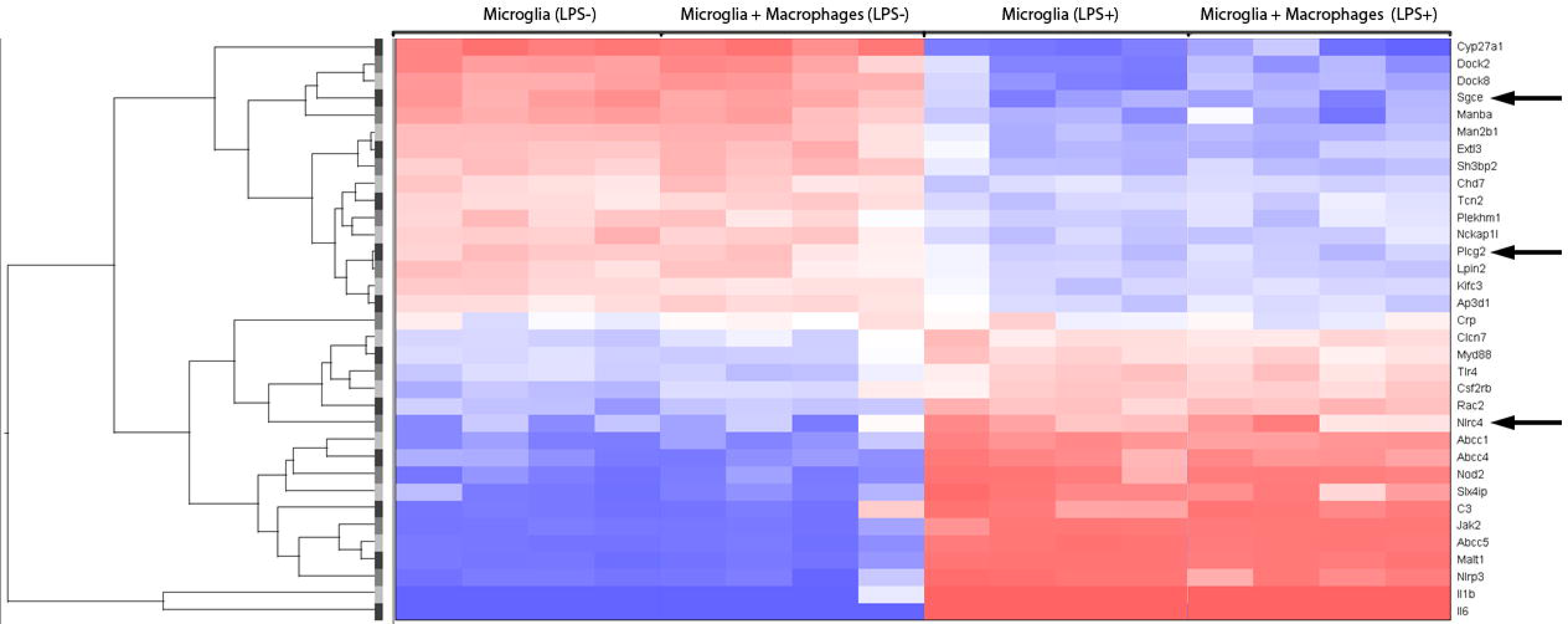
Heat map of LPS-mediated gene expression in microglia. Microarray gene expression data (GSE102482) from Greenhalgh et at. (reference 15) were analyzed to determine the expression pattern of a subset of 148 autism and pediatric immune disorder genes. The heat map shows the data obtained from RNA extracted from microglia grown on their own or co-cultured with peripheral macrophages. Arrows point to differentially expressed genes that are PANS candidates described in this report.

In addition to these PANS candidate genes, the expression of *Sgce*, which will be described in more detail in the next section, is induced in microglia following viral-mediated neuroinflammation in mice ^70^. Similarly, we previously found that *Sgce* expression increased significantly in microglia derived from a mouse model of Rett syndrome, in which innate immune pathways were the most enriched differentially expressed genes ^71^.

These findings support the idea that a subgroup of PANS candidate genes affect microglia function.

### Neuronal PANS candidate genes associated with neurological and neurodevelopmental disorders

Six PANS candidate genes identified in this study, SHANK3, GRIN2A, SYNGAP1, CACNA1B, GABRG2, and SGCE, are primarily expressed in neurons and variants in these genes have been associated with ASD and other neurodevelopmental disorders. *SHANK3,* for example, codes for a scaffold protein that regulates the assembly of PSD-95 at glutamatergic excitatory synapses ^72–75^. It is among the most commonly mutated genes in ASD and other neurodevelopmental disorders ^76–79^. Of the 1003 genes in the Simons Foundation Autism Research Initiative (SFARI) database, *SHANK3* has the second highest number of reports. Ultra-rare variants were found in three PANS patients (c.4622C>T, p.1541L; c.3637dupG, p.1213fsX68; c.4776dupC, p.T1593fsX86). Although *SHANK3* mutations in ASD are scattered throughout the gene, the majority, including our cases, are found in exon 21, which codes for the proline-rich region (PRR) that binds to and draws other synaptic proteins to the post-synaptic density. ASD-associated variants in *SHANK3* disturb mGluR-dependent synaptic plasticity ^73^.

Case 18 (c.3637dupG, p.1213fsX68) was diagnosed with Phelan-McDermid syndrome, which is usually caused by *SHANK3* deletions, developed the sudden onset of severe OCD following bacterial infections. Case 19 (c.4776dupC, p.T1593fsX86) has a history ASD with minimal verbal ability since early childhood. She developed the sudden onset of OCD and aggression as a child that improved dramatically with IVIG, although the baseline ASD and poor verbal ability did not improve.

Our finding of co-morbid ASD and PANS in the SHANK3 cases is similar to observations made by Bey et al., who identified *de novo SHANK3* variants in four girls with lifelong, stable developmental delay (DD) who developed subacute, severe psychiatric symptoms resembling PANS, that responded, with varying degrees of success, to immunotherapy ^80^. Similarly, in an analysis of 38 individuals with Phelan-McDermid Syndrome, acute regression triggered by infections, changes in hormonal status, and stressful life events were commonly observed, several of whom improved with IVIG ^81^.

These findings show that a subgroup of patients with *SHANK3*-associated ASD and developmental disorders have an underlying susceptibility to develop acute onset neuropsychiatric problems that have a neuroinflammatory component.

Four other PANS-associated neuronal candidate genes we identified, *SYNGAP1, GRIN2A, GABRG2,* and *CACNA1B* mutations have also been found in ASD, epilepsy, and IDD ^77, 82–86^. GRIN2A and SYNGAP1 bind to PSD-95. According to the SFARI database, *SYNGAP1* has the ninth highest number of reports. It codes for a postsynaptic Ras GTPase activating protein that regulates developing excitatory synaptic structure and function ^87^. Case 8 has a nonsense mutation at *SYNGAP1* codon 1179. Like the *SHANK3* cases, she was diagnosed with ASD as a child, and developed acute onset OCD and food restrictions following a culture positive streptococcus pharyngitis.

*GRIN2A* codes for the glutamate ionotropic receptor NMDA type subunit 2A. Mutations have been found in a variety of neurodevelopmental disorders including ASD, epilepsy, and developmental disabilities ^84, 88^. The ultra-rare variant found in case 6 maps to a conserved region of the CTD domain, which is involved in mediating intracellular signaling ^85^. The CTD region interacts with the MAGUK (membrane-associated guanylate kinase) family of proteins that bind to PSD-95 and function as important modulators of synaptic plasticity ^89^. This subject has a history of dyslexia, ADHD, and emotional problems.

*CACNA1B* codes for the voltage-dependent N-type calcium channel subunit alpha-1B, a constituent of the Cav2.2 channel. It too is a regulator of synaptic function that acts at the presynaptic terminal to increase neurotransmitter release, which then influences postsynaptic dendritic spine function ^90^. In addition to having frequent infections and PANS, Case 3 was diagnosed with Hodgkin’s lymphoma, nodular sclerosis subtype. He was treated with ABVD, and subsequently, autologous hematological stem cell transplant (HSCT). It is interesting to note that Hodgkin’s lymphoma is often found in patients with autoimmune problems and is associated with abnormalities in IL-13 signaling, a cytokine produced by mast cells, eosinophils, nuocytes, and Th2 cells ^92–94^. This patient also has psoriasis, a Th17 associated autoimmune disorder ^95, 96^. This suggests the possibility that *CACNA1B* could have unrecognized effects on the immune system by disrupting IL-13 and Th17 signaling leading to PANS, psoriasis, and Hodgkin lymphoma. It is also interesting to note that his neuropsychiatric symptoms have temporarily disappeared following HSCT. However, the follow-up period has only been a few months, so it is too early to determine if the treatment has had a positive impact on his neuropsychiatric symptoms. Autologous HSCT is an emerging technology for treating severe autoimmune disorders ^91^. Given these observations, HSCT could prove to be an option for treating a small subgroup of chronic, refractory, severely impaired PANS patients.

*GABRG2* codes for the gamma subunit of the GABA-A receptor, a ligand-gated chloride channel. It is one of the most mutated genes in febrile seizures and other forms of epilepsy ^97, 98^. We found a nonsense mutation, c.649C>T; p.Gln217X, in a child with severe PANS (case 5). This is the same mutation previously found in sleep-related epilepsy ^99^. Case 5 has a history of febrile seizures.

*SGCE* is a member of the sarcoglycan family of transmembrane proteins, a component of the dystrophin-associated glycoprotein complex (DGC). Studies carried out in knockout mice show that SGCE inhibits excitatory synapse formation, and that loss-of-function mutations lead to an increase in excitatory synapses ^100^. The DGC is involved in organizing aquaporin-4-containing protein complexes in glia that controls the formation of the glymphatic system. The glymphatic system clears waste from the brain, and impaired glymphatic drainage has been found in neuroinflammation and dementia ^101–103^. Ultra-rare mutations in *SGCE* were found in two unrelated subjects, cases 10 and 11.

Remarkably, *SGCE, CACNA1B,* and *GRIN2A* are well-known for causing Myoclonus-Dystonia (M-D), a hyperkinetic movement disorder, which is similar to tics, a diagnostic feature of PANDAS. *SGCE,* in fact, is the most commonly mutated gene in M-D, with more than one-third of cases having truncating mutations in exon 3, which contains the del150Iso in-frame mutation found in case 10 ^104–106^. Interestingly, patients with myoclonus caused by *SGCE* mutations frequently have co-morbid psychiatric symptoms, including depression, anxiety, bipolar disorder, phobias, alcoholism, and OCD ^106^, symptoms that overlap with PANS. Similarly, *GRIN2A* and *CACNA1B* mutations have been found in dystonic and other rare movement disorders ^85, 107–110^.

These findings suggest that deleterious variants in these “neuronal” genes can result in a range of clinical phenotypes; M-D, ASD, developmental disabilities, epilepsy, and PANS, or PANS co-morbid with these neurodevelopmental problems. It is important to add, however, that case 7 with the *PLCG2* ultra-rare variant c.617A>G, p.Y206F, as well as cases 12, 14, and 15, who have mutations in *NLRC4* and *RAG1*, also had preexisting ASD prior to developing PANS. Thus, PANS/ASD co-morbidity is not restricted to cases with variants in genes typically associated with neurodevelopmental disorders. Other genetic factors causing the neurodevelopmental problems, however, cannot be ruled out at this time.

### Connectivity Network

To assess potential functional connections between the PANS candidate genes, we generated a connectivity network to show the direct (solid lines) and indirect interactions (dotted lines) (**Figure 3**). Included in the network are two additional PANS candidate genes, *MTHC2* and *BID,* identified by one of us (R.T.) by GWAS that will be described in a separate paper. Central to the network is the NF-κB complex transcriptional regulator, which is activated by a variety of immune, infectious, and non-immune (e.g., oxidative stress; toxins) stressors ^111, 112^. Inappropriate activation of NF-κB has been associated with inflammatory diseases ^113^. Directly connected to the NF-κB hub are the PANS candidate genes; *PPM1D*, *PLCG2, NLRC4, RAG1,* and *BID*, along with *CHK2* and *MTCH2*, which bind to PPM1D and BID, respectively. This hub represents the set of PANS genes that likely to function through a disruption of peripheral and central innate immunity. The PANS candidate genes not directly connected to NF-κB expression are those that are primarily expressed in the brain and cause neurodevelopmental disorders (*CACNA1B, SYNGAP1, GRIN2A, SGCE, GABRG2,* and *SHANK3*).

**Figure 3.**
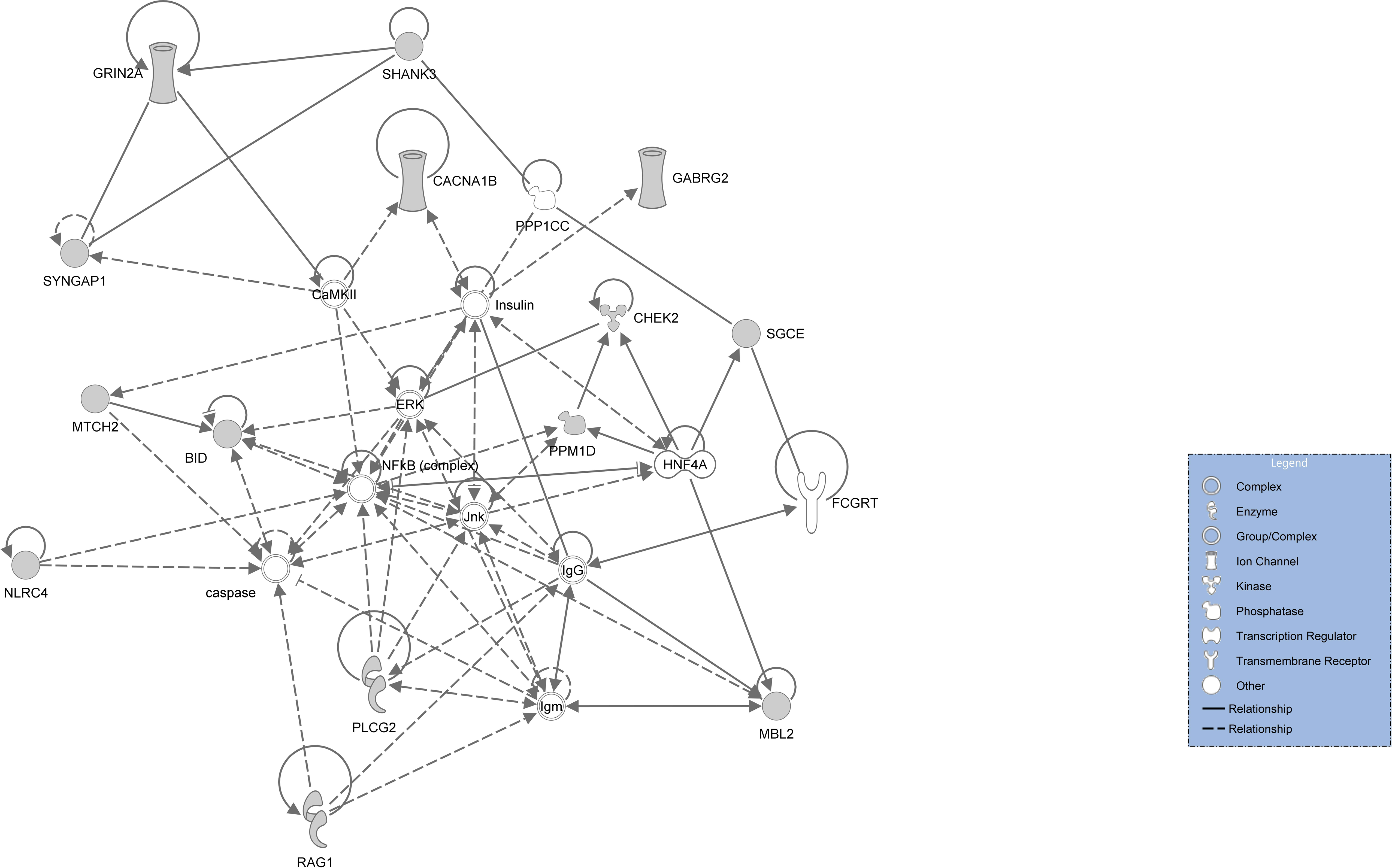
Connectivity network. A connectivity network was generated for each candidate gene using IPA software. Two additional PANS candidate genes, MTHC2 and BID, identified by one of us (R.T.) are included. These will be described in a separate paper. Central to the network is the NF-κB complex transcriptional regulator hub, which connects to PPM1D, PLCG2, NLRC4, RAG1, BID, CHK2 and MTCH2. The PANS candidate genes not directly connected to NF-κB expression are those that are primarily expressed in the brain and cause neurodevelopmental disorders (CACNA1B, SYNGAP1, GRIN2A, SGCE, GABRG2, and SHANK3). See Supplemental Data for symbol key.

Interestingly, CaMKII, a biomarker from the Cunningham panel, is also centrally localized within the connectivity network and has a close relation to multiple candidate genes.

### Tissue and Cell Expression Pattern of PANS Candidate Genes

To gain further insight into the mechanisms by which the PANS candidate genes lead to neuroinflammation, we examined their expression pattern using several gene expression resources, one of which is a single cell RNA-seq (scRNA-seq) database of white blood cells (WBCs)^16^. This database contains gene expression patterns in control samples (N=6) and hospitalized COVID-19 patients (N=7). The candidate genes that have established effects on peripheral immune function, *PPM1D, CHK2,* and *RAG1,* are expressed in multiple cell types, *PLCG2* is primarily expressed in B-cells, and *NLRC4* in monocytes and neutrophils (**Figure S1**). Interestingly, the expression of several genes was significantly affected in the COVID-19 patients. Most notably, a >100-fold increase in *NLRC4* expression was found in developing neutrophils. This suggests that PANS-associated *NLRC4* mutations could lead to a marked alteration in inflammasome function following infections. In addition, a >8-fold increase in *CHK2* (also known as CHEK2) was found in interferon stimulated genes in T4 cells.

Although most of the “neuronal” PANS candidate genes are expressed at very low levels in WBCs, there are three notable exceptions. Most striking is *GABRG2,* which shows marginal expression in all WBCs except for T.gd cells (γδ T cells) in COVID-19 patients, where a >20-fold increase in expression was found compared with controls. γδ T cells form a minor population of WBCs, but they increase dramatically during infections and play a key role in autoimmunity and immune surveillance ^114, 115^. Perhaps more importantly from the perspective of PANS is the finding that γδ T cells are found in meninges where they secrete the proinflammatory cytokine IL-17a and participate in the development of anxiety-like behavior in mice ^116^.

*SYNGAP1* is expressed in many WBC subtypes, especially T cells. Strikingly, like GABRG2, expression in γδ T cells increases nearly 4-fold in COVID-19 patients. And *SHANK3* expression, while negligible in most WBCs, is abundant in a cluster consisting of two groups of cells labeled as stem cells and eosinophils.

These finding suggest that some neuronal PANS candidate genes that typically cause neurodevelopmental disorders have unanticipated effects on immune function and their expression is extremely responsive to an infectious disease stressor. Parenthetically, these findings show that it will be important to monitor the potential impact of SARS-CoV-2 as an infectious disease inducer of PANS.

We next analyzed the cell and tissue expression pattern of each of the PANS candidate genes in normal human tissues using the GTEx dataset (**Figure S2**). The gene expression patterns, based on bulk RNA-seq, generally conform to the functional groups described above and the connectivity network. The genes affecting peripheral immunity that are connected to the NF-κB hub are expressed primarily in blood and EB-transformed lymphoblasts, and less so in the brain, while the genes associated with neurodevelopmental disorders show the opposite pattern. *PPM1D, RAG1,* and *SGCE* are highly expressed in both.

To gain further insight into the expression of the PANS candidate genes in the nervous system, we examined the cell type expression pattern in adolescent and fetal mouse brains. One of the more striking observations in the adolescent brain is the relatively high level of *Chk2* expression in ependymal cells compared with all other brain cell types (**Figure 4; see Table S1 for abbreviation key**). Ependymal cells line the ventricles and spinal canal and play an important role in the production of CSF ^117^. As part of the choroid plexus, ependymal cells regulate the blood-CSF barrier ^118^. The choroid plexus plays a key role in neuroinflammatory and neurodegenerative disorders and serves as a nexus for peripheral T-cells that can infiltrate the brain, as well as access, when damaged, to proinflammatory cytokines, complement, and autoantibodies from the circulation to the brain parenchyma ^119–121^. *Ppm1d, Sgce, Plcg2, Syngap1,* and *Shank3* are also expressed in these cells, suggesting that their mutated versions could disturb the blood-CSF barrier.

**Figure 4.**
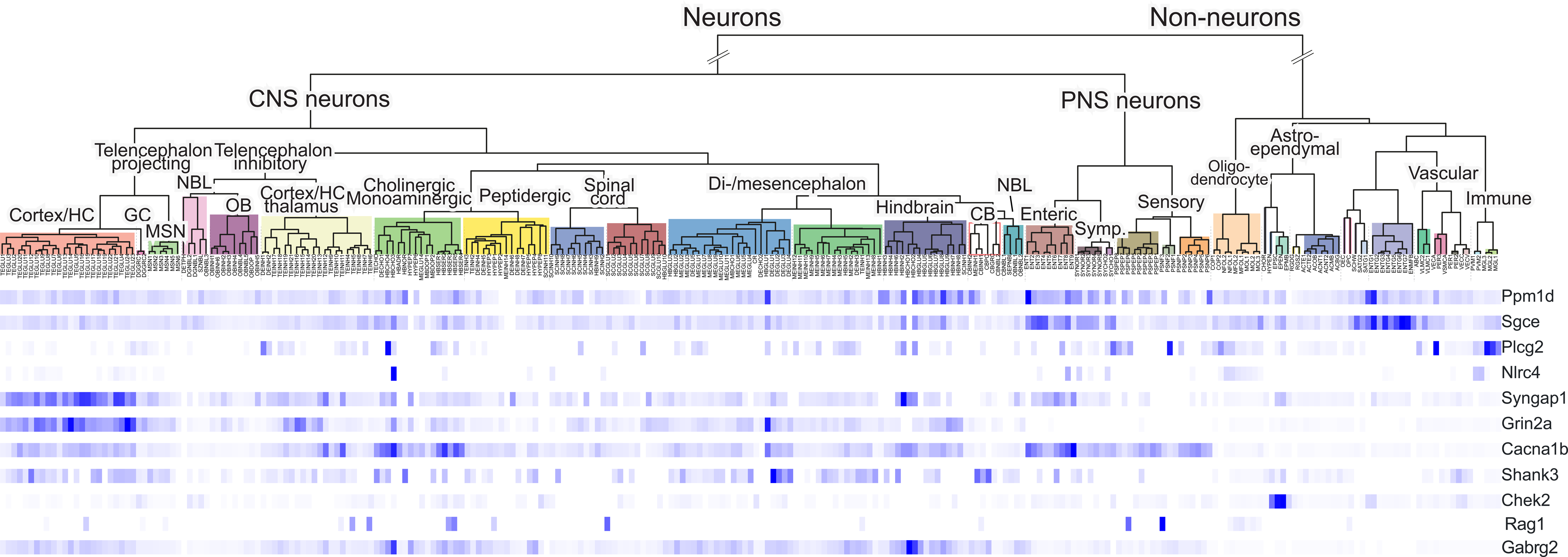
Single cell RNA-seq expression pattern of each PANS candidate gene in mouse adolescent brains (mouse brain atlas; see Web Resources)

Another interesting finding in adolescent nervous system is the relatively high level of expression of *Ppm1d, Syngap1, Cacna1b,* and *Sgce* throughout the enteric nervous system. The potential implications of this expression pattern will be described in the discussion section.

We also examined the single cell gene expression pattern of the PANS candidate genes in the developing mouse brain, considering the fact that several cause neurodevelopmental disorders **(Figure S3)**. One of the more interesting findings is the expression of *Shank3,* which, as expected, was diffusely expressed throughout the brain in both glutamatergic neurons and GABAergic interneurons. However, the highest levels were seen in the choroid plexus and cerebral vasculature. In fact, all of the PANS candidate genes, with the exception of *RAG1* are expressed in fetal choroid plexus, cerebral vasculature, and pericytes, with *Sgce, Chk2* and *Ppm1d,* along with *Shank3,* being most prominent. This is similar to the single cell expression pattern seen in adolescent brains (**Figure 4**), reinforcing the idea that disruption of the blood-CSF barrier and/or the BBB is playing a role in the emergence of PANS.

## Discussion

Genetic contributions to disease can be divided into common variants that have relatively small biological effects, which are generally identified by large-scale GWAS, and high impact, biologically significant ultra-rare variants. So far, no reports have been published in which common variants were identified, although there is a small, unpublished study derived from a direct-to-consumer DNA testing service in which five SNPs associated with PANS and PANDAS were found (see Web Resources), two of which occur in credible candidate genes (rs5029936 in *TNFAIP3*; rs76298532 in *MCPH1*).

In the current study, we searched for ultra-rare variants in PANS patients using next generation sequencing. For the European cohort, cases were selected for their severity. The index case was a teenage girl with severe, chronic PANS who was found, along with her affected brother, to have an ultra-rare *PPM1D* mutation (c.131C>G; p.S44W) (**Figure 1**). This mutation is upstream of the phosphatase domain and its effect on PPM1D catalytic function is not immediately clear. However, the nearby serine amino acids S40 and S46 could be playing a role the development of PANS in this family. These are substrates for the serine kinases DYRK1A and HIPK2, which cause IDD, ASD, and abnormal feeding behavior when mutated ^122–124^. DYRK1A also has important effects on innate immunity by regulating the balance between Th17 and T regulatory (Treg) cells, T cell subsets involved in the development of autoimmune diseases and inflammation ^125–129^. Interestingly, Th17 mediates the effects of *Streptococcus* on neuroinflammation in a mouse model of PANDAS by causing a breach in the BBB, with subsequent brain infiltration of Th17 lymphocytes ^125, 130–132^. HIPK2 regulates PPM1D protein levels by inducing its degradation in DNA-damaged cells, triggering activation of the DNA repair pathway ^133^. HIPK2 expression increases in LPS-stimulated macrophages, and its knockdown attenuates the expression of inflammatory cytokines ^134, 135^. S46 is also a HIPK2 substrate. Thus, an effect of S44W on immune regulation could be secondary to altered interactions between DYRK1A and HIPK2 with PPM1D.

Following the discovery of this variant, we identified an additional eight other severely affected patients through a PANS/PANDAS advocacy group that primarily operates in the European Union. WGS was carried out, which resulted in the identification of ultra-rare variants in plausible candidate genes in every patient in the European cohort. Although there is a clear ascertainment bias for these participants, the findings of ultra-rare variants in the same genes and other plausible candidates in an unselected, modestly sized population in the United States cohort suggest that severe PANS is often caused by biologically powerful ultra-rare genetic variants. The condition can be viewed as a heterogenous genetically dominant disorder, albeit, with reduced penetrance. Evidence that penetrance is not 100% is seen in several families in whom transmission occurred from a carrier parent who do not have neuropsychiatric problems typical of PANS. The *PPM1D* variant, for example, was inherited from an asymptomatic father who has a family history of autoimmune disorders, In addition, two siblings of case 10 (*SGCE* (del150 Iso; c.450_452) also inherited the variant. One has asthma, allergies, and contact dermatitis, but no neuropsychiatric problems; The other has similar atopic problems. Although he does not have PANS, he has developed debilitating, recurrent cluster-type headaches. Whether the headaches are a manifestation of neuroinflammation, or vasculitis is not known.

Finally, case 9 (NLRC4 c.772T>C; p.C258R), inherited the mutation from a mother who has with a history of psoriasis and arthritis. She was diagnosed with focal epilepsy as an adult but has no history of PANS. A younger sibling with the same variant has a history suggestive of PANS but has not yet been clinically ascertained.

The lack of complete penetrance is consistent with a small monozygotic twin study in PANDAS that showed a range of different phenotypes, including complete discordance ^136^. Similarly, the MZ twin concordance rate in autoimmune disorders ranges from approximately 20%-70% depending on the condition ^137^.

This suggests that environmental factors are playing a role in PANS. In the case of the generation divide between carrier parents and their affected children, changes in the prevalence of certain strains of bacteria and antigenic shifts in viruses implicated as PANS triggers, could account for their different clinical outcomes. *Borrelia burgdorferi,* which causes Lyme disease, is an example of an infectious disease that has markedly increased in prevalence in the past 30 years, while antigenic shifts in viruses that cause influenza or common cold are examples of infectious diseases that are subject to rapid genetic changes. In addition, the current generation of youth might be exposed to non-infectious environmental triggers that were not as prevalent in past generations. Although unaffected carrier parents might be exposed to the same infectious agents or non-infectious stressors that are currently triggering PANS flareups in their children, age-related differences in the blood-CSF barrier or BBB could make the adult brain less susceptible ^138^. Genetic background and stochastic T cell receptor and IgG gene rearrangement could also explain reduced penetrance in families, and discordance in MZ twins.

The candidate genes identified in this study can be broadly separated into those that have established effects on peripheral innate and adaptive immunity, and those that affect synaptic function in particular the PSD-95 complex. However, this peripheral vs central dichotomy, which could potentially explain the presence or absence of immune markers and response to immunomodulators, is an oversimplification. First, as noted earlier, several PANS candidate genes that have effects on peripheral innate immunity, such as *PPM1D, PLCG2,* and *NLRC4,* also affect microglia function and are differentially expressed in those cells following an immune challenge. Second, patients with mutations in genes that function as synaptic regulators, such as *SHANK3*, can respond to IVIG, as shown by Bey et al ^80^. Similarly, our cases with ultra-rare variants in *SHANK3*, *CACNA1B, SGCE* and *GABRG2* also responded to IVIG. Third, as shown in **Figure S1**, neuronal genes like *SHANK3, GABRG2* and *SYNGAP1* show expression patterns that strongly support an adverse effect on immune function, in particular γδ T-cells, based on their markedly altered expression during upon an infectious disease challenge. Fourth, some of the neuronal PANS candidate genes that have effects on synaptic function, such as *SHANK3, SGCE, CHK2,* and *PPM1D*, are also expressed in the choroid plexus and brain vascular endothelium, which could potentially connect peripheral inflammation with neuroinflammation through disruption of the brain/CSF barrier and BBB caused by mutations in these genes. SHANK3 is a cytoskeletal protein that regulates glutamatergic synaptogenesis, but it can also function as a scaffolding protein in epithelial cells^139^. In addition, PPM1D has been found to regulate BBB function and neuroinflammation in a co-culture of human brain-microvascular endothelial cells and human astrocytes treated with LPS ^140^.

Expression of PANS candidate genes in the choroid plexus and vascular endothelium during fetal development is also interesting when considering the phenomenon known as maternal immune activation (MIA), a proinflammatory state in pregnancy triggered by infection, maternal autoimmune disorders, and non-infectious peripheral inflammation^141, 142^. The fetal brain is vulnerable to changes in the maternal/fetal environment, such as MIA ^141–143^, which has been shown in animal models to adversely affect brain development, leading to behaviors and learning difficulties similar to those seen in patients with ASD and schizophrenia ^144–152^. Consequently, it is conceivable that the pathophysiological process that leads to the development of PANS and co-morbid neurodevelopmental disorders could begin during fetal life in some patients and genetic subgroups.

An effect on the choroid plexus and vasculature endothelium (and γδ T cells) could also explain the response to IVIG in those patients harboring mutations in the “neuronal” subgroup of candidate genes. IVIG can reduce neuroinflammation by affecting T-cell/microglia crosstalk, reducing levels of proinflammatory cytokines, blocking Fcγ receptors, inhibiting complement, and repairing disrupted brain-CSF barriers ^153–156^.

While extra-neuronal effects of synaptic proteins could have an impact on the development of PANS, their effects on synaptogenesis in the development of PANS should also be considered. One possible mechanism is a defect in PSD-95 assembly and glutamate receptor function leading to abnormal synaptic remodeling. This could trigger over-activation of microglia, which in turn would release proinflammatory cytokines, complement, prostaglandins, proteinases, and reactive oxygen species, causing neuroinflammation. An extension of this model is that a similar overactivation of microglia could be driven by a primary, intrinsic defect caused by PANS candidate genes that are highly expressed in microglia, but not neurons (e.g., *PLCG2, NLRC4*). This is consistent with our finding that *Plcg2, Sgce,* and *Nlrc4* are differentially expressed in LPS-treated microglia (**Figure 2)**. Non-neuronal stimuli that might induce microglia overactivation in PANS is open to speculation, but could include oxidative stress, environmental toxins, and peripheral infection/inflammation.

Finally, it is interesting to consider the implications of the finding that several PANS candidate genes are also mutated in D-M, ASD, and other neurodevelopmental disorders. PANS is occurring as an independent phenotype in some of our cases, and as a comorbid trait in others. These comorbid cases are like the *SHANK3-*mutated patients reported by Bey et al. ^80^. Similarly, Jones et al., described eight children with ASD and other neurodevelopmental disorders, with a strong family history of maternal autoimmune thyroid disorders, who presented with infection-induced, abrupt onset of neuropsychiatric symptoms, primarily OCD and tics, along with autistic or global regression^157^. Overall, comorbid ASD and other neuropsychiatric disorders were found in nine cases in our study, not including cases 1 and 2 who were typically developing prior to the onset of PANS and the development of chronic symptoms (**Table 2**)

The development of either PANS, a neurodevelopmental disorder, or PANS comorbid with a neurodevelopmental disorder, could be due to subtle mutation-specific effects on neuronal or immune function, environmental factors (e.g., MIA), or genetic background. A genetic connection between PANS, D-M, ASD, and other neurodevelopmental disorders also supports the idea that an inflammatory component is involved in the pathogenesis of subgroups of patients with schizophrenia and ASD, as suggested by numerous genetic and molecular studies ^157–164^ At the very least, an inflammatory component leading to the abrupt onset of PANS superimposed on a chronic neurodevelopmental disorder is a real phenomenon that needs to be recognized by the medical community because immunological therapies like IVIG could be very helpful.

### Expression of PANS/PANDAS candidate genes in the enteric nervous system

Another aspect of our genetic analysis was the finding that many of the candidate genes we identified are expressed in enteric neurons, which could perhaps explain the abdominal complaints experienced by PANS patients ^3^. In addition, enteric expression could be playing a direct role in the development of PANS in some genetic subgroups. For example, cholinergic fibers come in close contact with macrophages, plasma cells, and lymphocytes located in the intestinal mucosa, and gut-associated lymphoid tissue ^165^. In addition, patients with PANS/PANDAS have been found to have differences in the gut microbiome compared with controls ^166, 167^, and an increase in gut-derived LPS and markers of oxidative stress ^168^. More specific to the candidate genes we identified, several have established effects on the gut. PPM1D, for example, has been found to have a protective effect on oxidation stress-induced gut permeability ^165^. In addition, *Shank3* knockout mice have an altered microbiota composition, an increase in LPS levels in the liver, and altered gut permeability ^169, 170^. Furthermore, variants in and altered gut permeability *NLRC4* have been implicated in inflammatory bowel disease ^171, 172^.

Overall, the expression of PANS candidate genes in the enteric nervous system fits into the emerging idea that disruption of the gut-brain connection and the gut microbiome are involved in the pathogenesis of ASD, neurodegeneration, neuropsychiatric disorders, autoimmune disorders, and PANS ^173–177^.

In summary, we identified ultra-rare genetic variants in PANS patients that appear to function at multiple levels of the neuroinflammatory circuit, including peripheral and central innate immunity, synaptogenesis, the blood-CSF barrier, and perhaps the enteric nervous system. Dissecting the molecular and cellular pathogenesis of the PANS candidate variants will require an analysis in mouse models, as well as patient-specific induced pluripotent stem cells, from which cells of importance in the development of PANS, such as neurons, microglia, astrocytes, neuronal epithelium, vascular endothelium, and gut organoids can be derived.

## Data Availability

genotyping data are available from the co-first authors and the senior author

## Supplemental Information

**Supplemental data include 3 figures and 1 table, Supplemental Note: Case Reports**

## Acknowledgments

The authors want to thank EXPAND for directing researchers to PANS cases throughout the European Union, and Erika Pedrosa for copy editing the manuscript. HML is supported by the National Institute of Child Health and Human Development NIH/NICHD; P30 HD071593 to the Albert Einstein College of Medicine’s Rose F. Kennedy Intellectual and Developmental Disabilities Research Center. The Lachman lab also receives support from the Janice C. Blanchard Family Fund. P.J. van der Spek is supported by EU H2020 grants, an ImmunAID grant (ID: 7792950), and a MOODSTRATIFICATION grant (ID: 754740). The Bioinformatics infrastructure and team is supported by grants from KWF, NWO/ZonMW and the Dutch Heart foundation through the BDVA initiated H2020 Bigmedilytics program on Personalized Medicine. Neither the authors nor their academic institutions received payment or services from a third party for any aspect of the submitted work including the study design and manuscript preparation.

The authors dedicate this paper to Dr. Paul Janssen whose ideas about the role of the immune system in tics and psychosis help form the framework for this research project, and to participating families for their courage and support.

## Web Resources

https://www.nimh.nih.gov/health/publications/pandas/index.shtml

https://www.expand.care

http://rstats.immgen.org/Skyline_COVID-19/skyline.html

https://www.qiagenbioinformatics.com/products/ingenuitypathway-analysis

https://www.omim.org

https://www.pandaspansinstitute.com

https://gene.sfari.org/database/human-gene

https://www.gtexportal.org/home

https://genome.ucsc.edu

http://mousebrain.org

https://www.phosphosite.org//homeAction.action

https://osf.io/q4y2k/

