## Supplementary material for "Identification of ultra-rare genetic variants in Pediatric Acute Onset Neuropsychiatric Syndrome (PANS) by exome and whole genome sequencing": supp fig 1

### Figure S1:  cell RNA-seq (scRNA-seq) peripheral white blood cells

http://rstats.immgen.org/Skyline_COVID-19/skyline.html

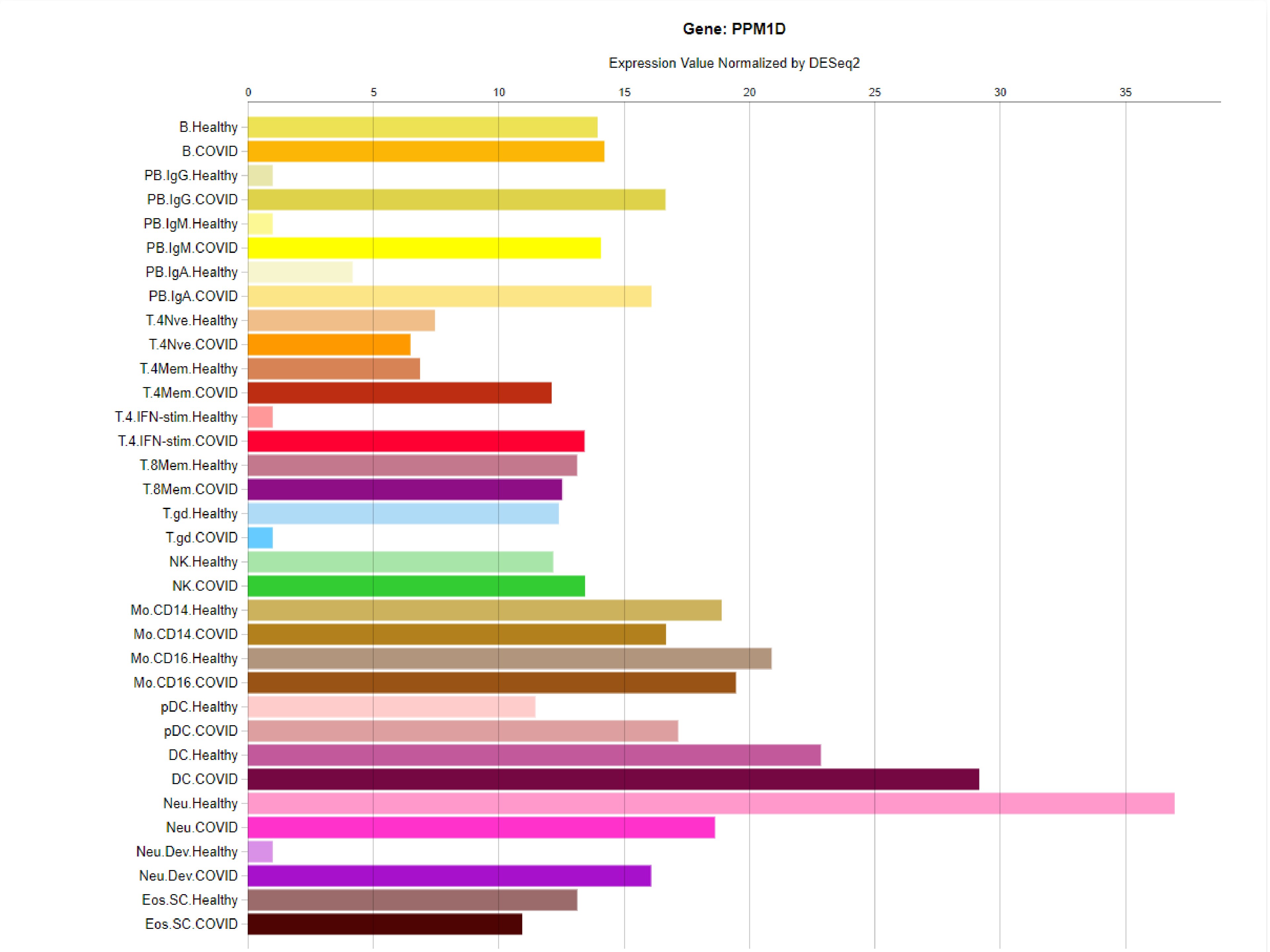
Gene: PPM1D
Expression Value Normalized by DESeq2
0
5
10
15
20
25
30
35
B.Healthy
B.COVID
PB.IgG.Healthy
PB.IgG.COVID
PB.IgM.Healthy
PB.IgM.COVID
PB.IgA.Healthy
PB.IgA.COVID
T.4Nve.Healthy
T.4Nve.COVID
T.4Mem.Healthy
T.4Mem.COVID
T.4.IFN-stim.Healthy
T.4.IFN-stim.COVID
T.8Mem.Healthy
T.8Mem.COVID
T.gd.Healthy
T.gd.COVID
NK.Healthy
NK.COVID
Mo.CD14.Healthy
Mo.CD14.COVID
Mo.CD16.Healthy
Mo.CD16.COVID
pDC.Healthy
pDC.COVID
DC.Healthy
DC.COVID
Neu.Healthy
Neu.COVID
Neu.Dev.Healthy
Neu.Dev.COVID
Eos.SC.Healthy
Eos.SC.COVID

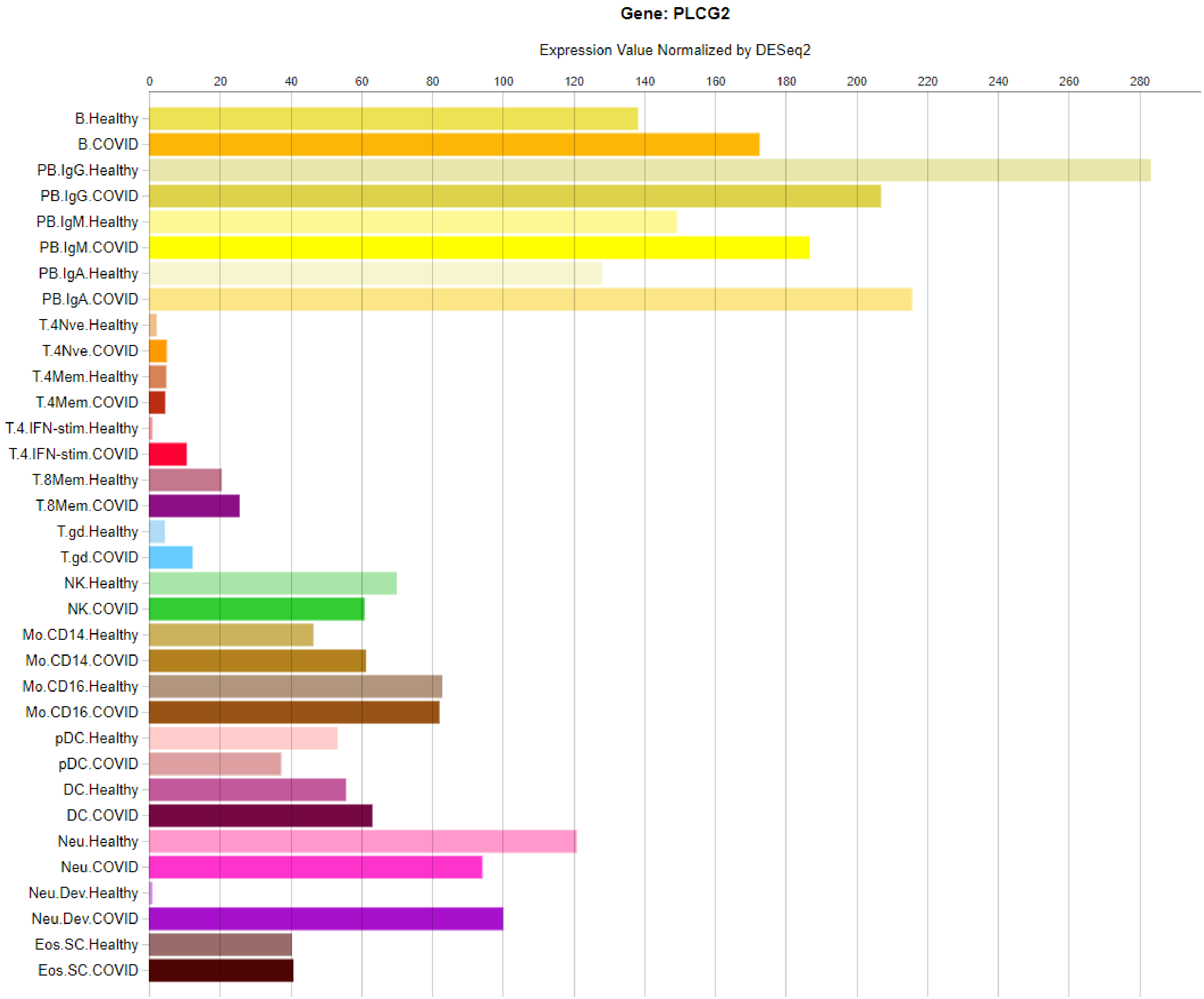
Gene: PLCG2
Expression Value Normalized by DESeq2
0
20
40
60
80
100
120
140
160
180
200
220
240
260
280
B.Healthy
B.COVID
PB.IgG.Healthy
PB.IgG.COVID
PB.IgM.Healthy
PB.IgM.COVID
PB.IgA.Healthy
PB.IgA.COVID
T.4Nve.Healthy
T.4Nve.COVID
T.4Mem.Healthy
T.4Mem.COVID
T.4.IFN-stim.Healthy
T.4.IFN-stim.COVID
T.8Mem.Healthy
T.8Mem.COVID
T.gd.Healthy
T.gd.COVID
NK.Healthy
NK.COVID
Mo.CD14.Healthy
Mo.CD14.COVID
Mo.CD16.Healthy
Mo.CD16.COVID
pDC.Healthy
pDC.COVID
DC.Healthy
DC.COVID
Neu.Healthy
Neu.COVID
Neu.Dev.Healthy
Neu.Dev.COVID
Eos.SC.Healthy
Eos.SC.COVID

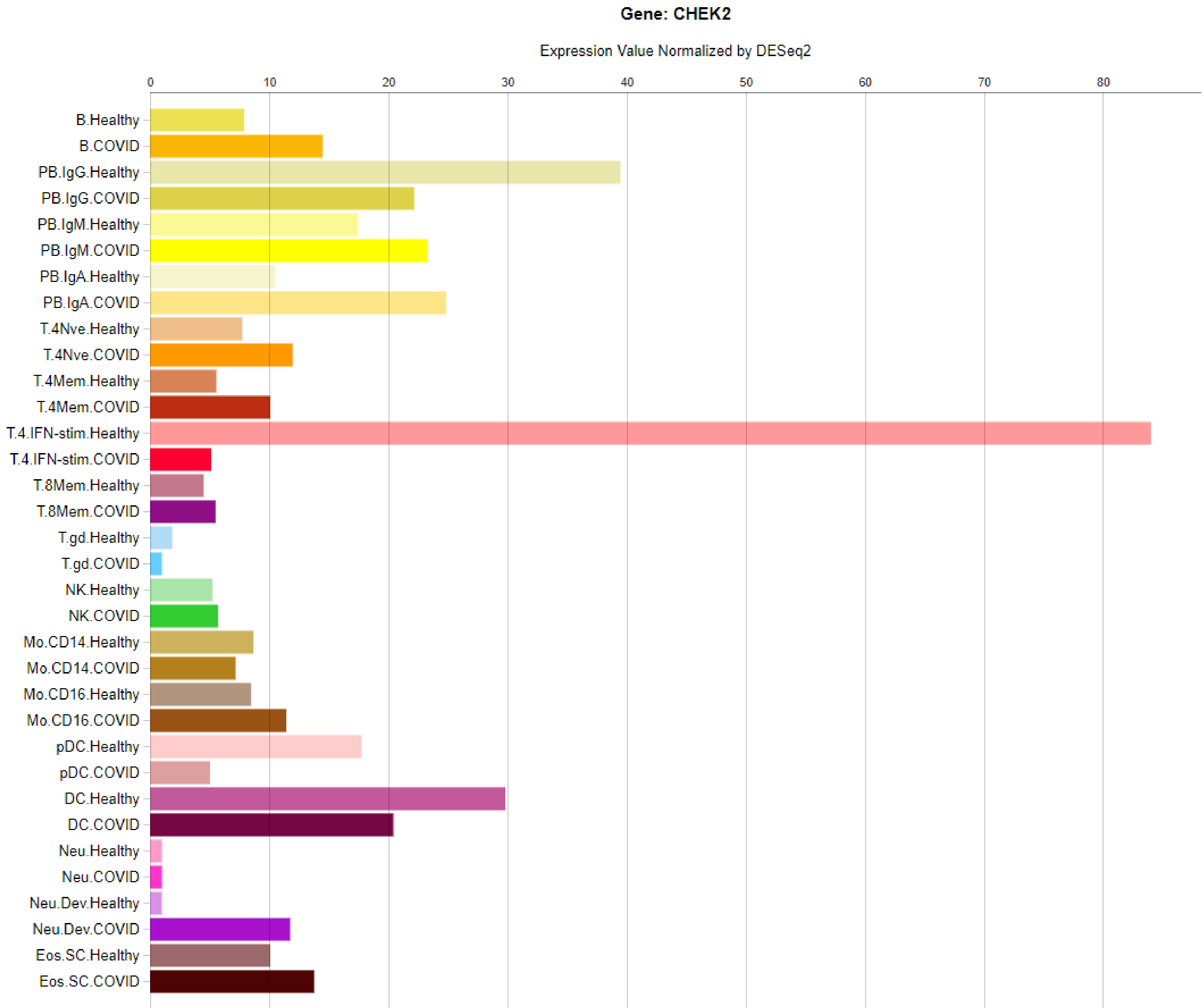
Gene: CHEK2
Expression Value Normalized by DESeq2
0
10
20
30
40
50
60
70
80
B.Healthy
B.COVID
PB.IgG.Healthy
PB.IgG.COVID
PB.IgM.Healthy
PB.IgM.COVID
PB.IgA.Healthy
PB.IgA.COVID
T.4Nve.Healthy
T.4Nve.COVID
T.4Mem.Healthy
T.4Mem.COVID
T.4.IFN-stim.Healthy
T.4.IFN-stim.COVID
T.8Mem.Healthy
T.8Mem.COVID
T.gd.Healthy
T.gd.COVID
NK.Healthy
NK.COVID
Mo.CD14.Healthy
Mo.CD14.COVID
Mo.CD16.Healthy
Mo.CD16.COVID
pDC.Healthy
pDC.COVID
DC.Healthy
DC.COVID
Neu.Healthy
Neu.COVID
Neu.Dev.Healthy
Neu.Dev.COVID
Eos.SC.Healthy
Eos.SC.COVID

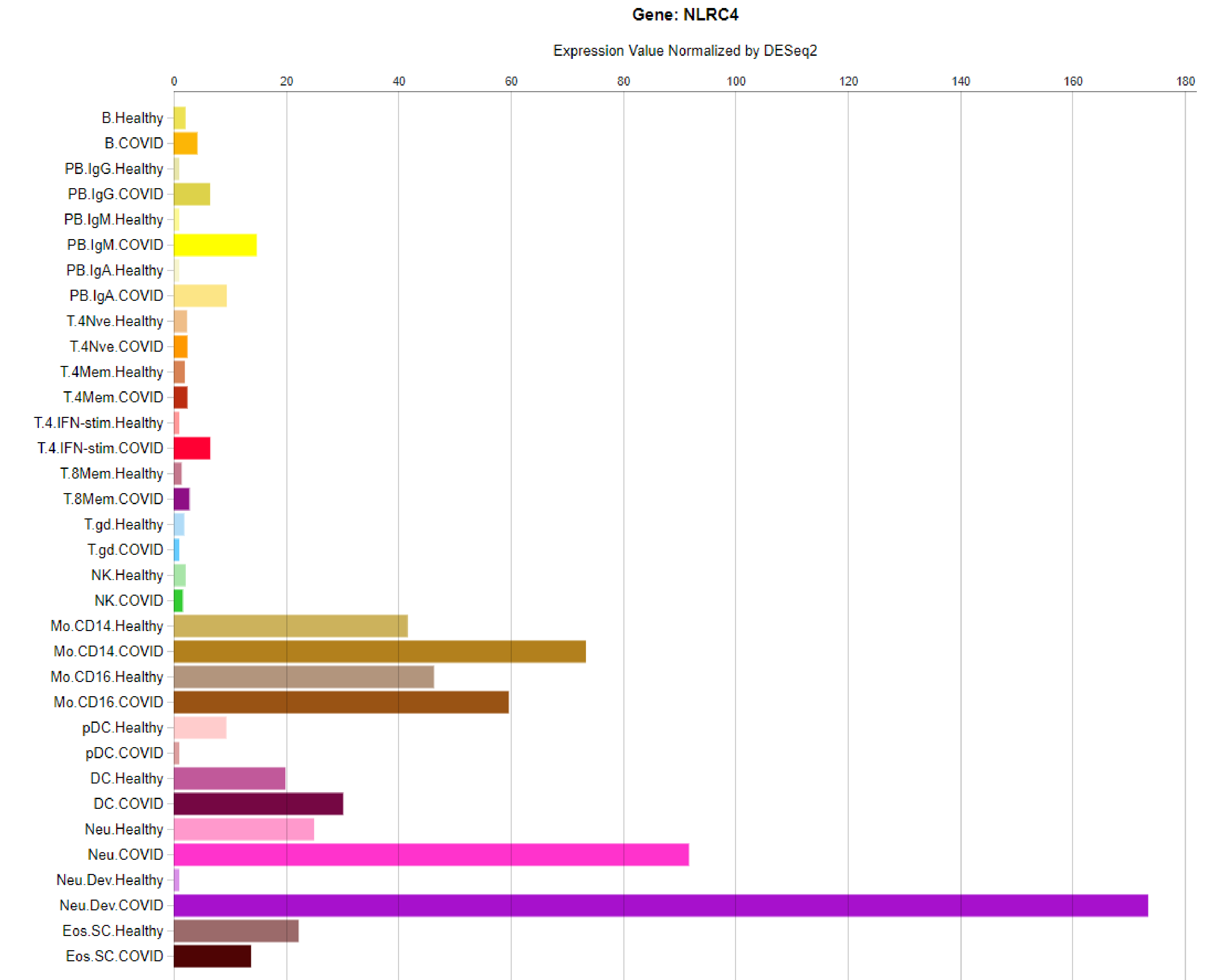

Gene: NLRC4
Expression Value Normalized by DESeq2
0
20
40
60
80
100
120
140
160
180
B.Healthy
B.COVID
PB.IgG.Healthy
PB.IgG.COVID
PB.IgM.Healthy
PB.IgM.COVID
PB.IgA.Healthy
PB.IgA.COVID
T.4Nve.Healthy
T.4Nve.COVID
T.4Mem.Healthy
T.4Mem.COVID
T.4.IFN-stim.Healthy
T.4.IFN-stim.COVID
T.8Mem.Healthy
T.8Mem.COVID
T.gd.Healthy
T.gd.COVID
NK.Healthy
NK.COVID
Mo.CD14.Healthy
Mo.CD14.COVID
Mo.CD16.Healthy
Mo.CD16.COVID
pDC.Healthy
pDC.COVID
DC.Healthy
DC.COVID
Neu.Healthy
Neu.COVID
Neu.Dev.Healthy
Neu.Dev.COVID
Eos.SC.Healthy
Eos.SC.COVID

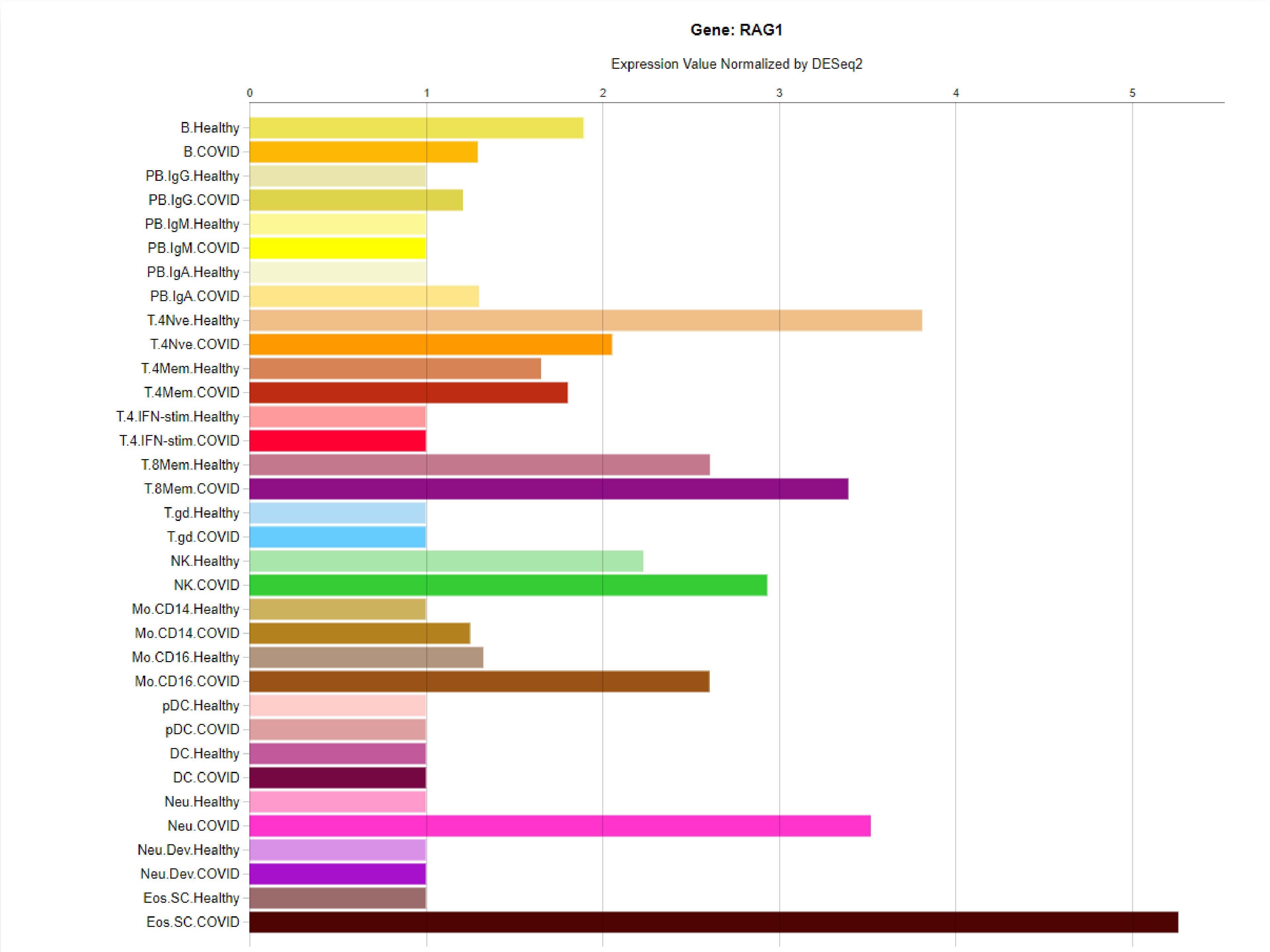

Gene: RAG1
Expression Value Normalized by DESeq2
0
1
2
3
4
5
B.Healthy
B.COVID
PB.IgG.Healthy
PB.IgG.COVID
PB.IgM.Healthy
PB.IgM.COVID
PB.IgA.Healthy
PB.IgA.COVID
T.4Nve.Healthy
T.4Nve.COVID
T.4Mem.Healthy
T.4Mem.COVID
T.4.IFN-stim.Healthy
T.4.IFN-stim.COVID
T.8Mem.Healthy
T.8Mem.COVID
T.gd.Healthy
T.gd.COVID
NK.Healthy
NK.COVID
Mo.CD14.Healthy
Mo.CD14.COVID
Mo.CD16.Healthy
Mo.CD16.COVID
pDC.Healthy
pDC.COVID
DC.Healthy
DC.COVID
Neu.Healthy
Neu.COVID
Neu.Dev.Healthy
Neu.Dev.COVID
Eos.SC.Healthy
Eos.SC.COVID

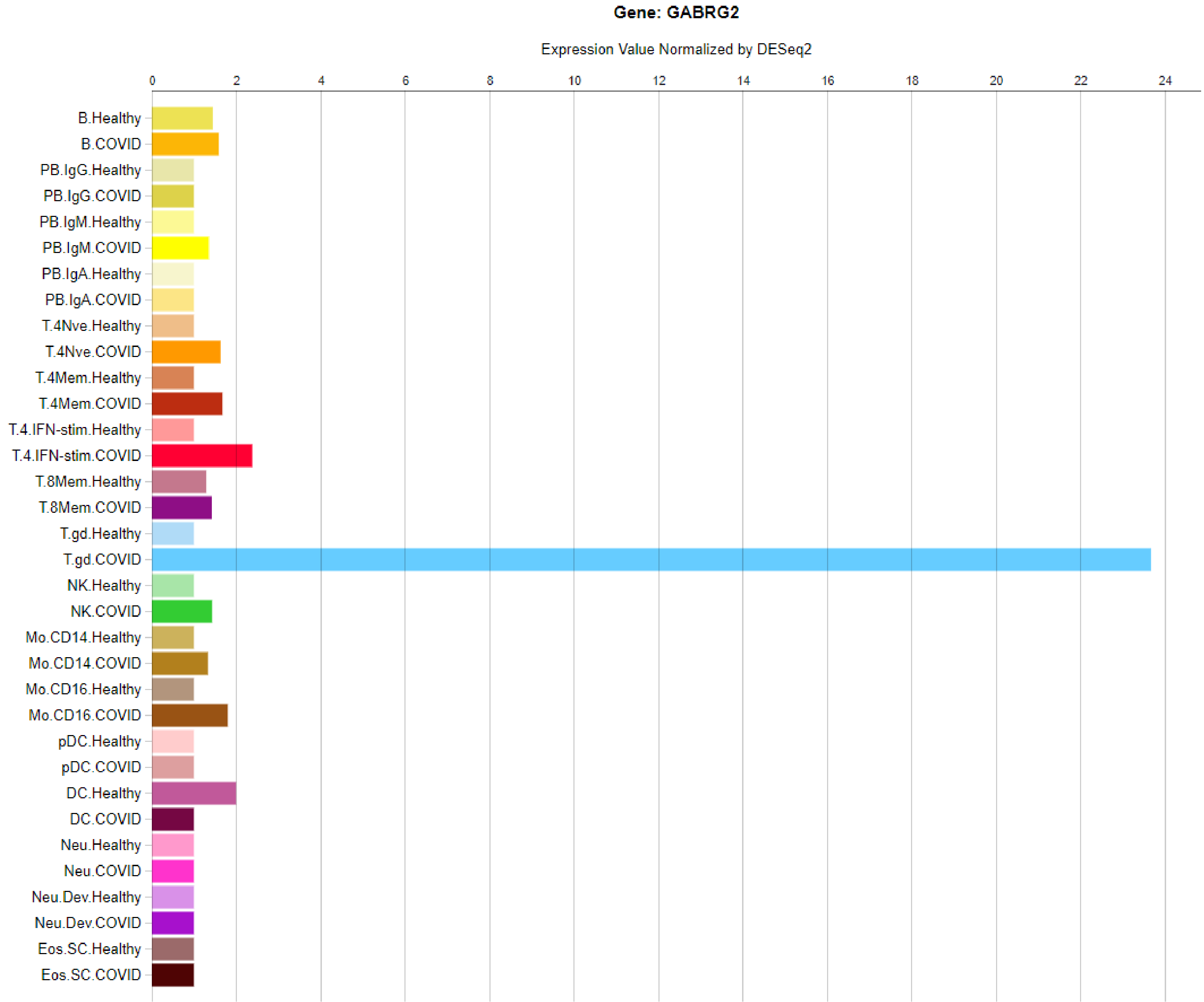
Gene: GABRG2
Expression Value Normalized by DESeq2
0
2
4
6
8
10
12
14
16
18
20
22
24
B.Healthy
B.COVID
PB.IgG.Healthy
PB.IgG.COVID
PB.IgM.Healthy
PB.IgM.COVID
PB.IgA.Healthy
PB.IgA.COVID
T.4Nve.Healthy
T.4Nve.COVID
T.4Mem.Healthy
T.4Mem.COVID
T.4.IFN-stim.Healthy
T.4.IFN-stim.COVID
T.8Mem.Healthy
T.8Mem.COVID
T.gd.Healthy
T.gd.COVID
NK.Healthy
NK.COVID
Mo.CD14.Healthy
Mo.CD14.COVID
Mo.CD16.Healthy
Mo.CD16.COVID
pDC.Healthy
pDC.COVID
DC.Healthy
DC.COVID
Neu.Healthy
Neu.COVID
Neu.Dev.Healthy
Neu.Dev.COVID
Eos.SC.Healthy
Eos.SC.COVID

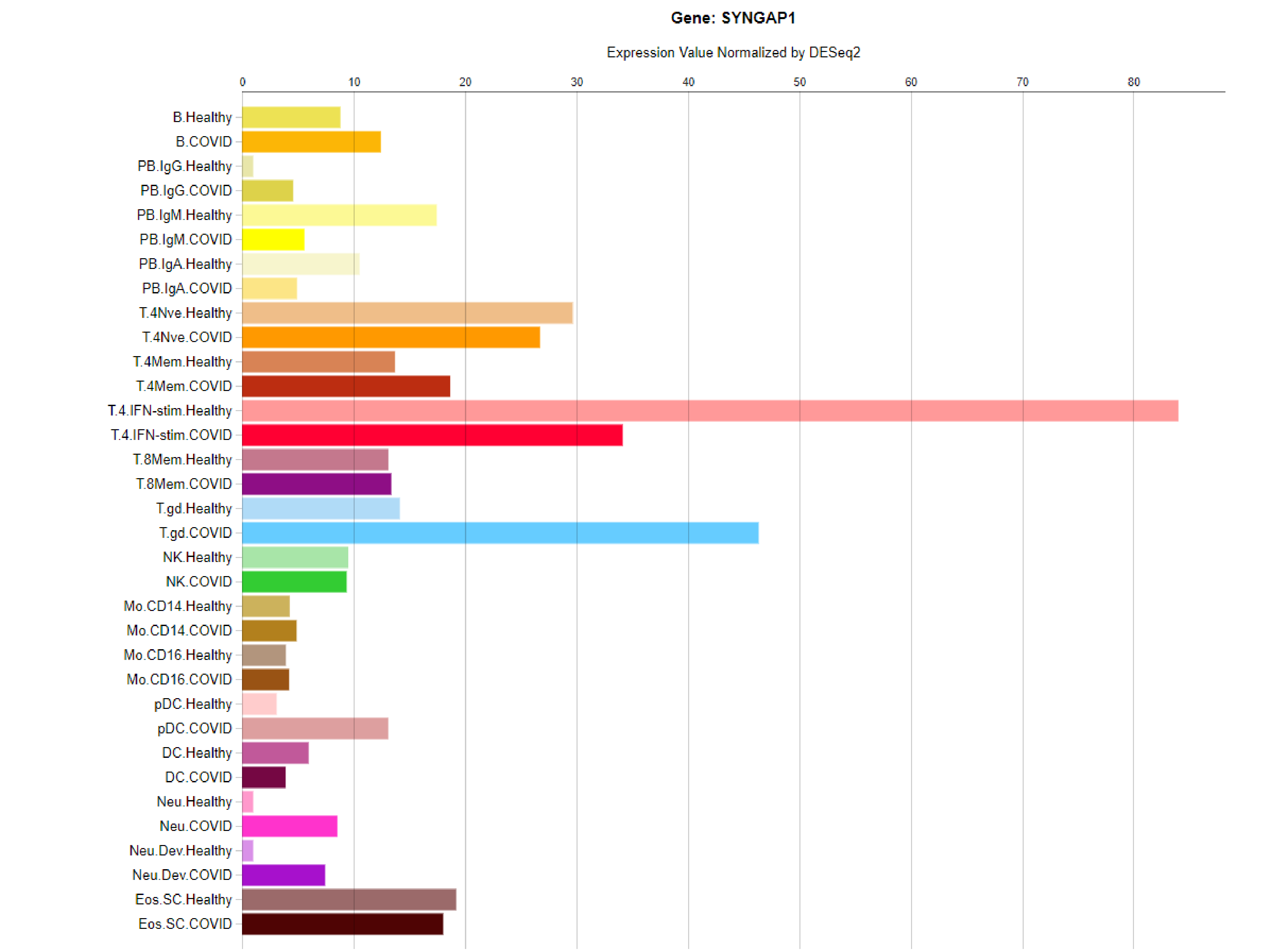
Gene: SYNGAP1
Expression Value Normalized by DESeq2
0
10
20
30
40
50
60
70
80
B.Healthy
B.COVID
PB.IgG.Healthy
PB.IgG.COVID
PB.IgM.Healthy
PB.IgM.COVID
PB.IgA.Healthy
PB.IgA.COVID
T.4Nve.Healthy
T.4Nve.COVID
T.4Mem.Healthy
T.4Mem.COVID
T.4.IFN-stim.Healthy
T.4.IFN-stim.COVID
T.8Mem.Healthy
T.8Mem.COVID
T.gd.Healthy
T.gd.COVID
NK.Healthy
NK.COVID
Mo.CD14.Healthy
Mo.CD14.COVID
Mo.CD16.Healthy
Mo.CD16.COVID
pDC.Healthy
pDC.COVID
DC.Healthy
DC.COVID
Neu.Healthy
Neu.COVID
Neu.Dev.Healthy
Neu.Dev.COVID
Eos.SC.Healthy
Eos.SC.COVID

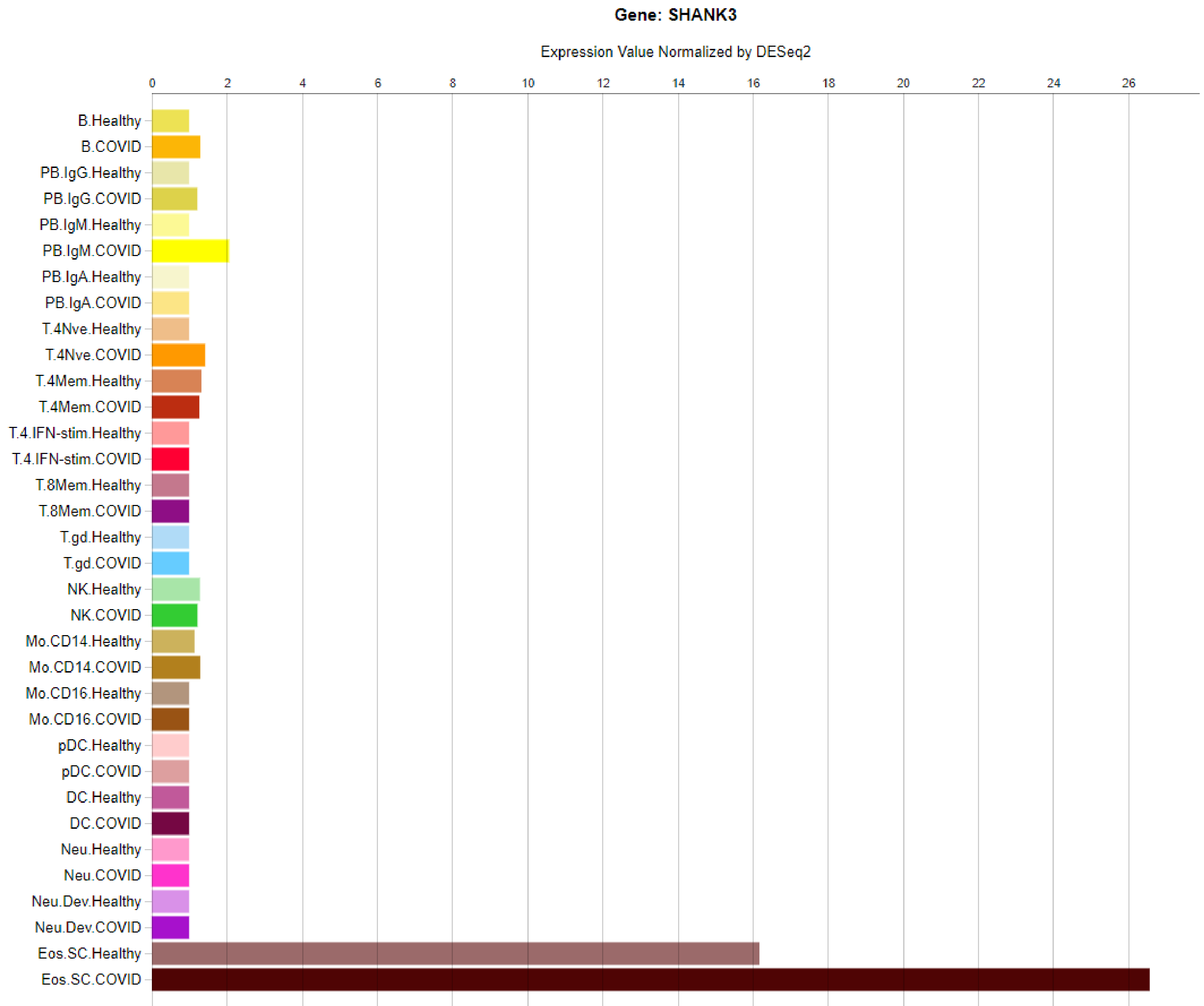
Gene: SHANK3
Expression Value Normalized by DESeq2
0
2
4
6
8
10
12
14
16
18
20
22
24
26
B.Healthy
B.COVID
PB.IgG.Healthy
PB.IgG.COVID
PB.IgM.Healthy
PB.IgM.COVID
PB.IgA.Healthy
PB.IgA.COVID
T.4Nve.Healthy
T.4Nve.COVID
T.4Mem.Healthy
T.4Mem.COVID
T.4.IFN-stim.Healthy
T.4.IFN-stim.COVID
T.8Mem.Healthy
T.8Mem.COVID
T.gd.Healthy
T.gd.COVID
NK.Healthy
NK.COVID
Mo.CD14.Healthy
Mo.CD14.COVID
Mo.CD16.Healthy
Mo.CD16.COVID
pDC.Healthy
pDC.COVID
DC.Healthy
DC.COVID
Neu.Healthy
Neu.COVID
Neu.Dev.Healthy
Neu.Dev.COVID
Eos.SC.Healthy
Eos.SC.COVID

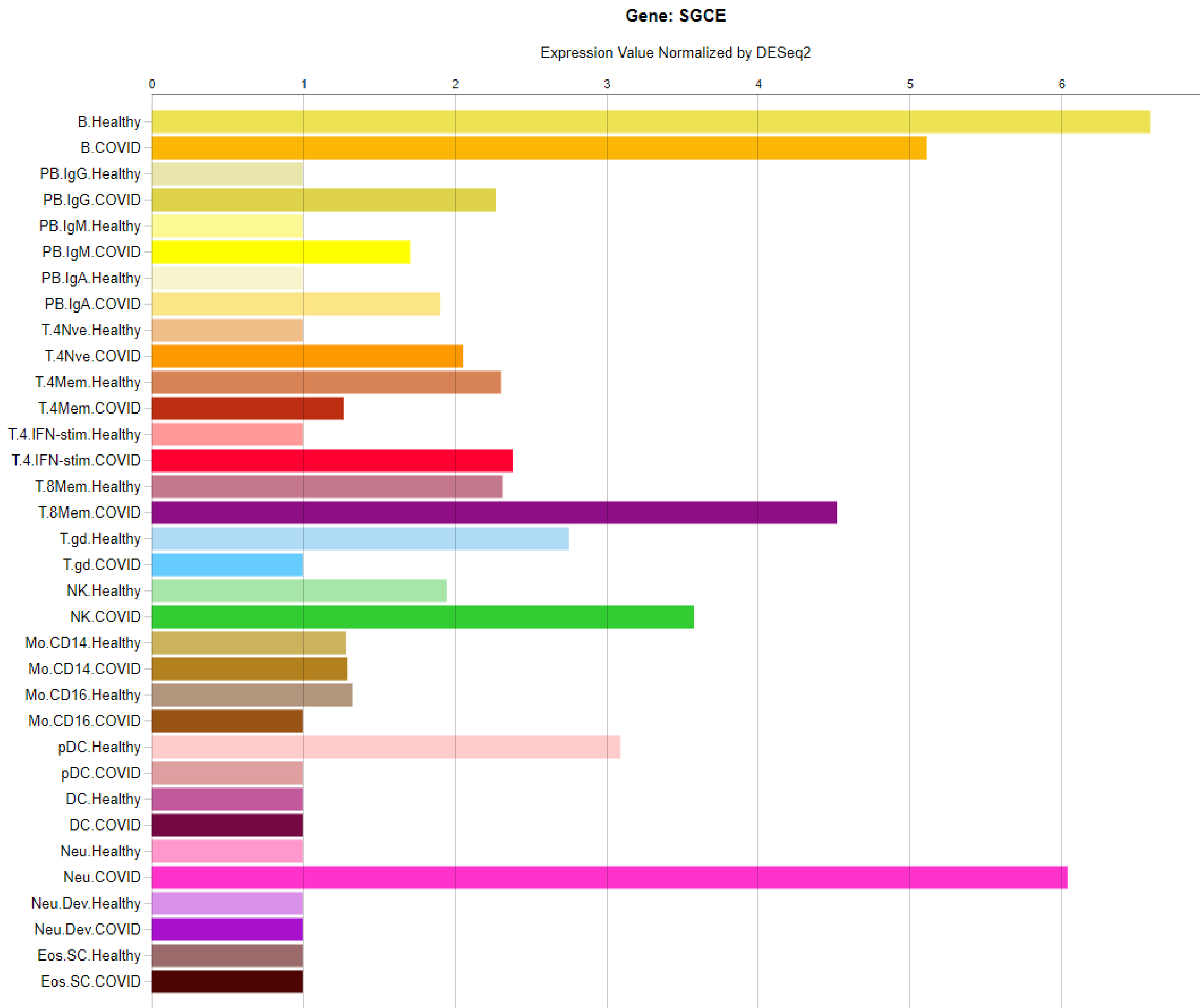
Gene: SGCE
Expression Value Normalized by DESeq2
0
1
2
3
4
5
6
B.Healthy
B.COVID
PB.IgG.Healthy
PB.IgG.COVID
PB.IgM.Healthy
PB.IgM.COVID
PB.IgA.Healthy
PB.IgA.COVID
T.4Nve.Healthy
T.4Nve.COVID
T.4Mem.Healthy
T.4Mem.COVID
T.4.IFN-stim.Healthy
T.4.IFN-stim.COVID
T.8Mem.Healthy
T.8Mem.COVID
T.gd.Healthy
T.gd.COVID
NK.Healthy
NK.COVID
Mo.CD14.Healthy
Mo.CD14.COVID
Mo.CD16.Healthy
Mo.CD16.COVID
pDC.Healthy
pDC.COVID
DC.Healthy
DC.COVID
Neu.Healthy
Neu.COVID
Neu.Dev.Healthy
Neu.Dev.COVID
Eos.SC.Healthy
Eos.SC.COVID

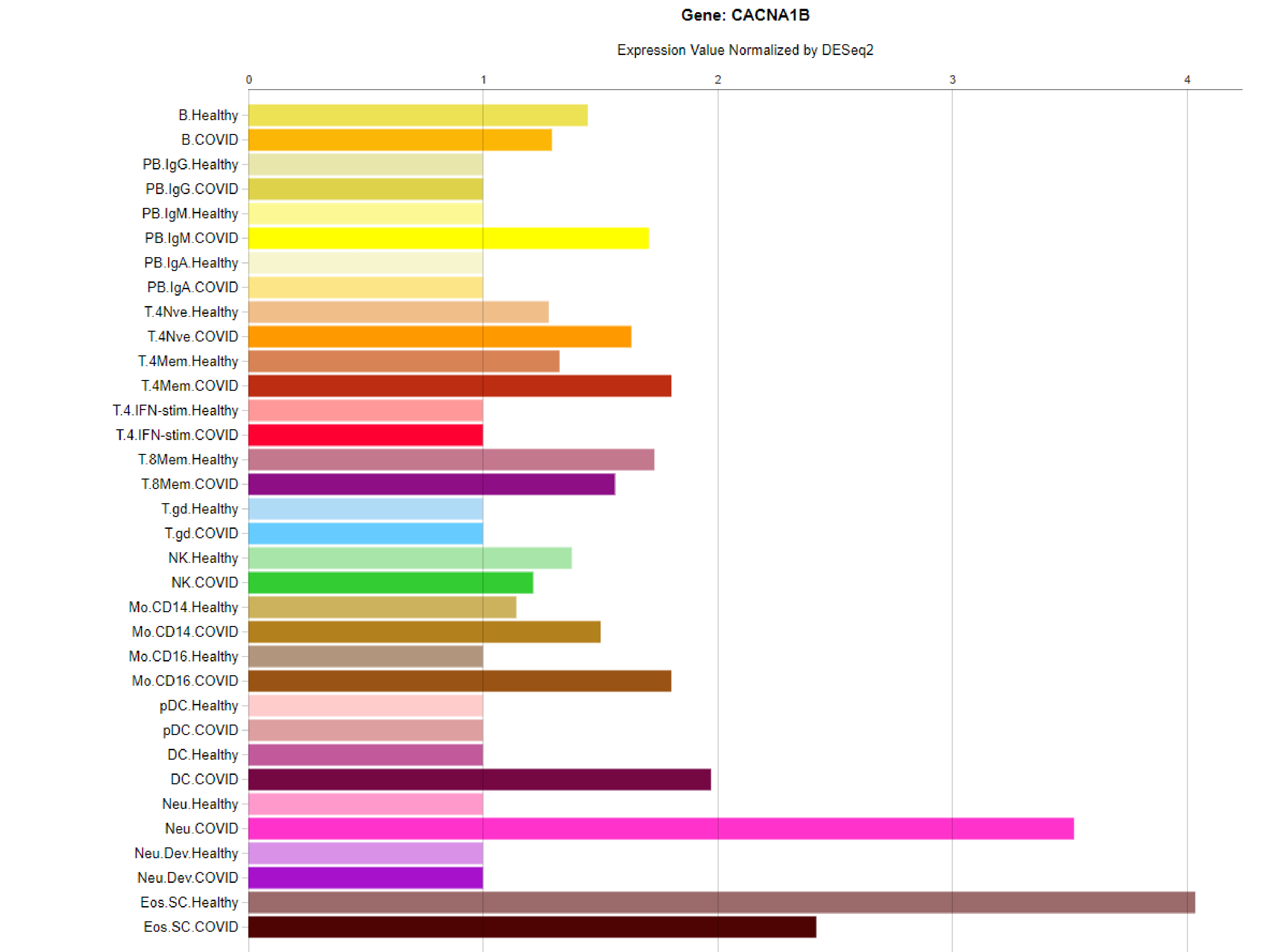

Gene: CACNA1B
Expression Value Normalized by DESeq2
0
1
2
3
4
B.Healthy
B.COVID
PB.IgG.Healthy
PB.IgG.COVID
PB.IgM.Healthy
PB.IgM.COVID
PB.IgA.Healthy
PB.IgA.COVID
T.4Nve.Healthy
T.4Nve.COVID
T.4Mem.Healthy
T.4Mem.COVID
T.4.IFN-stim.Healthy
T.4.IFN-stim.COVID
T.8Mem.Healthy
T.8Mem.COVID
T.gd.Healthy
T.gd.COVID
NK.Healthy
NK.COVID
Mo.CD14.Healthy
Mo.CD14.COVID
Mo.CD16.Healthy
Mo.CD16.COVID
pDC.Healthy
pDC.COVID
DC.Healthy
DC.COVID
Neu.Healthy
Neu.COVID
Neu.Dev.Healthy
Neu.Dev.COVID
Eos.SC.Healthy
Eos.SC.COVID

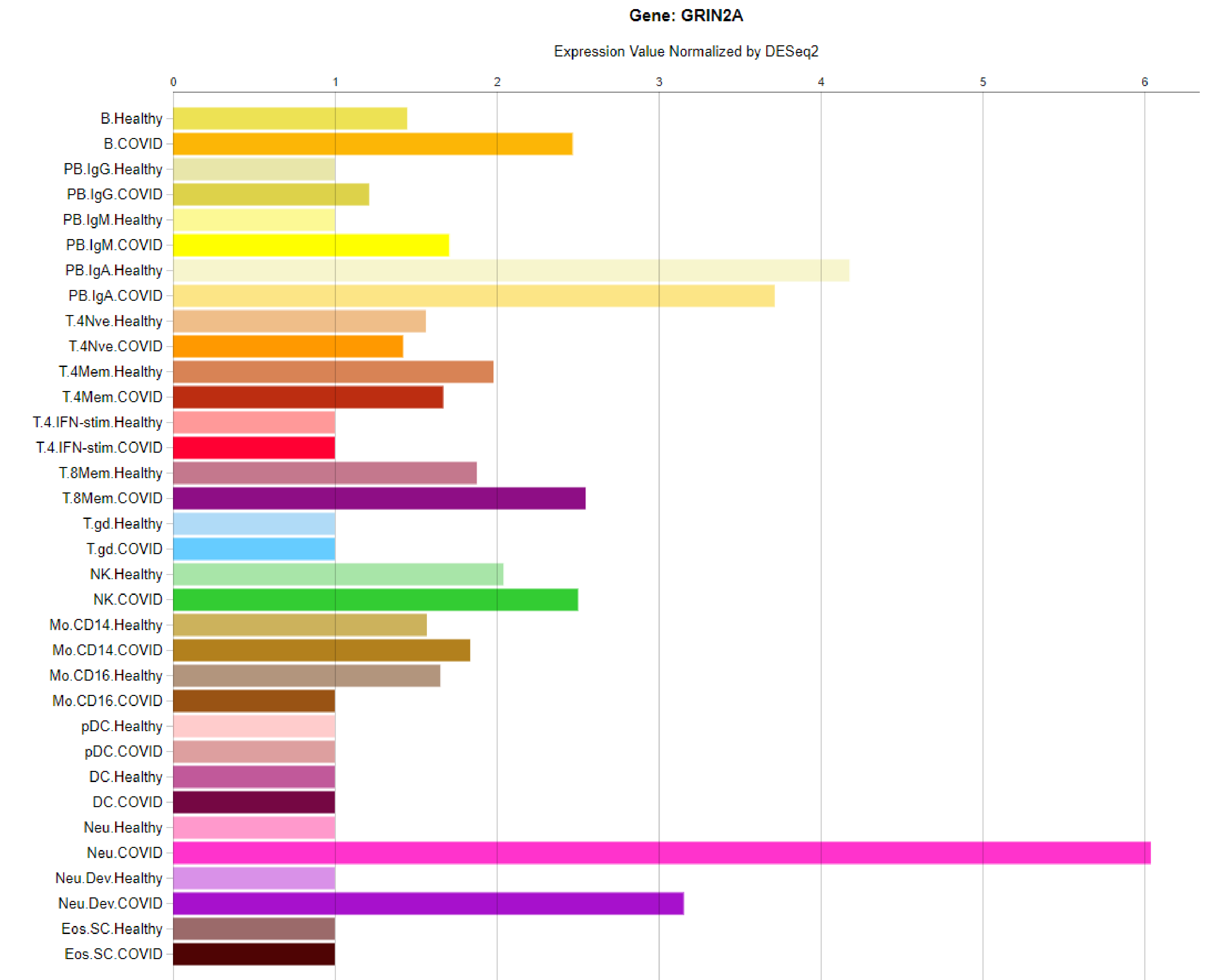

Gene: GRIN2A
Expression Value Normalized by DESeq2
0
1
2
3
4
5
6
B.Healthy
B.COVID
PB.IgG.Healthy
PB.IgG.COVID
PB.IgM.Healthy
PB.IgM.COVID
PB.IgA.Healthy
PB.IgA.COVID
T.4Nve.Healthy
T.4Nve.COVID
T.4Mem.Healthy
T.4Mem.COVID
T.4.IFN-stim.Healthy
T.4.IFN-stim.COVID
T.8Mem.Healthy
T.8Mem.COVID
T.gd.Healthy
T.gd.COVID
NK.Healthy
NK.COVID
Mo.CD14.Healthy
Mo.CD14.COVID
Mo.CD16.Healthy
Mo.CD16.COVID
pDC.Healthy
pDC.COVID
DC.Healthy
DC.COVID
Neu.Healthy
Neu.COVID
Neu.Dev.Healthy
Neu.Dev.COVID
Eos.SC.Healthy
Eos.SC.COVID
