## Supplementary figures and images for "Identification of ultra-rare genetic variants in Pediatric Acute Onset Neuropsychiatric Syndrome (PANS) by exome and whole genome sequencing"

### supp fig 2

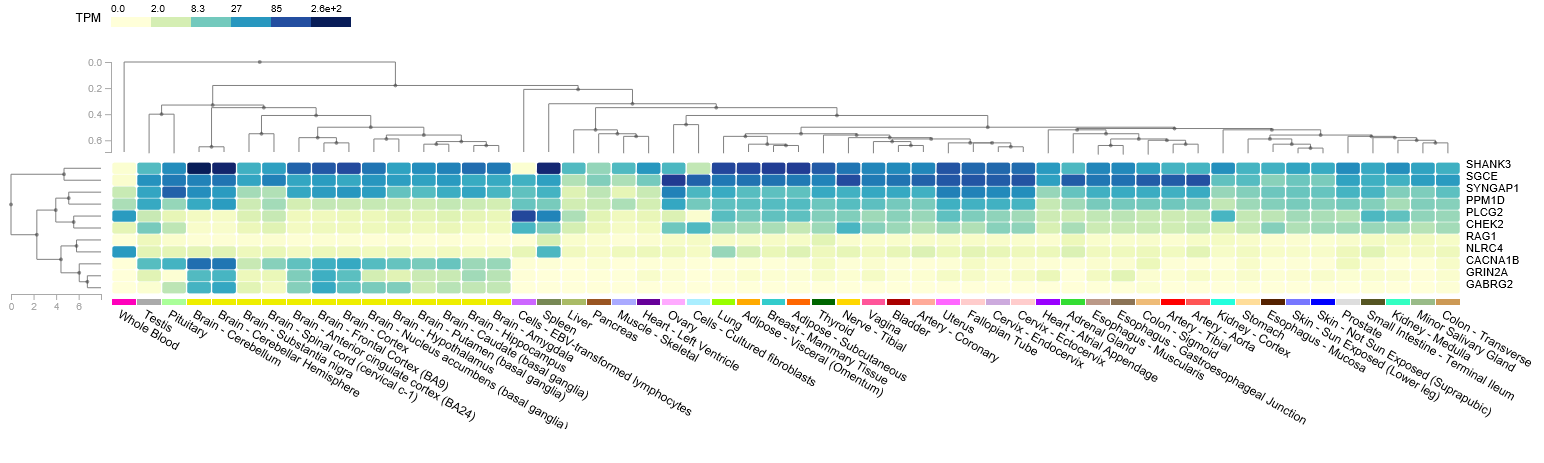
