## Supplementary material for "Identification of ultra-rare genetic variants in Pediatric Acute Onset Neuropsychiatric Syndrome (PANS) by exome and whole genome sequencing": supp fig 3

### Figure S3

**Legend.** Wheel plots showing single cell expression data from the developing mouse brain (http://mousebrain.org). The expression pattern is displayed as a UMAP clusters of2 different cell types determined by the expression pattern of cell specific markers. Slides 2 and 3 show the different cell types making up the clusters, while slide 4 shows the brain regions covered by the clusters. Slide 5 shows an example of a gene that is expressed in a cell type-specific manner, which was used to create the gene clusters. Slides 6-16 show the expression pattern at a common developmental stage. The box on the left of each wheel pot is the relative expression level within the different cell types.

### Cell types

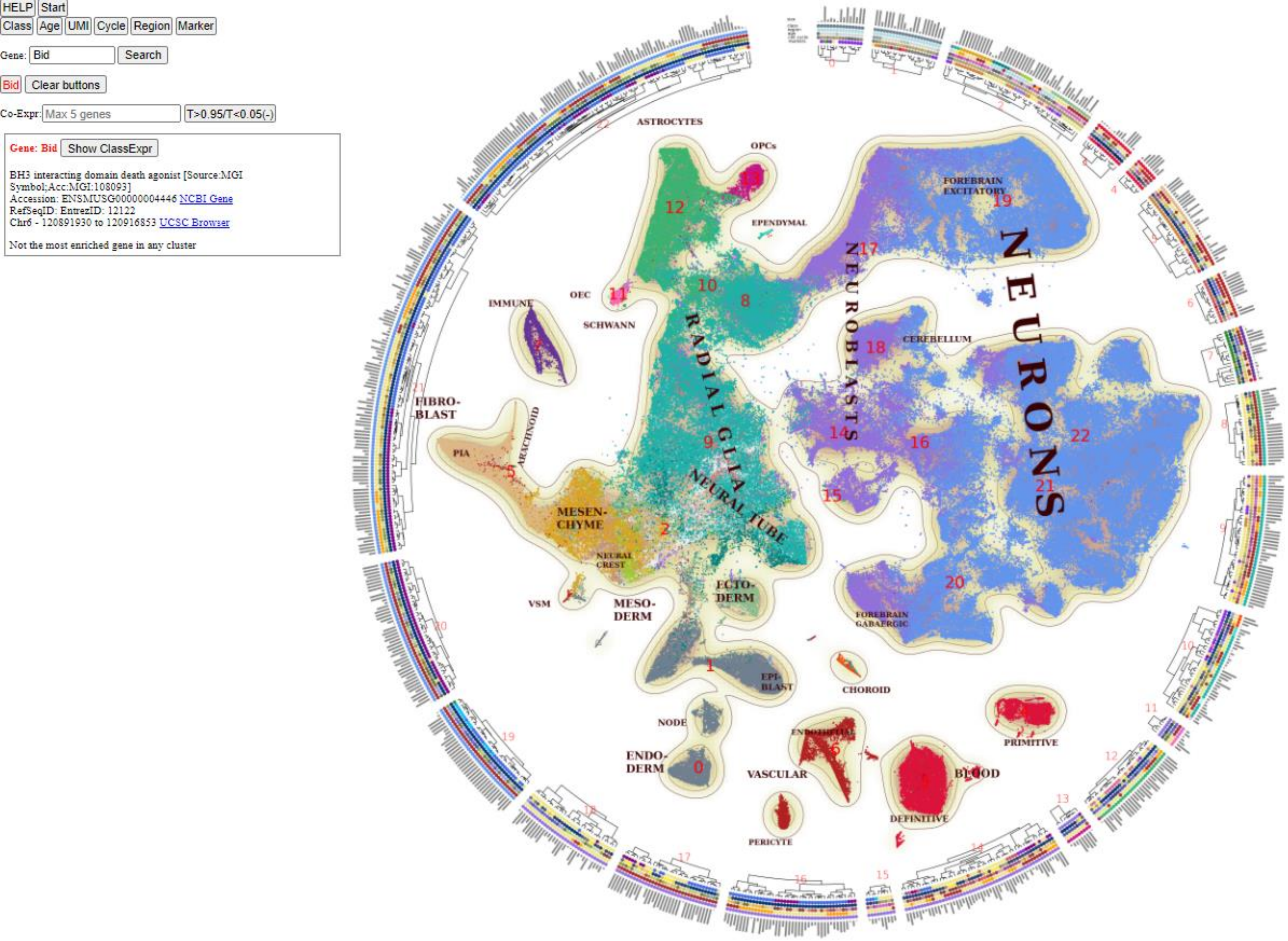
HELP
Start
Class
Age
UMI
Cycle
Region
Marker
Gene: Bid
Search
Bid
Clear buttons
Co-Expr: Max 5 genes
T>0.95/T<0.05(-)
Gene: Bid
Show ClassExpr
BH3 interacting domain death agonist [Source:MGI Symbol;Acc:MGI:108093]
Accession: ENSMUSG00000004446 NCBI Gene
RefSeqID: EntrezID: 12122
Chr6 - 120891930 to 120916853 UCSC Browser
Not the most enriched gene in any cluster
ASTROCYTES
OPCs
EPENDYMAL
FOREBRAIN EXCITATORY
NEURONS
NEUROBLASTS
OEC
IMMUNE
SCHWANN
RADIAL GLIA
CEREBELLUM
FIBRO-BLAST
ARACHNOID
PIA
NEURAL TUBE
MESEN-CHYME
NEURAL CREST
VSM
MESO-DERM
ECTO-DERM
FOREBRAIN GABAERGIC
EPI-BLAST
CHOROID
NODE
ENDO-DERM
ENDOTHELIAL
VASCULAR
BLOOD
PRIMITIVE
DEFINITIVE
PERICYTE

### Class

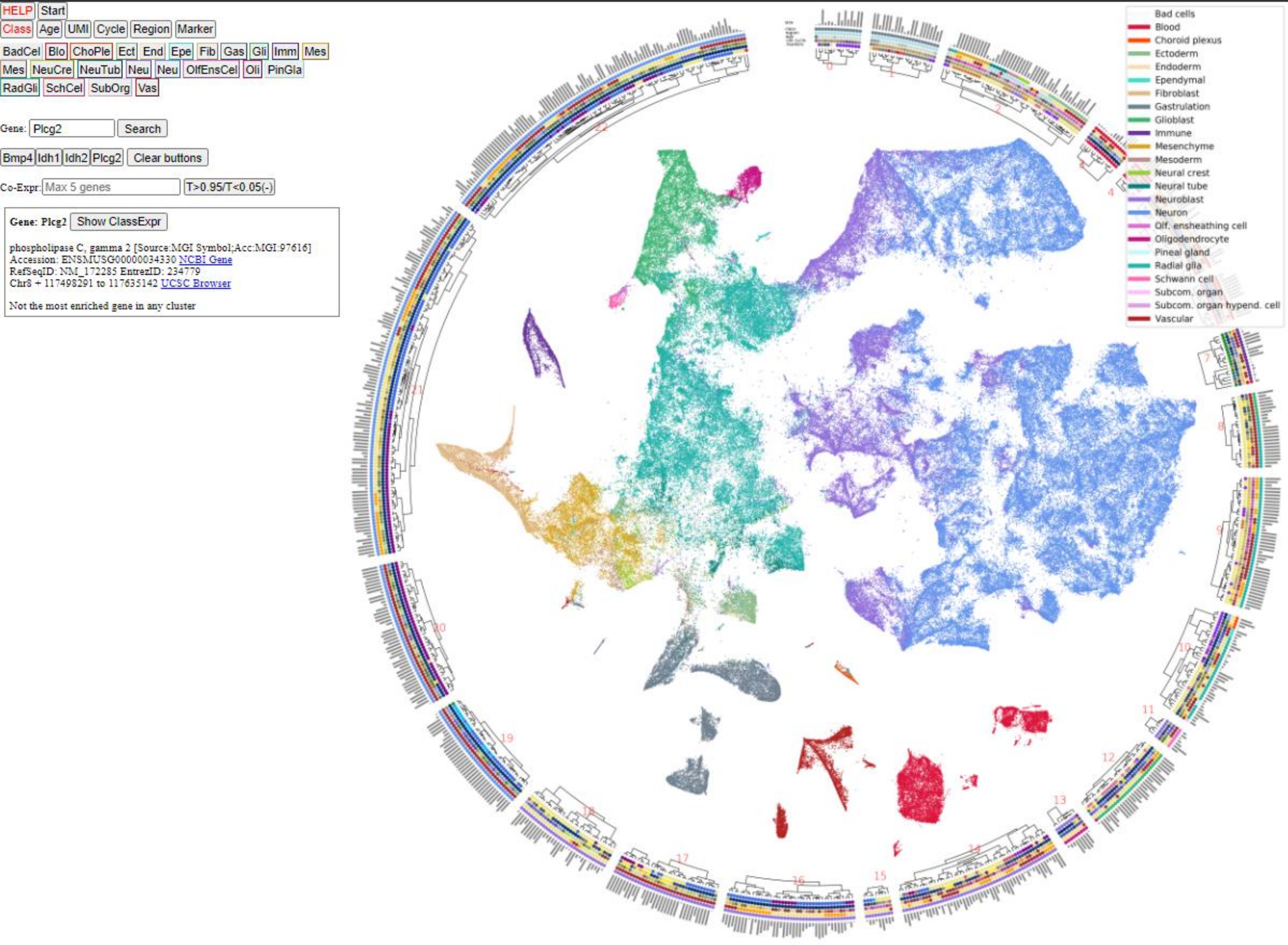
HELP
Start
Class
Age
UMI
Cycle
Region
Marker
BadCel Blo ChoPle Ect End Epe Fib Gas Gli Imm Mes
Mes NeuCre NeuTub Neu Neu OlfEnsCel Oli PinGla
RadGli SchCel SubOrg Vas
Gene: Plcg2
Search
Bmp4 Idh1 Idh2 Plcg2
Clear buttons
Co-Expr: Max 5 genes
T>0.95/T<0.05(-)
Gene: Plcg2
Show ClassExpr
phospholipase C, gamma 2 [Source:MGI Symbol;Acc:MGI:97616]
Accession: ENSMUSG00000034330 NCBI Gene
RefSeqID: NM_172285 EntrezID: 234779
Chr8 + 117498291 to 117635142 UCSC Browser
Not the most enriched gene in any cluster
Bad cells
Blood
Choroid plexus
Ectoderm
Endoderm
Ependymal
Fibroblast
Gastrulation
Glioblast
Immune
Mesenchyme
Mesoderm
Neural crest
Neural tube
Neuroblast
Neuron
Olf. ensheathing cell
Oligodendrocyte
Pineal gland
Radial glia
Schwann cell
Subcom. organ
Subcom. organ hypend. cell
Vascular

### Region

HELP Start

Class Age UMI Cycle Region Marker

Gene: Plcg2 Search

Bmp4 Idh1 Idh2 Plcg2 Clear buttons

Co-Expr: Max 5 genes T>0.95/T<0.05(-)

**Gene: Plcg2** Show ClassExpr

phospholipase C, gamma 2 [Source:MGI Symbol;Acc:MGI:97616]
Accession: ENSMUSG00000034330 NCBI Gene
RefSeqID: NM_172285 EntrezID: 234779
Chr8 + 117498291 to 117635142 UCSC Browser

Not the most enriched gene in any cluster

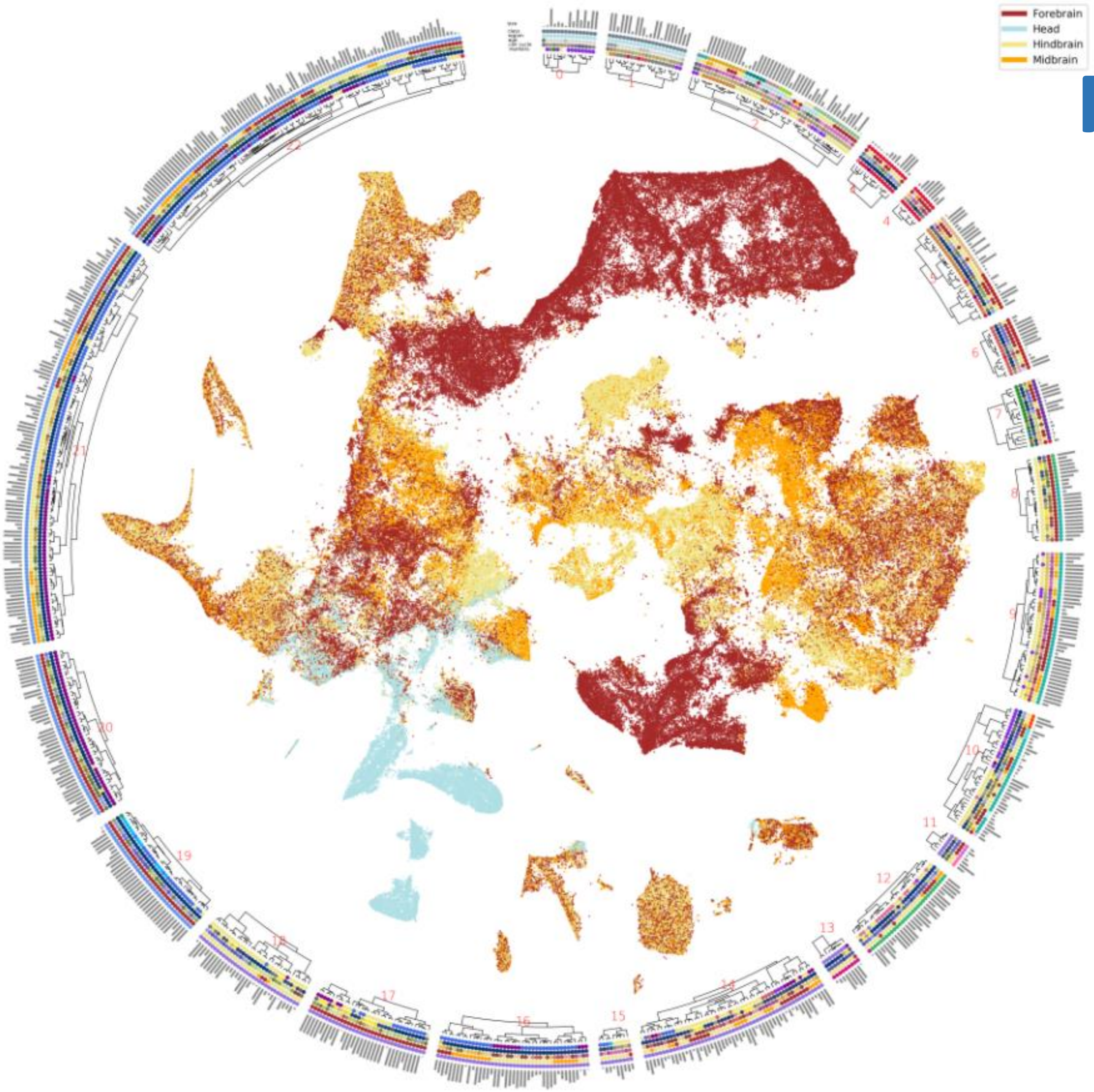

### Marker

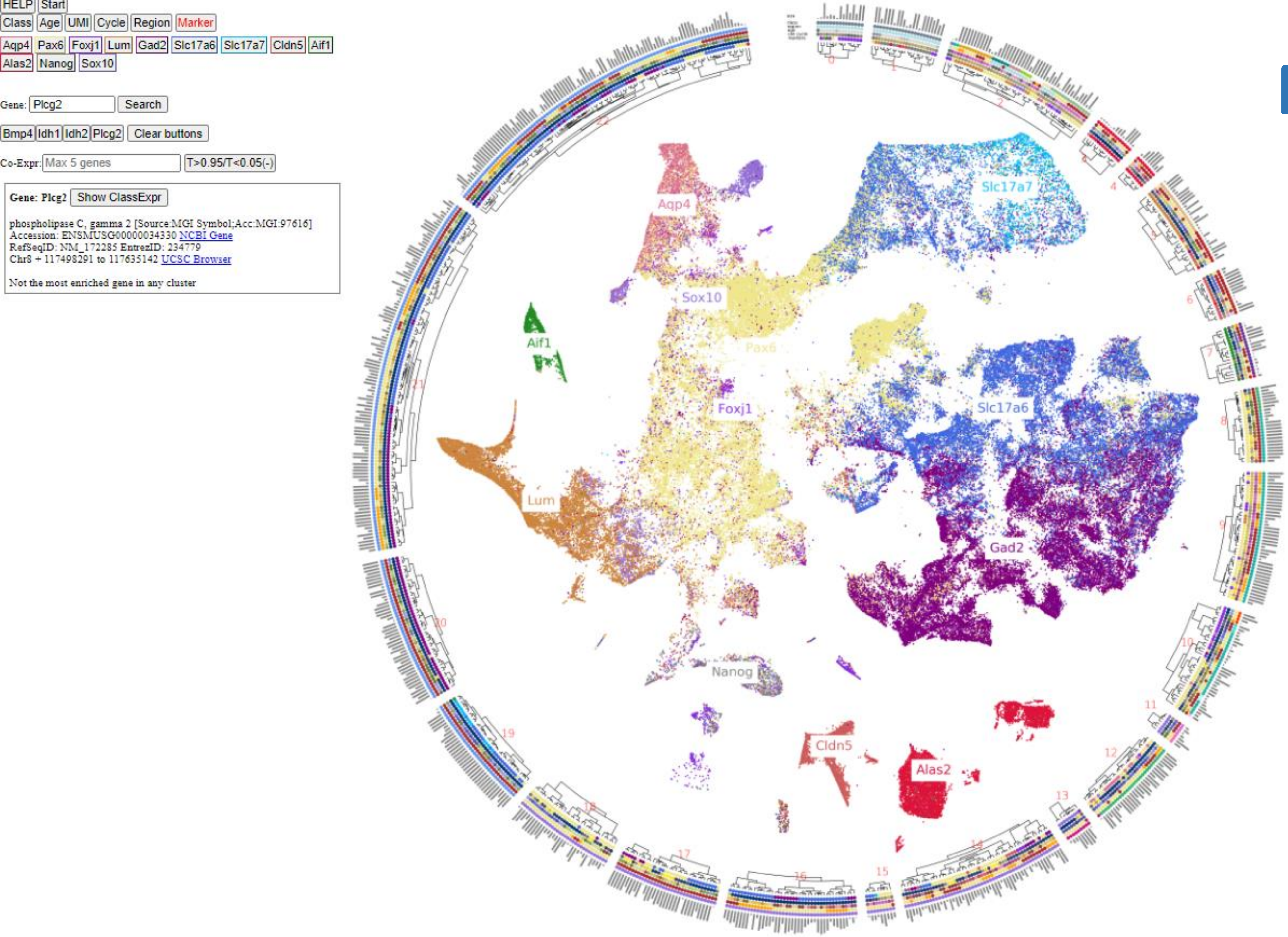
HELP Start
Class Age UMI Cycle Region Marker
Aqp4 Pax6 Foxj1 Lum Gad2 Slc17a6 Slc17a7 Cldn5 Aif1
Alas2 Nanog Sox10
Gene: Plcg2 Search
Bmp4 Idh1 Idh2 Plcg2 Clear buttons
Co-Expr: Max 5 genes T>0.95/T<0.05(-)
Gene: Plcg2 Show ClassExpr
phospholipase C, gamma 2 [Source:MGI Symbol;Acc:MGI:97616]
Accession: ENSMUSG00000034330 NCBI Gene
RefSeqID: NM_172285 EntrezID: 234779
Chr8 + 117498291 to 117635142 UCSC Browser
Not the most enriched gene in any cluster
Aqp4
Slc17a7
Sox10
Pax6
Aif1
Foxj1
Slc17a6
Lum
Gad2
Nanog
Cldn5
Alas2

HELP Start

Class Age UMI Cycle Region Marker

ene: Shank3 Search

Shank3 Clear buttons

o-Expr: Max 5 genes T>0.95/T<0.05(-)

**Gene: Shank3** Hide ClassExpr

| | |
|---|---|
| Bad cells | 0.00 |
| Blood | 0.01 |
| Choroid plexus | 0.21 |
| Ectoderm | 0.01 |
| Endoderm | 0.01 |
| Ependymal | 0.23 |
| Fibroblast | 0.04 |
| Gastrulation | 0.03 |
| Glioblast | 0.02 |
| Immune | 0.01 |
| Mesenchyme | 0.03 |
| Mesoderm | 0.03 |
| Neural crest | 0.02 |
| Neural tube | 0.03 |
| Neuroblast | 0.04 |
| Neuron | 0.10 |
| Olfactory ensheathing cell | 0.02 |
| Oligodendrocyte | 0.04 |
| Pineal gland | 0.04 |
| Radial glia | 0.03 |
| Schwann cell | 0.01 |
| Subcommissural organ | 0.13 |
| Vascular | 0.60 |

SH3 and multiple ankyrin repeat domains 3 [Source:MGI Symbol;Acc:MGI:1930016]
Accession: ENSMUSG00000022623 NCBI Gene
RefSeqID: EntrezID: 58234
Chr15 + 89499623 to 89560261 UCSC Browser

Not the most enriched gene in any cluster

< **Cluster: 302** > **AstroMix5. 451 cells.**

Class:Glioblast Subclass: Mixed region astrocytes
Mostly Hindbrain.

Show Distribution

AutoAnnotations:

@AEP @IEG AC-NFB Astro2 NbFor5 Rgl Rgl2 RglF2 RglH1 RglM2

Markers:

Agt Il33 Slc7a10 Cldn10 Slc6a9 Cyp2j9

Enriched TFs:

Dbx2 Olig1 Etv4 Rxrg Klf9 Klf15 Hes5 Bhlhe40

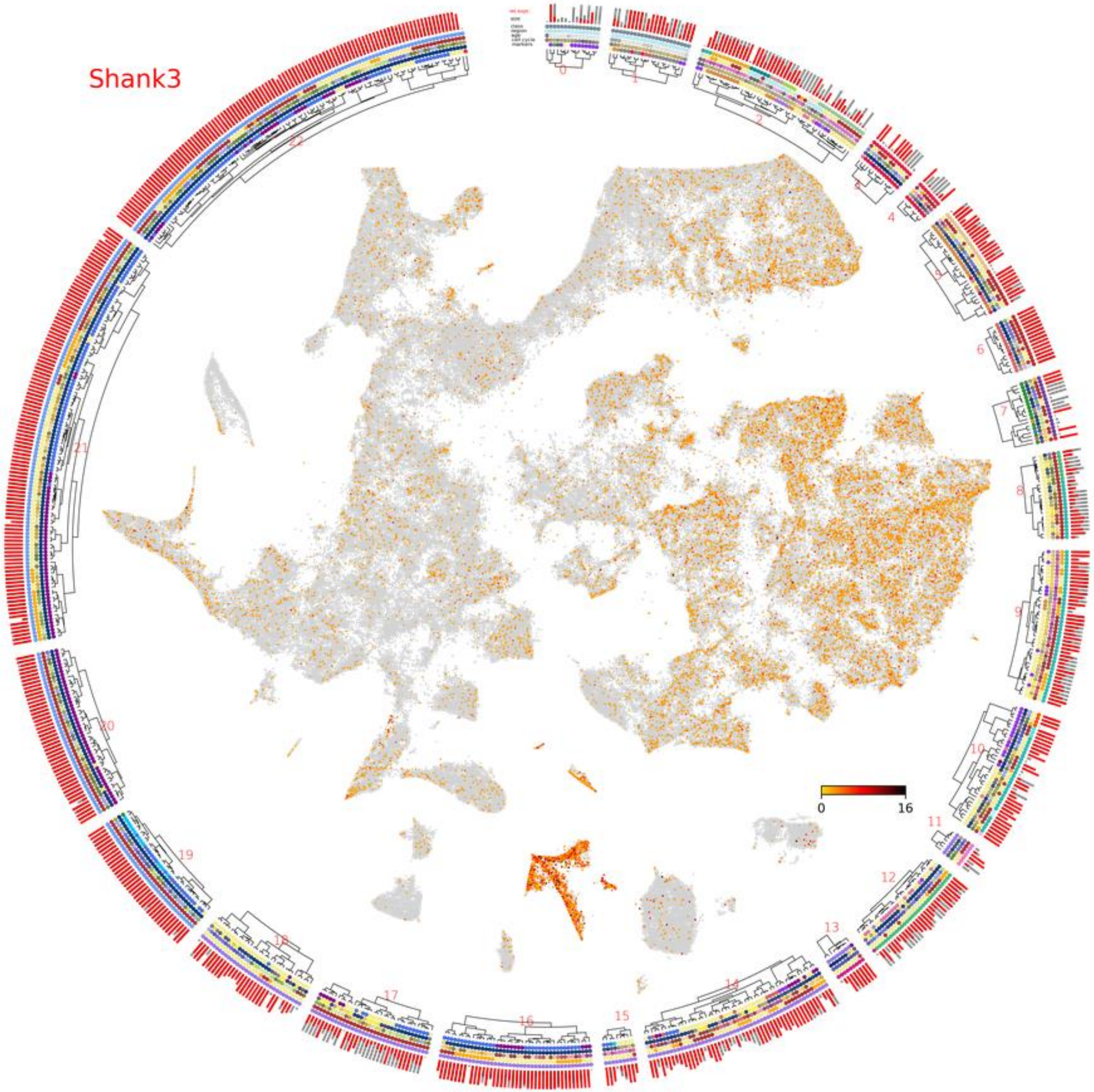

HELP Start

Class Age UMI Cycle Region Marker

Gene: Cacna1b Search

Ppm1d Sgce Cacna1b Clear buttons

Co-Expr: CACNA1B T>0.95/T<0.05(-)

**Gene: Cacna1b** Hide ClassExpr

| | |
|---|---|
| Bad cells | 0.00 |
| Blood | 0.01 |
| Choroid plexus | 0.03 |
| Ectoderm | 0.24 |
| Endoderm | 0.10 |
| Ependymal | 0.08 |
| Fibroblast | 0.02 |
| Gastrulation | 0.04 |
| Glioblast | 0.01 |
| Immune | 0.03 |
| Mesenchyme | 0.01 |
| Mesoderm | 0.01 |
| Neural crest | 0.03 |
| Neural tube | 0.07 |
| Neuroblast | 0.13 |
| Neuron | 0.37 |
| Olfactory ensheathing cell | 0.06 |
| Oligodendrocyte | 0.08 |
| Pineal gland | 0.01 |
| Radial glia | 0.02 |
| Schwann cell | 0.01 |
| Subcommissural organ | 0.02 |
| Vascular | 0.03 |

calcium channel, voltage-dependent, N type, alpha 1B subunit
[Source:MGI Symbol;Acc:MGI:88296]
Accession: ENSMUSG00000004113 NCBI Gene
RefSeqID: EntrezID: 12287
Chr2 - 24603887 to 24763152 UCSC Browser

Not the most enriched gene in any cluster

< **Cluster: 566** > **Neur566. 338 cells.**

Class:Neuron Subclass: Forebrain GABAergic
Mostly ForebrainVentral.

Show Distribution

AutoAnnotations:
@GABA Dlx IntHc1 MGEInt NbFor5 NbInh1

Markers:
Sst Fam135b Fam222a Sox6 Gm45881 Dlgap1

Enriched TFs:
Lhx6 Arx Dlx2 Dlx5 Dlx1 Sox6 Mafb Sp9

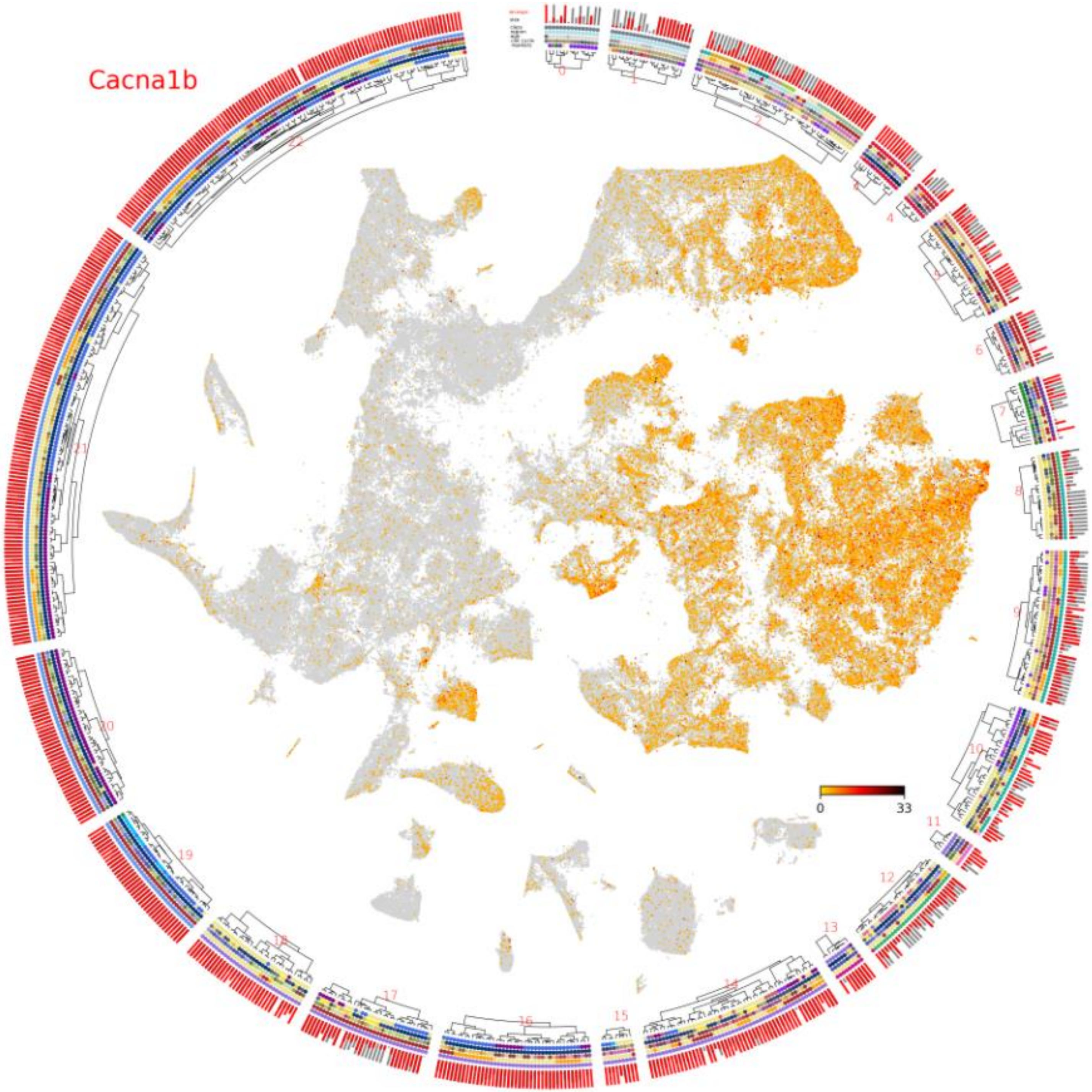

HELP Start

Class Age UMI Cycle Region Marker

Gene: Syngap1 Search

Syngap1 Clear buttons

Co-Expr: Max 5 genes T>0.95/T<0.05(-)

**Gene: Syngap1** Hide ClassExpr

| | |
|---|---|
| Bad cells | 0.00 |
| Blood | 0.00 |
| Choroid plexus | 0.05 |
| Ectoderm | 0.01 |
| Endoderm | 0.01 |
| Ependymal | 0.08 |
| Fibroblast | 0.01 |
| Gastrulation | 0.01 |
| Glioblast | 0.01 |
| Immune | 0.00 |
| Mesenchyme | 0.02 |
| Mesoderm | 0.01 |
| Neural crest | 0.01 |
| Neural tube | 0.02 |
| Neuroblast | 0.01 |
| Neuron | 0.02 |
| Olfactory ensheathing cell | 0.01 |
| Oligodendrocyte | 0.01 |
| Pineal gland | 0.01 |
| Radial glia | 0.02 |
| Schwann cell | 0.01 |
| Subcommissural organ | 0.01 |
| Vascular | 0.01 |

synaptic Ras GTPase activating protein 1 homolog (rat) [Source:MGI Symbol;Acc:MGI:3039785]
Accession: ENSMUSG00000067629 NCBI Gene
RefSeqID: EntrezID: 240057
Chr17 + 26941253 to 26972434 UCSC Browser

Not the most enriched gene in any cluster

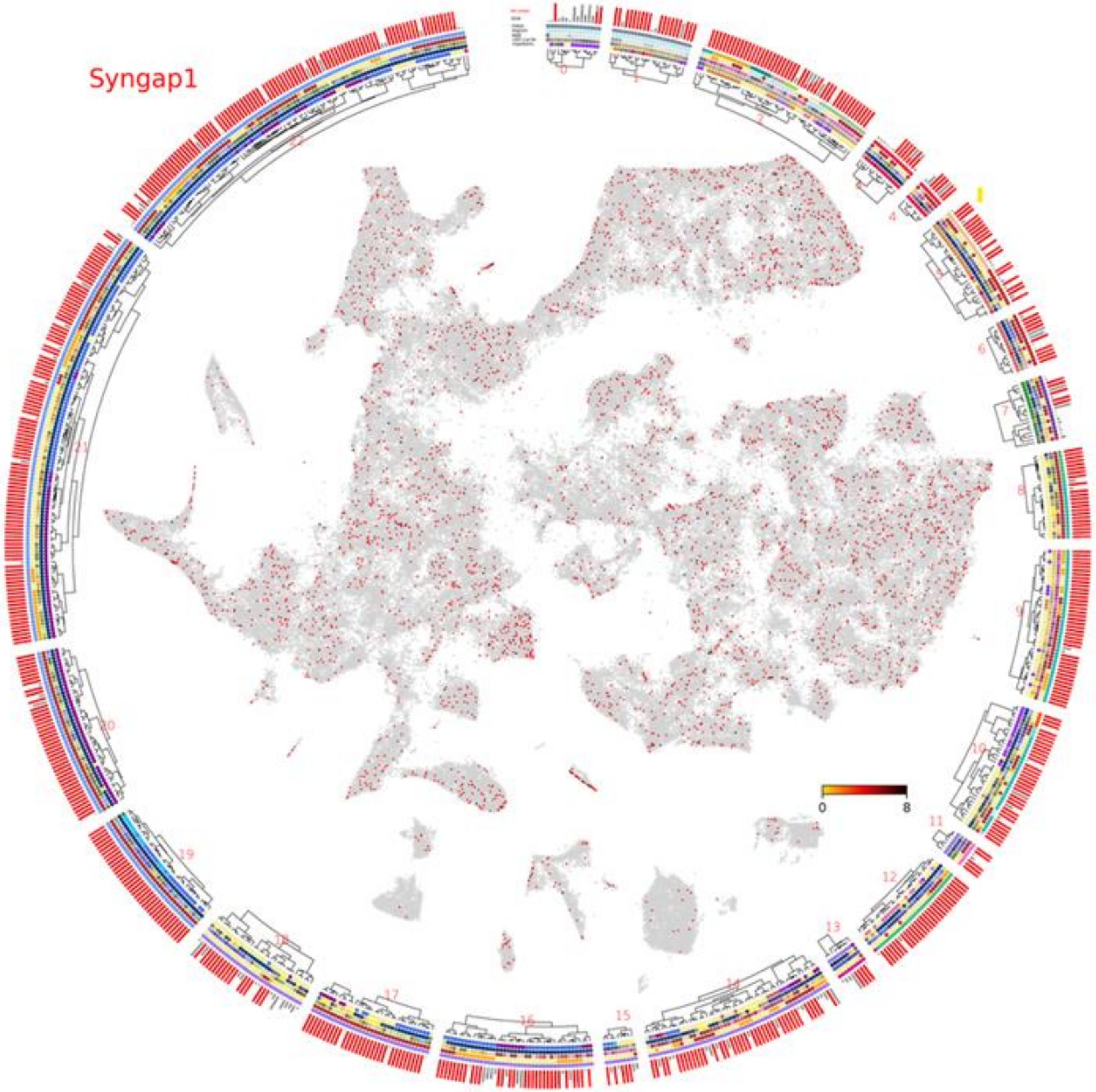

HELP Start

Class Age UMI Cycle Region Marker

Gene: Sgce Search

Ppm1d Sgce Clear buttons

Co-Expr: Max 5 genes T>0.95/T<0.05(-)

**Gene: Sgce** Hide ClassExpr

| | |
|---|---|
| Bad cells | 0.14 |
| Blood | 0.03 |
| Choroid plexus | 0.58 |
| Ectoderm | 0.38 |
| Endoderm | 0.56 |
| Ependymal | 0.87 |
| Fibroblast | 0.89 |
| Gastrulation | 0.30 |
| Glioblast | 0.57 |
| Immune | 0.32 |
| Mesenchyme | 0.55 |
| Mesoderm | 0.53 |
| Neural crest | 0.34 |
| Neural tube | 0.50 |
| Neuroblast | 0.37 |
| Neuron | 0.32 |
| Olfactory ensheathing cell | 0.46 |
| Oligodendrocyte | 0.44 |
| Pineal gland | 0.41 |
| Radial glia | 0.60 |
| Schwann cell | 0.46 |
| Subcommissural organ | 0.45 |
| Vascular | 0.78 |

sarcoglycan, epsilon [Source:MGI Symbol;Acc:MGI:1329042]
Accession: ENSMUSG00000004631 NCBI Gene
RefSeqID: EntrezID: -
Chr6 - 4674350 to 4747207 UCSC Browser

Not the most enriched gene in any cluster

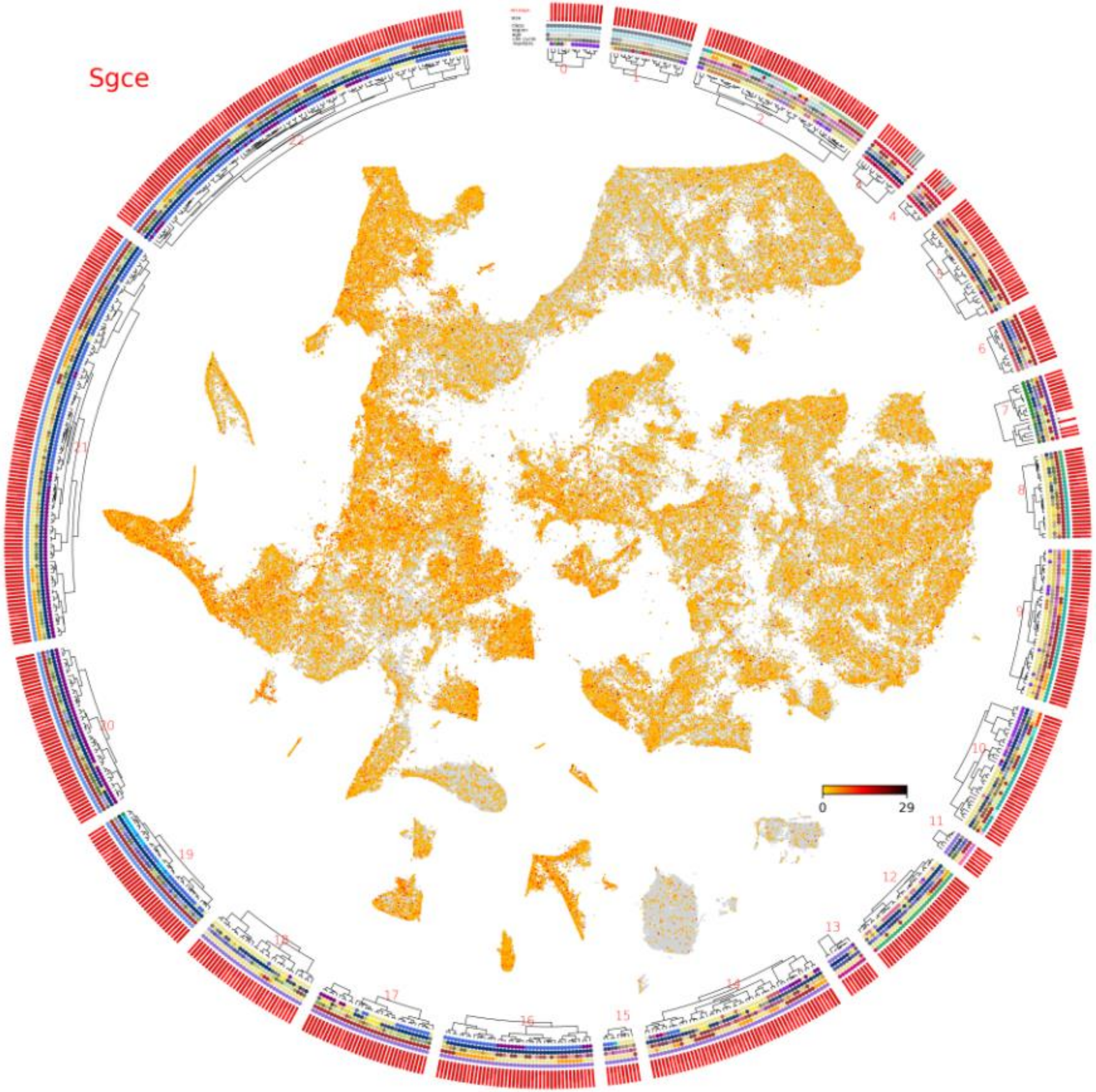

HELP Start

Class Age UMI Cycle Region Marker

Gene: Grin2a Search

Syngap1 Grin2a Clear buttons

Co-Expr: Max 5 genes T>0.95/T<0.05(-)

**Gene: Grin2a** Hide ClassExpr

| | |
|---|---|
| Bad cells | 0.00 |
| Blood | 0.01 |
| Choroid plexus | 0.01 |
| Ectoderm | 0.01 |
| Endoderm | 0.00 |
| Ependymal | 0.28 |
| Fibroblast | 0.01 |
| Gastrulation | 0.01 |
| Glioblast | 0.05 |
| Immune | 0.01 |
| Mesenchyme | 0.03 |
| Mesoderm | 0.01 |
| Neural crest | 0.01 |
| Neural tube | 0.05 |
| Neuroblast | 0.04 |
| Neuron | 0.12 |
| Olfactory ensheathing cell | 0.01 |
| Oligodendrocyte | 0.02 |
| Pineal gland | 0.00 |
| Radial glia | 0.05 |
| Schwann cell | 0.02 |
| Subcommissural organ | 0.01 |
| Vascular | 0.09 |

glutamate receptor, ionotropic, NMDA2A (epsilon 1) [Source:MGI Symbol;Acc:MGI:95820]
Accession: ENSMUSG00000059003 NCBI Gene
RefSeqID: EntrezID: -
Chr16 - 9567898 to 9995560 UCSC Browser

Not the most enriched gene in any cluster

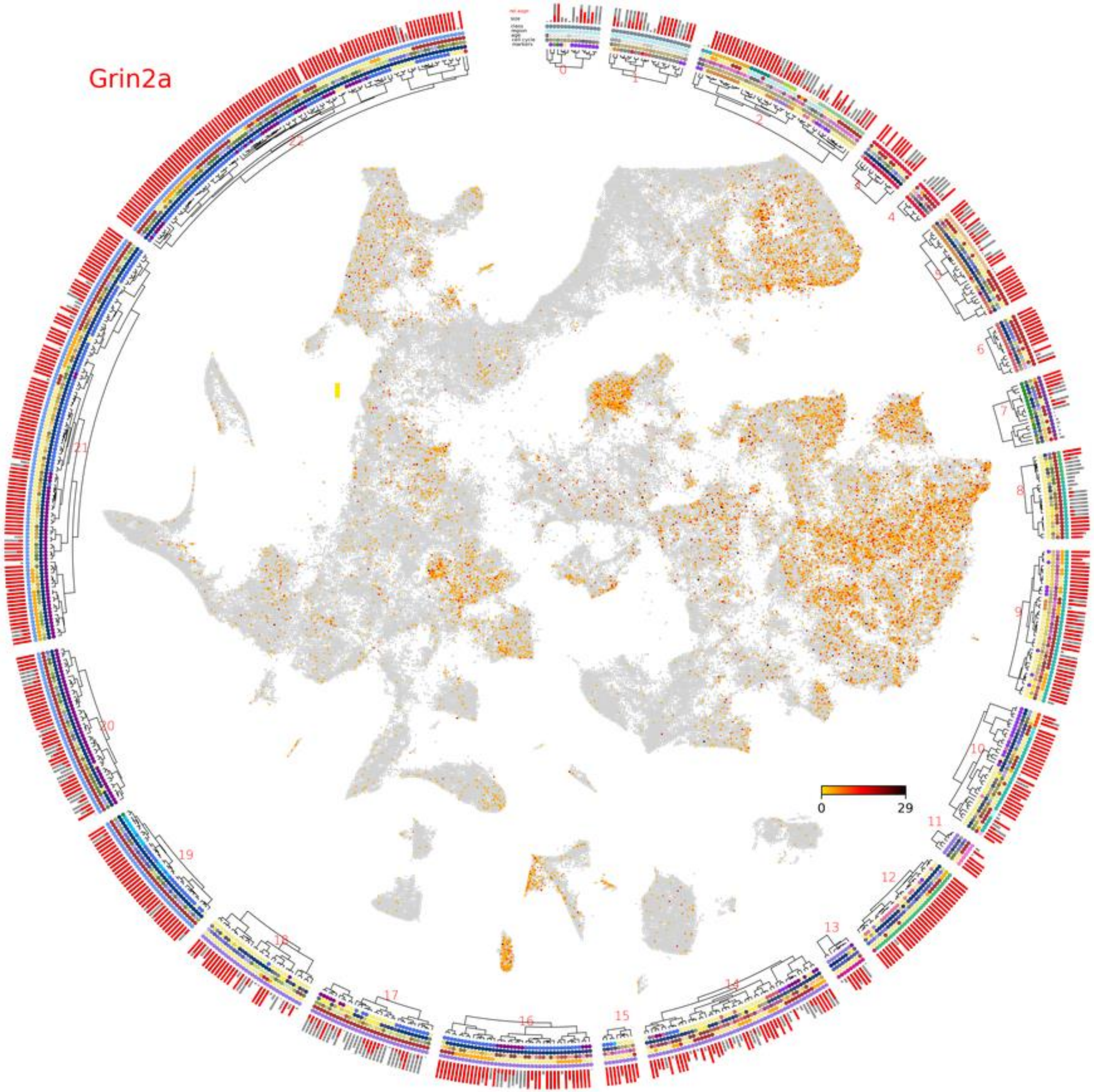

HELP Start

Class Age UMI Cycle Region Marker

Gene: Nlrc4 Search

Grin2a Nlrc4 Clear buttons

Co-Expr: Max 5 genes T>0.95/T<0.05(-)

**Gene: Nlrc4** Hide ClassExpr

| | |
|---|---|
| Bad cells | 0.01 |
| Blood | 0.01 |
| Choroid plexus | 0.03 |
| Ectoderm | 0.03 |
| Endoderm | 0.02 |
| Ependymal | 0.06 |
| Fibroblast | 0.02 |
| Gastrulation | 0.03 |
| Glioblast | 0.01 |
| Immune | 0.11 |
| Mesenchyme | 0.03 |
| Mesoderm | 0.02 |
| Neural crest | 0.02 |
| Neural tube | 0.02 |
| Neuroblast | 0.01 |
| Neuron | 0.01 |
| Olfactory ensheathing cell | 0.02 |
| Oligodendrocyte | 0.01 |
| Pineal gland | 0.01 |
| Radial glia | 0.02 |
| Schwann cell | 0.01 |
| Subcommissural organ | 0.00 |
| Vascular | 0.02 |

NLR family, CARD domain containing 4 [Source:MGI Symbol;Acc:MGI:3036243]
Accession: ENSMUSG00000039193 NCBI Gene
RefSeqID: EntrezID: 268973
Chr17 - 74426295 to 74459108 UCSC Browser

Not the most enriched gene in any cluster

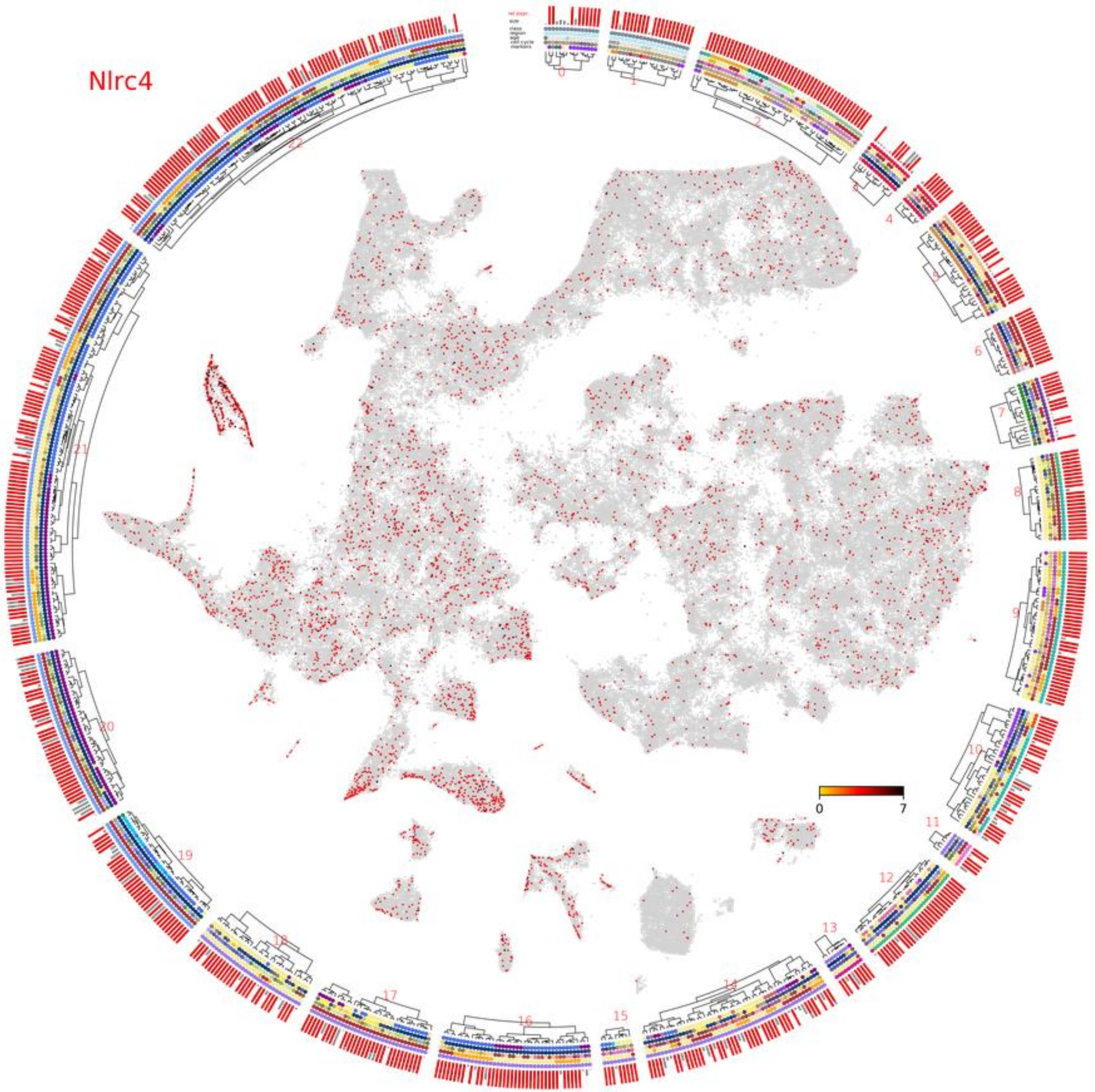

HELP Start

Class Age UMI Cycle Region Marker

Gene: Ppm1d Search

Ppm1d Clear buttons

Co-Expr: Max 5 genes T>0.95/T<0.05(-)

**Gene: Ppm1d** Hide ClassExpr

| | |
|---|---|
| Bad cells | 0.16 |
| Blood | 0.16 |
| Choroid plexus | 0.23 |
| Ectoderm | 0.28 |
| Endoderm | 0.29 |
| Ependymal | 0.23 |
| Fibroblast | 0.15 |
| Gastrulation | 0.31 |
| Glioblast | 0.20 |
| Immune | 0.11 |
| Mesenchyme | 0.33 |
| Mesoderm | 0.31 |
| Neural crest | 0.40 |
| Neural tube | 0.40 |
| Neuroblast | 0.28 |
| Neuron | 0.18 |
| Olfactory ensheathing cell | 0.23 |
| Oligodendrocyte | 0.19 |
| Pineal gland | 0.22 |
| Radial glia | 0.37 |
| Schwann cell | 0.19 |
| Subcommissural organ | 0.16 |
| Vascular | 0.23 |

protein phosphatase 1D magnesium-dependent, delta isoform
[Source:MGI Symbol;Acc:MGI:1858214]
Accession: ENSMUSG00000020525 NCBI Gene
RefSeqID: EntrezID: 53892
Chr11 + 85311244 to 85347066 UCSC Browser

Not the most enriched gene in any cluster

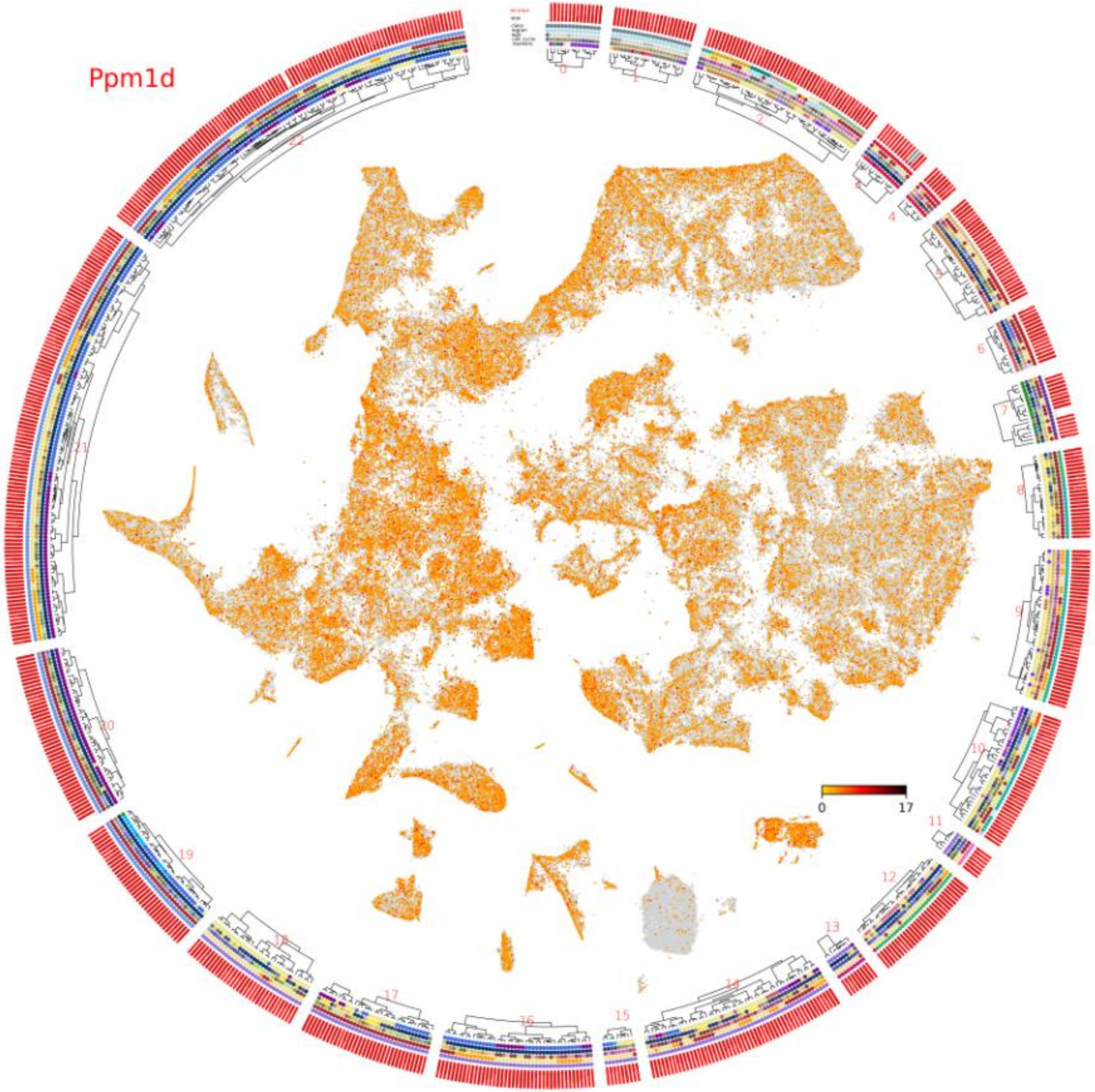

HELP Start

Class Age UMI Cycle Region Marker

Gene: Plcg2 Search

Grin2a Nlrc4 Plcg2 Clear buttons

Co-Expr: Max 5 genes T>0.95/T<0.05(-)

**Gene: Plcg2** Hide ClassExpr

| | |
|---|---|
| Bad cells | 0.02 |
| Blood | 0.01 |
| Choroid plexus | 0.02 |
| Ectoderm | 0.07 |
| Endoderm | 0.05 |
| Ependymal | 0.03 |
| Fibroblast | 0.01 |
| Gastrulation | 0.09 |
| Glioblast | 0.00 |
| Immune | 0.43 |
| Mesenchyme | 0.12 |
| Mesoderm | 0.03 |
| Neural crest | 0.72 |
| Neural tube | 0.05 |
| Neuroblast | 0.00 |
| Neuron | 0.00 |
| Olfactory ensheathing cell | 0.00 |
| Oligodendrocyte | 0.04 |
| Pineal gland | 0.00 |
| Radial glia | 0.01 |
| Schwann cell | 0.00 |
| Subcommissural organ | 0.00 |
| Vascular | 0.05 |

phospholipase C, gamma 2 [Source:MGI Symbol;Acc:MGI:97616]
Accession: ENSMUSG00000034330 NCBI Gene
RefSeqID: NM_172285 EntrezID: 234779
Chr8 + 117498291 to 117635142 UCSC Browser

Not the most enriched gene in any cluster

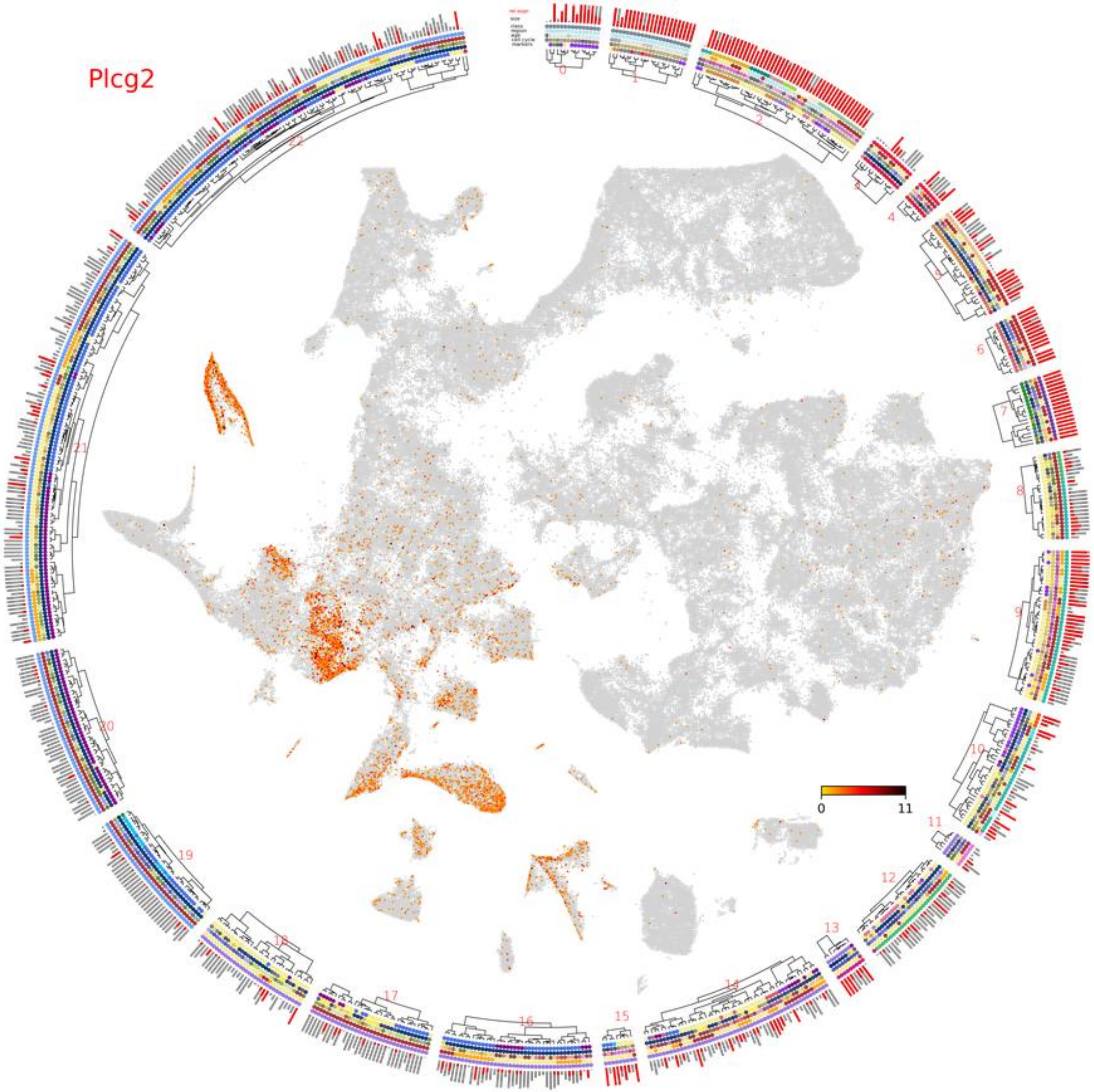

HELP Start

Class Age UMI Cycle Region Marker

Gene: Chek2 Search

Grin2a Nlrc4 Plcg2 Chek2 Clear buttons

Co-Expr: Max 5 genes T>0.95/T<0.05(-)

**Gene: Chek2** Hide ClassExpr

| | |
|---|---|
| Bad cells | 0.09 |
| Blood | 0.06 |
| Choroid plexus | 0.04 |
| Ectoderm | 0.21 |
| Endoderm | 0.22 |
| Ependymal | 1.09 |
| Fibroblast | 0.04 |
| Gastrulation | 0.26 |
| Glioblast | 0.09 |
| Immune | 0.10 |
| Mesenchyme | 0.23 |
| Mesoderm | 0.23 |
| Neural crest | 0.31 |
| Neural tube | 0.36 |
| Neuroblast | 0.07 |
| Neuron | 0.00 |
| Olfactory ensheathing cell | 0.04 |
| Oligodendrocyte | 0.06 |
| Pineal gland | 0.11 |
| Radial glia | 0.24 |
| Schwann cell | 0.05 |
| Subcommissural organ | 0.49 |
| Vascular | 0.13 |

checkpoint kinase 2 [Source:MGI Symbol;Acc:MGI:1355321]
Accession: ENSMUSG00000029521 NCBI Gene
RefSeqID: EntrezID: 50883
Chr5 + 110839979 to 110874145 UCSC Browser

Not the most enriched gene in any cluster

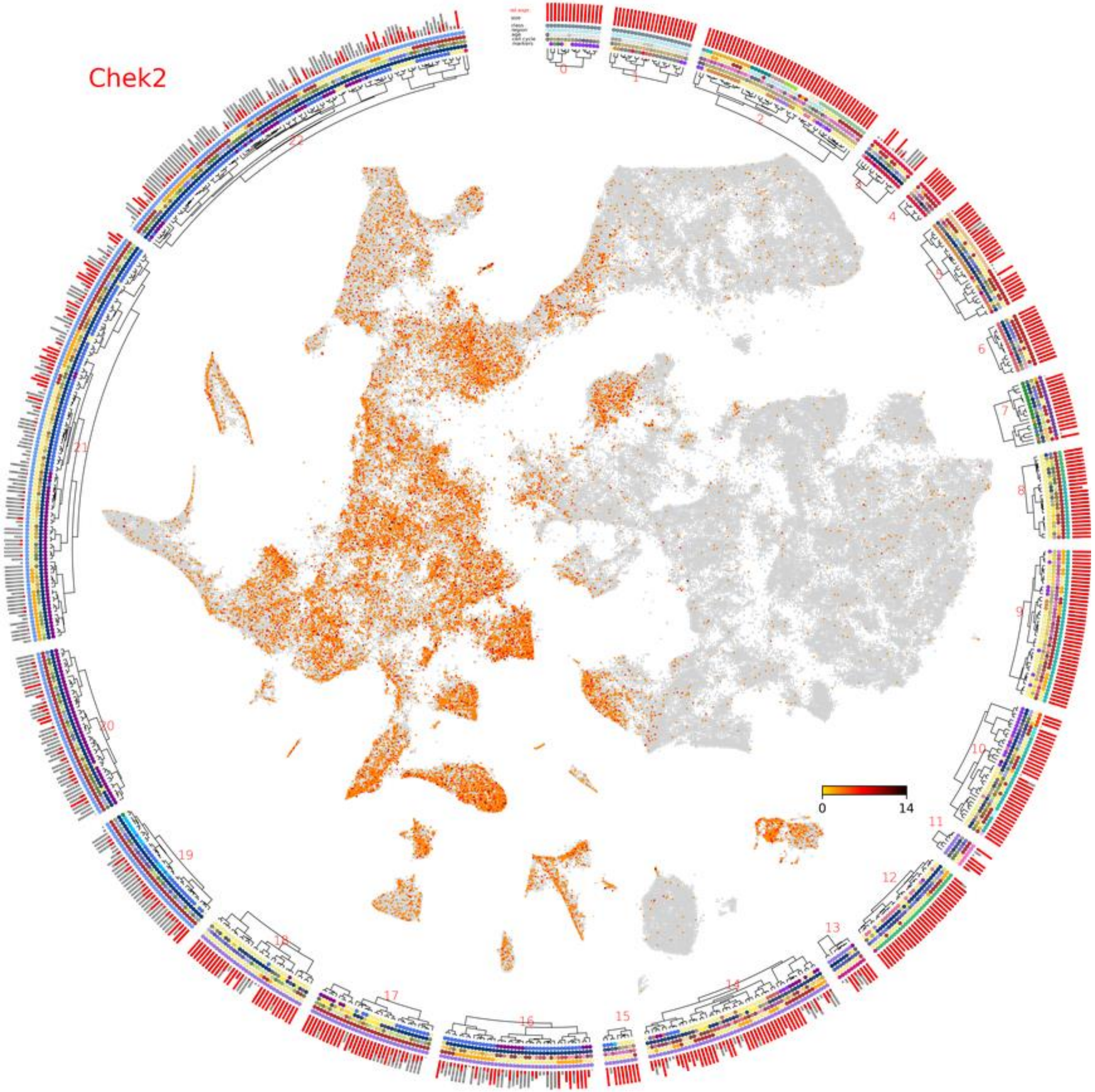

HELP Start

Class Age UMI Cycle Region Marker

Gene: Rag1 Search

Rag1 Clear buttons

Co-Expr: Max 5 genes T>0.95/T<0.05(-)

**Gene: Rag1** Hide ClassExpr

| | |
|---|---|
| Bad cells | 0.00 |
| Blood | 0.00 |
| Choroid plexus | 0.00 |
| Ectoderm | 0.00 |
| Endoderm | 0.00 |
| Ependymal | 0.00 |
| Fibroblast | 0.00 |
| Gastrulation | 0.00 |
| Glioblast | 0.00 |
| Immune | 0.00 |
| Mesenchyme | 0.00 |
| Mesoderm | 0.00 |
| Neural crest | 0.00 |
| Neural tube | 0.00 |
| Neuroblast | 0.00 |
| Neuron | 0.00 |
| Olfactory ensheathing cell | 0.00 |
| Oligodendrocyte | 0.00 |
| Pineal gland | 0.00 |
| Radial glia | 0.00 |
| Schwann cell | 0.00 |
| Subcommissural organ | 0.00 |
| Vascular | 0.00 |

recombination activating 1 [Source:MGI Symbol;Acc:MGI:97848]
Accession: ENSMUSG00000061311 NCBI Gene
RefSeqID: NM_009019 EntrezID: 19373
Chr2 - 101638282 to 101649501 UCSC Browser

Not the most enriched gene in any cluster

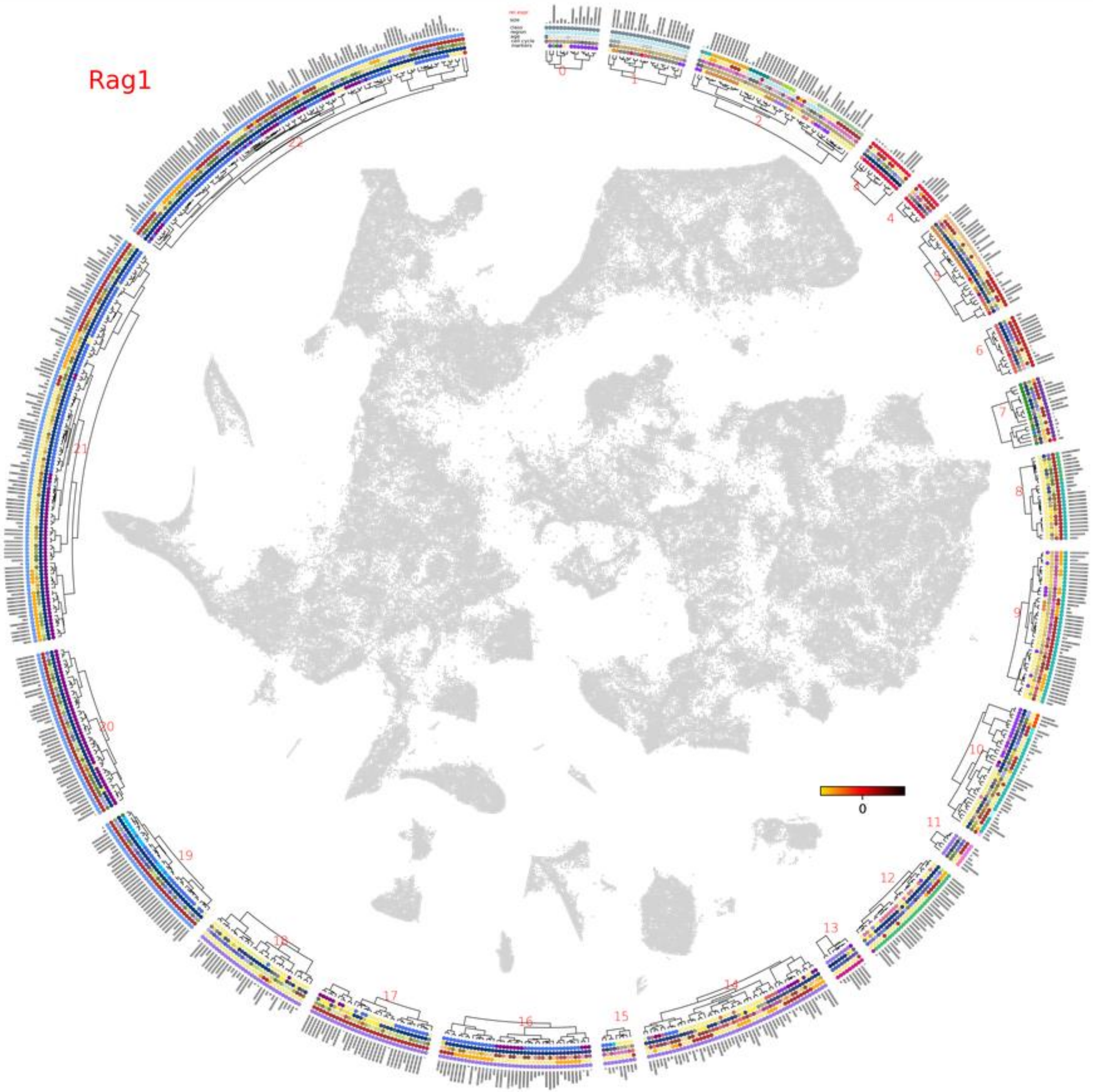
