## supplemental text for "Identification of ultra-rare genetic variants in Pediatric Acute Onset Neuropsychiatric Syndrome (PANS) by exome and whole genome sequencing"

**Supplemental Figure Legends**

**Figure S1: Single cell RNA-seq (scRNA-seq) peripheral blood cells.**

**Figure S2: Expression pattern of each of the PANS candidate genes in normal human tissues.** Composite image of gene expression across multiple tissues using the GTEx dataset

**Figure S3: Wheel plots showing single cell expression data from the developing mouse brain** ([http://mousebrain.org](http://mousebrain.org/)). The expression pattern is displayed as a UMAP clusters of2 different cell types determined by the expression pattern of cell specific markers. Slides 2 and 3 show the different cell types making up the clusters, while slide 4 shows the brain regions covered by the clusters. Slide 5 shows an example of a gene that is expressed in a cell type-specific manner, which was used to create the gene clusters. Slides 6-16 show the expression pattern at a common developmental stage. The box on the left of each wheel plot is the relative expression level within the different cell types.

**Supplemental Tables**

**Table S1: abbreviation key to Figure 4 (mouse adolescent brain single cell RNA-seq**

1	TEGLU1 <http://mousebrain.org/celltypes/TEGLU1>	Excitatory neurons, cerebral cortex

2	TEGLU3 <http://mousebrain.org/celltypes/TEGLU3>	Excitatory neurons, cerebral cortex

3	TEGLU2 <http://mousebrain.org/celltypes/TEGLU2>	Excitatory neurons, cerebral cortex

4	TEGLU20 <http://mousebrain.org/celltypes/TEGLU20>	Excitatory neurons, cerebral cortex

5	TEGLU11 <http://mousebrain.org/celltypes/TEGLU11>	Excitatory neurons, cerebral cortex

6	TEGLU12 <http://mousebrain.org/celltypes/TEGLU12>	Excitatory neurons, cerebral cortex

7	TEGLU10 <http://mousebrain.org/celltypes/TEGLU10>	Excitatory neurons, cerebral cortex

8	TEGLU9 <http://mousebrain.org/celltypes/TEGLU9>	Excitatory neurons, cerebral cortex

9	TEGLU8 <http://mousebrain.org/celltypes/TEGLU8>	Excitatory neurons, cerebral cortex

10	TEGLU7 <http://mousebrain.org/celltypes/TEGLU7>	Excitatory neurons, cerebral cortex

11	TEGLU6 <http://mousebrain.org/celltypes/TEGLU6>	Excitatory neurons, cerebral cortex

12	TEGLU13 <http://mousebrain.org/celltypes/TEGLU13>	Excitatory neurons, cerebral cortex

13	TEGLU14 <http://mousebrain.org/celltypes/TEGLU14>	Excitatory neurons, cerebral cortex

14	TEGLU5 <http://mousebrain.org/celltypes/TEGLU5>	Excitatory neurons, cerebral cortex

15	TEGLU16 <http://mousebrain.org/celltypes/TEGLU16>	Excitatory neurons, cerebral cortex

16	TEGLU15 <http://mousebrain.org/celltypes/TEGLU15>	Excitatory neurons, cerebral cortex

17	TEGLU17 <http://mousebrain.org/celltypes/TEGLU17>	Excitatory neurons, cerebral cortex

18	TEGLU18 <http://mousebrain.org/celltypes/TEGLU18>	Excitatory neurons, cerebral cortex

19	TEGLU19 <http://mousebrain.org/celltypes/TEGLU19>	Excitatory neurons, cerebral cortex

20	TEGLU22 <http://mousebrain.org/celltypes/TEGLU22>	Excitatory neurons, amygdala

21	TEGLU21 <http://mousebrain.org/celltypes/TEGLU21>	Excitatory neurons, hippocampus CA1

22	TEGLU4 <http://mousebrain.org/celltypes/TEGLU4>	Excitatory neurons, cerebral cortex

23	TEGLU24 <http://mousebrain.org/celltypes/TEGLU24>	Excitatory neurons, hippocampus CA1

24	TEGLU23 <http://mousebrain.org/celltypes/TEGLU23>	Excitatory neurons, hippocampus CA3

25	DGGRC1 <http://mousebrain.org/celltypes/DGGRC1>	Granule neuroblasts, dentate gyrus

26	DGGRC2 <http://mousebrain.org/celltypes/DGGRC2>	Granule neurons, dentate gyrus

27	MSN1 <http://mousebrain.org/celltypes/MSN1>	D1 medium spiny neurons, striatum

28	MSN2 <http://mousebrain.org/celltypes/MSN2>	D2 medium spiny neurons, striatum

29	MSN3 <http://mousebrain.org/celltypes/MSN3>	D2 medium spiny neurons, striatum

30	MSN4 <http://mousebrain.org/celltypes/MSN4>	D1 medium spiny neurons, striatum

31	MSN5 <http://mousebrain.org/celltypes/MSN5>	Patch D1/D2 neurons, striatum

32	MSN6 <http://mousebrain.org/celltypes/MSN6>	Matrix D1 neurons, striatum

33	DETPH <http://mousebrain.org/celltypes/DETPH>	Neuroblast-like, habenula

34	DGNBL2 <http://mousebrain.org/celltypes/DGNBL2>	Granule neuroblasts, dentate gyrus

35	DGNBL1 <http://mousebrain.org/celltypes/DGNBL1>	Granule neuroblasts, dentate gyrus

36	SZNBL <http://mousebrain.org/celltypes/SZNBL>	Neuronal intermidate progenitor cells

37	OBNBL3 <http://mousebrain.org/celltypes/OBNBL3>	Neuroblasts, olfactory bulb

38	OBINH1 <http://mousebrain.org/celltypes/OBINH1>	Inhibitory neurons, olfactory bulb

39	OBINH5 <http://mousebrain.org/celltypes/OBINH5>	Inhibitory neurons, olfactory nulb

40	OBINH2 <http://mousebrain.org/celltypes/OBINH2>	Inner horizontal cell, olfactory bulb

41	OBINH3 <http://mousebrain.org/celltypes/OBINH3>	Inhibitory neurons, olfactory bulb

42	OBINH4 <http://mousebrain.org/celltypes/OBINH4>	Inhibitory neurons, olfactory bulb

43	OBNBL4 <http://mousebrain.org/celltypes/OBNBL4>	Inhibitory neurons, olfactory bulb

44	OBNBL5 <http://mousebrain.org/celltypes/OBNBL5>	Inhibitory neurons, olfactory bulb

45	OBDOP <http://mousebrain.org/celltypes/OBDOP>	Dopaminergic periglomerular interneuron, olfactory bulb

46	OBINH6 <http://mousebrain.org/celltypes/OBINH6>	External plexiform layer interneuron, olfactory bulb

47	DEINH1 <http://mousebrain.org/celltypes/DEINH1>	Inhibitory neurons, thalamus

48	DEINH2 <http://mousebrain.org/celltypes/DEINH2>	Inhibitory neurons, thalamus

49	TEINH17 <http://mousebrain.org/celltypes/TEINH17>	Axo-axonic, cortex/hippocampus

50	TEINH18 <http://mousebrain.org/celltypes/TEINH18>	Basket and bistratified cells, cortex/hippocampus

51	TEINH19 <http://mousebrain.org/celltypes/TEINH19>	Hippocamposeptal projection, cortex/hippocampus

52	TEINH21 <http://mousebrain.org/celltypes/TEINH21>	Sleep-active, long-range projection interneurons, cortex/hippocampus

53	TEINH16 <http://mousebrain.org/celltypes/TEINH16>	Ivy and MGE-derived neurogliaform cells, cortex/hippocampus

54	TEINH15 <http://mousebrain.org/celltypes/TEINH15>	CGE-derived neurogliaform cells, cortex/hippocampus

55	TEINH14 <http://mousebrain.org/celltypes/TEINH14>	CGE-derived neurogliaform cells Cxcl14+, cortex/hippocampus

56	TEINH20 <http://mousebrain.org/celltypes/TEINH20>	Inhibitory interneurons, hippocampus

57	TEINH13 <http://mousebrain.org/celltypes/TEINH13>	Trilaminar cells, hippocampus

58	TEINH12 <http://mousebrain.org/celltypes/TEINH12>	Non-border Cck interneurons, cortex/hippocampus

59	TEINH9 <http://mousebrain.org/celltypes/TEINH9>	Non-border Cck interneurons, hippocampus

60	TEINH10 <http://mousebrain.org/celltypes/TEINH10>	R-LM border Cck interneurons, cortex/hippocampus

61	TEINH11 <http://mousebrain.org/celltypes/TEINH11>	R-LM border Cck interneurons, cortex/hippocampus

62	TEINH4 <http://mousebrain.org/celltypes/TEINH4>	Interneuron-selective interneurons, cortex/hippocampus

63	TEINH5 <http://mousebrain.org/celltypes/TEINH5>	Interneuron-selective interneurons, cortex/hippocampus

64	TEINH8 <http://mousebrain.org/celltypes/TEINH8>	Interneuron-selective interneurons, hippocampus

65	TEINH7 <http://mousebrain.org/celltypes/TEINH7>	Interneuron-selective interneurons, hippocampus

66	TEINH6 <http://mousebrain.org/celltypes/TEINH6>	Interneuron-selective interneurons, cortex/hippocampus

67	TECHO <http://mousebrain.org/celltypes/TECHO>	Cholinergic interneurons, telencephalon

68	DECHO1 <http://mousebrain.org/celltypes/DECHO1>	Cholinergic neurons, septal nucleus, Meissnert and diagonal band

69	HBCHO4 <http://mousebrain.org/celltypes/HBCHO4>	Afferent nuclei of cranial nerves III-V

70	HBCHO3 <http://mousebrain.org/celltypes/HBCHO3>	Afferent nuclei of cranial nerves VI-XII

71	HBADR <http://mousebrain.org/celltypes/HBADR>	Adrenergic cell groups of the medulla

72	HBNOR <http://mousebrain.org/celltypes/HBNOR>	Noradrenergic neurons of the medulla

73	HYPEP7 <http://mousebrain.org/celltypes/HYPEP7>	Pmch neurons, hypothalamus

74	HYPEP6 <http://mousebrain.org/celltypes/HYPEP6>	Orexin-producing neurons, hypothalamus

75	MEGLU14 <http://mousebrain.org/celltypes/MEGLU14>	Glutamatergic projection neurons of the raphe nucleus

76	MBDOP1 <http://mousebrain.org/celltypes/MBDOP1>	Dopaminergic neurons, periaqueductal grey

77	MBDOP2 <http://mousebrain.org/celltypes/MBDOP2>	Dopaminergic neurons, ventral midbrain (SNc, VTA)

78	HBSER1 <http://mousebrain.org/celltypes/HBSER1>	Serotonergic neurons, hindbrain

79	HBSER2 <http://mousebrain.org/celltypes/HBSER2>	Serotonergic neurons, hindbrain

80	HBSER3 <http://mousebrain.org/celltypes/HBSER3>	Serotonergic neurons, hindbrain

81	HBSER5 <http://mousebrain.org/celltypes/HBSER5>	Serotonergic neurons, hindbrain

82	HBSER4 <http://mousebrain.org/celltypes/HBSER4>	Serotonergic neurons, hindbrain

83	TEINH3 <http://mousebrain.org/celltypes/TEINH3>	Inhibitory neurons, telencephalon

84	TEINH2 <http://mousebrain.org/celltypes/TEINH2>	Inhibitory neurons, septal nucleus

85	DEINH4 <http://mousebrain.org/celltypes/DEINH4>	Inhibitory neurons, thalamus

86	DEINH5 <http://mousebrain.org/celltypes/DEINH5>	Peptidergic neurons, hypothalamus

87	HYPEP3 <http://mousebrain.org/celltypes/HYPEP3>	Peptidergic neurons, hypothalamus

88	HYPEP1 <http://mousebrain.org/celltypes/HYPEP1>	Peptidergic neurons, hypothalamus

89	HYPEP2 <http://mousebrain.org/celltypes/HYPEP2>	Peptidergic neurons, hypothalamus

90	MEINH14 <http://mousebrain.org/celltypes/MEINH14>	Inhibitory neurons, midbrain

91	DEINH6 <http://mousebrain.org/celltypes/DEINH6>	Peptidergic neurons, hypothalamus

92	DEINH8 <http://mousebrain.org/celltypes/DEINH8>	Interneurons, hypothalamus

93	DEINH7 <http://mousebrain.org/celltypes/DEINH7>	Inhibitory neurons, hypothalamus

94	HYPEP5 <http://mousebrain.org/celltypes/HYPEP5>	Vasopressin-producing cells, hypothalamus

95	HYPEP4 <http://mousebrain.org/celltypes/HYPEP4>	Oxytocin-producing cells, hypothalamus

96	HYPEP8 <http://mousebrain.org/celltypes/HYPEP8>	Peptidergic neurons, hypothalamus

97	SCINH11 <http://mousebrain.org/celltypes/SCINH11>	Central canal neurons, spinal cord

98	SCINH10 <http://mousebrain.org/celltypes/SCINH10>	Inhibitory neurons, spinal cord

99	SCINH9 <http://mousebrain.org/celltypes/SCINH9>	Inhibitory neurons, spinal cord

100	SCINH8 <http://mousebrain.org/celltypes/SCINH8>	Inhibitory neurons, spinal cord

101	SCINH7 <http://mousebrain.org/celltypes/SCINH7>	Inhibitory neurons, spinal cord

102	SCINH6 <http://mousebrain.org/celltypes/SCINH6>	Inhibitory neurons, spinal cord

103	SCINH5 <http://mousebrain.org/celltypes/SCINH5>	Inhibitory neurons, spinal cord

104	SCINH4 <http://mousebrain.org/celltypes/SCINH4>	Inhibitory neurons, spinal cord

105	SCINH3 <http://mousebrain.org/celltypes/SCINH3>	Inhibitory neurons, spinal cord

106	HBINH9 <http://mousebrain.org/celltypes/HBINH9>	Inhibitory neurons, hindbrain

107	SCINH2 <http://mousebrain.org/celltypes/SCINH2>	Inhibitory neurons, spinal cord

108	SCGLU1 <http://mousebrain.org/celltypes/SCGLU1>	Excitatory neurons, spinal cord

109	SCGLU2 <http://mousebrain.org/celltypes/SCGLU2>	Excitatory neurons, spinal cord

110	SCGLU3 <http://mousebrain.org/celltypes/SCGLU3>	Excitatory neurons, spinal cord

111	SCGLU4 <http://mousebrain.org/celltypes/SCGLU4>	Excitatory neurons, spinal cord

112	SCGLU5 <http://mousebrain.org/celltypes/SCGLU5>	Excitatory neurons, spinal cord

113	SCGLU6 <http://mousebrain.org/celltypes/SCGLU6>	Excitatory neurons, spinal cord

114	SCGLU7 <http://mousebrain.org/celltypes/SCGLU7>	Excitatory neurons, spinal cord

115	SCGLU8 <http://mousebrain.org/celltypes/SCGLU8>	Excitatory neurons, spinal cord

116	SCGLU9 <http://mousebrain.org/celltypes/SCGLU9>	Excitatory neurons, spinal cord

117	SCGLU10 <http://mousebrain.org/celltypes/SCGLU10>	Excitatory neurons, spinal cord

118	HBGLU10 <http://mousebrain.org/celltypes/HBGLU10>	Excitatory neurons, hindbrain

119	HBGLU3 <http://mousebrain.org/celltypes/HBGLU3>	Excitatory neurons, hindbrain

120	HBGLU2 <http://mousebrain.org/celltypes/HBGLU2>	Excitatory neurons, hindbrain

121	MEGLU2 <http://mousebrain.org/celltypes/MEGLU2>	Excitatory neurons, midbrain

122	MEGLU3 <http://mousebrain.org/celltypes/MEGLU3>	Excitatory neurons, midbrain

123	DEGLU5 <http://mousebrain.org/celltypes/DEGLU5>	Excitatory neurons, midbrain

124	MEGLU1 <http://mousebrain.org/celltypes/MEGLU1>	Excitatory neurons, midbrain

125	MEGLU7 <http://mousebrain.org/celltypes/MEGLU7>	Excitatory neurons, midbrain

126	MEGLU8 <http://mousebrain.org/celltypes/MEGLU8>	Excitatory neurons, midbrain

127	MEGLU9 <http://mousebrain.org/celltypes/MEGLU9>	Excitatory neurons, midbrain

128	MEGLU10 <http://mousebrain.org/celltypes/MEGLU10>	Excitatory neurons, midbrain

129	MEGLU11 <http://mousebrain.org/celltypes/MEGLU11>	Excitatory neurons, midbrain

130	MBCHO1 <http://mousebrain.org/celltypes/MBCHO1>	Cholinergic neurons, midbrain red nucleus 131	MEGLU6 <http://mousebrain.org/celltypes/MEGLU6>	Excitatory neurons, midbrain

132	MEGLU5 <http://mousebrain.org/celltypes/MEGLU5>	Excitatory neurons, midbrain

133	MEGLU4 <http://mousebrain.org/celltypes/MEGLU4>	Excitatory neurons, midbrain

134	CR <http://mousebrain.org/celltypes/CR>	Cajal-Retzius cells, hippocampus

135	DECHO2 <http://mousebrain.org/celltypes/DECHO2>	Cholinergic neurons, habenula

136	HBGLU1 <http://mousebrain.org/celltypes/HBGLU1>	Excitatory neurons, hindbrain

137	DEGLU1 <http://mousebrain.org/celltypes/DEGLU1>	Excitatory neurons, thalamus

138	DEGLU2 <http://mousebrain.org/celltypes/DEGLU2>	Excitatory neurons, hypothalamus

139	DEGLU3 <http://mousebrain.org/celltypes/DEGLU3>	Excitatory neurons, thalamus

140	DEGLU4 <http://mousebrain.org/celltypes/DEGLU4>	Excitatory neurons, thalamus

141	MEINH12 <http://mousebrain.org/celltypes/MEINH12>	Inhibitory neurons, midbrain

142	MEINH11 <http://mousebrain.org/celltypes/MEINH11>	Inhibitory neurons, midbrain

143	MEINH10 <http://mousebrain.org/celltypes/MEINH10>	Inhibitory neurons, midbrain

144	MEINH9 <http://mousebrain.org/celltypes/MEINH9>	Inhibitory neurons, midbrain

145	MEINH5 <http://mousebrain.org/celltypes/MEINH5>	Inhibitory neurons, midbrain

146	MEINH6 <http://mousebrain.org/celltypes/MEINH6>	Inhibitory neurons, midbrain

147	MEINH7 <http://mousebrain.org/celltypes/MEINH7	Inhibitory neurons, midbrain

148	MEINH4 <http://mousebrain.org/celltypes/MEINH4>	Inhibitory neurons, midbrain

149	MEINH3 <http://mousebrain.org/celltypes/MEINH3>	Inhibitory neurons, midbrain

150	HBINH5 <http://mousebrain.org/celltypes/HBINH5>	Inhibitory neurons, hindbrain

151	MEINH2 <http://mousebrain.org/celltypes/MEINH2>	Inhibitory neurons, midbrain

152	DEINH3 <http://mousebrain.org/celltypes/DEINH3>	Inhibitory neurons, hypothalamus

153	TEINH1 <http://mousebrain.org/celltypes/TEINH1>	Inhibitory neurons, pallidum

154	MEINH13 <http://mousebrain.org/celltypes/MEINH13>	Inhibitory neurons, midbrain

155	MEINH8 <http://mousebrain.org/celltypes/MEINH8>	Inhibitory neurons, midbrain

156	HBINH1 <http://mousebrain.org/celltypes/HBINH1>	Inhibitory neurons, hindbrain

157	HBINH3 <http://mousebrain.org/celltypes/HBINH3>	Inhibitory neurons, hindbrain

158	HBINH4 <http://mousebrain.org/celltypes/HBINH4>	Inhibitory neurons, hindbrain

159	HBINH6 <http://mousebrain.org/celltypes/HBINH6>	Inhibitory neurons, hindbrain

160	HBINH2 <http://mousebrain.org/celltypes/HBINH2>	Inhibitory neurons, hindbrain

161	HBCHO1 <http://mousebrain.org/celltypes/HBCHO1>	Cholinergic neurons, hindbrain

162	HBCHO2 <http://mousebrain.org/celltypes/HBCHO2>	Cholinergic neurons, hindbrain

163	HBGLU4 <http://mousebrain.org/celltypes/HBGLU4>	Excitatory neurons, hindbrain

164	HBGLU5 <http://mousebrain.org/celltypes/HBGLU5>	Excitatory neurons, hindbrain

165	HBGLU6 <http://mousebrain.org/celltypes/HBGLU6>	Excitatory neurons, hindbrain

166	HBGLU7 <http://mousebrain.org/celltypes/HBGLU7>	Excitatory neurons, hindbrain

167	HBGLU8 <http://mousebrain.org/celltypes/HBGLU8>	Excitatory neurons, hindbrain

168	HBGLU9 <http://mousebrain.org/celltypes/HBGLU9>	Excitatory neurons, hindbrain

169	HBINH7 <http://mousebrain.org/celltypes/HBINH7>	Inhibitory neurons, hindbrain

170	HBINH8 <http://mousebrain.org/celltypes/HBINH8>	Inhibitory neurons, hindbrain

171	SCINH1 <http://mousebrain.org/celltypes/SCINH1>	Inhibitory neurons, spinal cord

172	CBINH2 <http://mousebrain.org/celltypes/CBINH2>	Granular layer interneurons, cerebellum

173	MEINH1 <http://mousebrain.org/celltypes/MEINH1>	Inhibitory neurons, midbrain

174	CBINH1 <http://mousebrain.org/celltypes/CBINH1>	Molecular layer interneurons, cerebellum

175	CBPC <http://mousebrain.org/celltypes/CBPC>	Purkinje cells

176	CBGRC <http://mousebrain.org/celltypes/CBGRC>	Granule neurons, cerebellum

177	CBNBL2 <http://mousebrain.org/celltypes/CBNBL2>	Neuroblasts, cerebellum

178	CBNBL1 <http://mousebrain.org/celltypes/CBNBL1>	Neuroblasts, cerebellum

179	SEPNBL <http://mousebrain.org/celltypes/SEPNBL>	Neuroblasts, septum

180	OBNBL1 <http://mousebrain.org/celltypes/OBNBL1>	Neuroblasts, olfactory

181	OBNBL2 <http://mousebrain.org/celltypes/OBNBL2>	Neuroblasts, olfactory bulb

182	ENT1 <http://mousebrain.org/celltypes/ENT1>	Nitrergic enteric neurons

183	ENT2 <http://mousebrain.org/celltypes/ENT2>	Nitrergic enteric neurons

184	ENT3 <http://mousebrain.org/celltypes/ENT3>	Nitrergic enteric neurons

185	ENT4 <http://mousebrain.org/celltypes/ENT4>	Cholinergic enteric neurons

186	ENT5 <http://mousebrain.org/celltypes/ENT5>	Cholinergic enteric neurons

187	ENT6 <http://mousebrain.org/celltypes/ENT6>	Cholinergic enteric neurons

188	ENT7 <http://mousebrain.org/celltypes/ENT7>	Cholinergic enteric neurons, VGLUT2

189	ENT8 <http://mousebrain.org/celltypes/ENT8>	Cholinergic enteric neurons, VGLUT2

190	ENT9 <http://mousebrain.org/celltypes/ENT9>	Cholinergic enteric neurons

191	SYNOR1 <http://mousebrain.org/celltypes/SYNOR1>	Noradrenergic erector muscle neurons

192	SYNOR2 <http://mousebrain.org/celltypes/SYNOR2>	Noradrenergic neurons, sympathetic

193	SYNOR3 <http://mousebrain.org/celltypes/SYNOR3>	Noradrenergic neurons, sympathetic

194	SYNOR4 <http://mousebrain.org/celltypes/SYNOR4>	Noradrenergic erector muscle neurons	195	SYNOR5 <http://mousebrain.org/celltypes/SYNOR5>	Noradrenergic erector muscle neurons

196	SYCHO2 <http://mousebrain.org/celltypes/SYCHO2>	Cholinergic neurons, sympathetic

197	SYCHO1 <http://mousebrain.org/celltypes/SYCHO1>	Cholinergic neurons, sympathetic

198	PSPEP8 <http://mousebrain.org/celltypes/PSPEP8>	Peptidergic (TrpM8), DRG

199	PSPEP7 <http://mousebrain.org/celltypes/PSPEP7>	Peptidergic (TrpM8), DRG

200	PSPEP6 <http://mousebrain.org/celltypes/PSPEP6>	Peptidergic (TrpM8), DRG

201	PSPEP5 <http://mousebrain.org/celltypes/PSPEP5>	Peptidergic (PEP1.2), DRG

202	PSPEP2 <http://mousebrain.org/celltypes/PSPEP2>	Peptidergic (PEP1.3), DRG

203	PSPEP4 <http://mousebrain.org/celltypes/PSPEP4>	Peptidergic (PEP1.1), DRG

204	PSPEP3 <http://mousebrain.org/celltypes/PSPEP3>	Peptidergic (PEP1..4), DRG

205	PSPEP1 <http://mousebrain.org/celltypes/PSPEP1>	Peptidergicv (PEP2), DRG

206	PSNF3 <http://mousebrain.org/celltypes/PSNF3>	Neurofilament (NF2/3), DRG

207	PSNF2 <http://mousebrain.org/celltypes/PSNF2>	Neurofilament (NF4/5), DRG

208	PSNF1 <http://mousebrain.org/celltypes/PSNF1>	Neurofilament (NF1), DRG

209	PSNP1 <http://mousebrain.org/celltypes/PSNP1>	Non-peptidergic (TH), DRG

210	PSNP2 <http://mousebrain.org/celltypes/PSNP2>	Non-peptidergic (NP1.1), DRG

211	PSNP3 <http://mousebrain.org/celltypes/PSNP3>	Non-peptidergic (NP1.2), DRG

212	PSNP4 <http://mousebrain.org/celltypes/PSNP4>	Non-peptidergic (NP2.1), DRG

213	PSNP5 <http://mousebrain.org/celltypes/PSNP5>	Non-peptidergic (NP2.2), DRG

214	PSNP6 <http://mousebrain.org/celltypes/PSNP6>	Non-peptidergic (NP3), DRG

215	COP1 <http://mousebrain.org/celltypes/COP1>	Committed oligodendrocytes cells (COP)

216	COP2 <http://mousebrain.org/celltypes/COP2>	Committed oligodendrocytes cells (COP), pons/medulla specific

217	NFOL2 <http://mousebrain.org/celltypes/NFOL2>	Newly formed oligodendrocytes (NFOL), pons/medulla specific

218	NFOL1 <http://mousebrain.org/celltypes/NFOL1>	Newly formed oligodendrocytes (NFOL)

219	MFOL2 <http://mousebrain.org/celltypes/MFOL2>	Myelin forming oligodendrocytes (MFOL)

220	MFOL1 <http://mousebrain.org/celltypes/MFOL1>	Myelin forming oligodendrocytes (MFOL)

221	MOL1 <http://mousebrain.org/celltypes/MOL1>	Mature oligodendrocytes

222	MOL2 <http://mousebrain.org/celltypes/MOL2>	Mature oligodendrocytes, hindbrain

223	MOL3 <http://mousebrain.org/celltypes/MOL3>	Mature oligodendrocytes, spinal cord enriched (high Klk6)

224	CHOR <http://mousebrain.org/celltypes/CHOR>	Chorid plexus epithelial cells

225	HYPEN <http://mousebrain.org/celltypes/HYPEN>	Hypendymal cell, subcommissural organ

226	EPSC <http://mousebrain.org/celltypes/EPSC>	Ependymal cells, spinal cord

227	EPEN <http://mousebrain.org/celltypes/EPEN>	Ependymal cells

228	EPMB <http://mousebrain.org/celltypes/EPMB>	Ependymal cells, midbrain

229	RGDG <http://mousebrain.org/celltypes/RGDG>	Dentate gyrus radial glia-like cells

230	RGSZ <http://mousebrain.org/celltypes/RGSZ>	Subventricular zone radial glia-like cells

231	ACTE1 <http://mousebrain.org/celltypes/ACTE1>	Telencephalon astrocytes, fibrous

232	ACTE2 <http://mousebrain.org/celltypes/ACTE2>	Telencephalon astrocytes, protoplasmic

233	ACOB <http://mousebrain.org/celltypes/ACOB>	Olfactory astrocytes

234	ACNT1 <http://mousebrain.org/celltypes/ACNT1>	Non-telencephalon astrocytes, protoplasmic 235	ACNT2 <http://mousebrain.org/celltypes/ACNT2>	Non-telencephalon astrocytes, fibrous

236	ACMB <http://mousebrain.org/celltypes/ACMB>	Dorsal midbrain Myoc-expressing astrocyte-like

237	ACBG <http://mousebrain.org/celltypes/ACBG>	Bergmann glia

238	OEC <http://mousebrain.org/celltypes/OEC>	Olfactory ensheathing cells

239	OPC <http://mousebrain.org/celltypes/OPC>	Oligodendrocytes precursor cells

240	SCHW <http://mousebrain.org/celltypes/SCHW>	Schwann cells

241	SATG2 <http://mousebrain.org/celltypes/SATG2>	Satellite glia

242	SATG1 <http://mousebrain.org/celltypes/SATG1>	Satellite glia, proliferating

243	ENTG1 <http://mousebrain.org/celltypes/ENTG1>	Enteric glia, proliferating

244	ENTG2 <http://mousebrain.org/celltypes/ENTG2>	Enteric glia

245	ENTG3 <http://mousebrain.org/celltypes/ENTG3>	Enteric glia

246	ENTG4 <http://mousebrain.org/celltypes/ENTG4>	Enteric glia

247	ENTG5 <http://mousebrain.org/celltypes/ENTG5>	Enteric glia

248	ENTG6 <http://mousebrain.org/celltypes/ENTG6>	Enteric glia

249	ENTG7 <http://mousebrain.org/celltypes/ENTG7>	Enteric glia

250	ENMFB <http://mousebrain.org/celltypes/ENMFB>	Enteric mesothelial fibroblasts

251	ABC <http://mousebrain.org/celltypes/ABC>	Vascular leptomeningeal cells

252	VLMC2 <http://mousebrain.org/celltypes/VLMC2>	Vascular leptomeningeal cells

253	VLMC1 <http://mousebrain.org/celltypes/VLMC1>	Vascular leptomeningeal cells

254	VECA <http://mousebrain.org/celltypes/VECA>	Vascular endothelial cells, arterial

255	PER3 <http://mousebrain.org/celltypes/PER3>	Pericytes

256	VSMCA <http://mousebrain.org/celltypes/VSMCA>	Vascular smooth muscle cells, arterial

257	PER1 <http://mousebrain.org/celltypes/PER1>	Pericytes

258	PER2 <http://mousebrain.org/celltypes/PER2>	Pericytes, possibly mixed with VENC

259	VECC <http://mousebrain.org/celltypes/VECC>	Vascular endothelial cells, capillary

260	VECV <http://mousebrain.org/celltypes/VECV>	Vascular endothelial cells, venous

261	PVM1 <http://mousebrain.org/celltypes/PVM1>	Perivascular macrophages

262	PVM2 <http://mousebrain.org/celltypes/PVM2>	Perivascular macrophages, activated

263	MGL3 <http://mousebrain.org/celltypes/MGL3>	Microglia, activated

264	MGL2 <http://mousebrain.org/celltypes/MGL2>	Microglia, activated

265	MGL1 <http://mousebrain.org/celltypes/MGL1>	Microglia
